# Examining the relationships between psychological distress and financial hardship and uncertainty during the COVID-19 pandemic and the subsequent cost-of-living crisis in the United Kingdom

**DOI:** 10.1101/2024.01.14.24301283

**Authors:** Jacques Wels, Natasia Hamarat

## Abstract

**Background:** The study tests whether financial hardship and uncertainty have increased during the COVID-19 pandemic and the early stage of the subsequent cost-of-living crisis and whether these might explain increased psychological distress among the UK population.

**Methods:** We derive two cohorts from Understanding Society, a study representative of the UK population. Cohort 1 (C1) starts in 2016 and includes a 3-year follow-up until 2019. Cohort 2 (C2) starts in 2019 and ends in 2022. We provide descriptive statistics on financial hardship and uncertainty and apply parallel Latent Growth Modelling (LGM) on each cohort to explain variations in psychological distress (GHQ-36) based on baseline and follow-up financial trajectories. The sample is adjusted using cross-sectional weights and inverse probability weights for attrition.

**Results:** Financial hardship rates do not differ across cohorts but a marginal increase of 10 percent in financial uncertainty is observed in 2022 for C2. No significant difference in associations is observed across cohorts in the LGM with constant financial hardship increasing the GHQ-36 slope by 0.89 (95%CI=0.76;1.02) and 0.89 (95%CI=0.73;1.05) units in C1 and C2 and constant financial uncertainty increasing it respectively by 0.95 (95%CI=0.74;1.17) and 1.04 (95%CI=0.82;1.25). Baseline hardship and uncertainty increase the intercept by 2.39 (95%CI=2.11;2.67) and 1.74 (195%CI=1.38;2.10) in C1 and 2.97 (95%CI=2.65;3.29) and 2.12 (95%CI=1.75;2.49) in C2.

**Discussion:** The uncertainty caused by the 2022 cost-of-living crisis might have contributed to increase psychological distress within the UK population. Stronger detrimental effects might be expected if financial hardship were to increase.

## Background

Financial hardship is a crucial determinant of poor mental health and recent years, punctuated by a two-year-long pandemic and a subsequent and ongoing cost-of-living crisis, have significantly challenged both aspects.

Research on the relationship between financial wellbeing and mental health is sparse ^1^ and most studies usually look at financial wellbeing as an outcome and not as an explanatory variable ^2^. Among those looking at financial wellbeing as exposure, the association between poor financial wellbeing and poor mental health is consistent and is irrespective of how financial wellbeing is measured. A systematic review of 24 cross-sectional studies has found a moderate, yet positive association between financial satisfaction and subjective wellbeing ^3^ but longitudinal studies suggest stronger associations. For instance, Butterworth and al. ^4^ have demonstrated that financial hardship is much strongly associated with poor mental health outcomes than other socio-economic characteristics such as employment or education and that this association varies over time as past financial hardship only partially explain later mental health status. Financial hardship correlates with poor mental health and longitudinal measures of such a relationship show that those who have reported financial hardship in the past are more likely to report current mental health problems but mental health problems are greater when financial hardship is reported ^5^.

Such a perspective raises two major methodological issues. On the one hand, financial hardship can be derived from objective or subjective indicators, with a potential mismatch ^1^. What the literature shows is that, unlike objective measurements, financial strain is a robust predictor of worsening mental health ^6^. Studies indicate that all measures of financial wellbeing including financial capability, financial distress and financial security lead to the same effect in terms of physical and mental health ^7^ but with varying degrees of association. On the other hand, one must account for potential reverse causation ^8^ as those with poor mental health are more likely to face financial difficulties because of health care costs (that vary by country), low sick leave benefits or labour market consequences ^9^.

Beyond an individual perspective, structural contexts also affect the nature of this relationship. Economic crises have a true cost in terms of psychological wellbeing ^10^ but the role of contextual factors such as inflation or unemployment are not well known ^2^. Three recent events are of particular interest: the 2008 Great Recession, the 2020-2021 COVID-19 pandemic and the subsequent cost of living crisis. The impact of the 2008 Great Recession on (mental) health has been well documented ^11^ showing that men were more at risk of poor mental health during the crisis and that strong social security systems, particularly in Europe, may have mitigated the effect of Recession ^12^. Other studies have shown that life satisfaction appears to be uncorrelated with GDP growth and the effect of the crisis on overall life satisfaction is small ^13^. However, specific sub-populations were more at risk to be affected by the crisis with potential long-lasting effects on their mental health in post-recession times caused by housing, job-related and financial impacts ^14^. The setting was slightly different during the COVID-19 pandemic. Some countries have experienced economic shock and economic vulnerability. In Australia, for instance, lower levels of financial wellbeing were observed during the early stage of the pandemic ^15^. But in other countries such as Italy, Spain or the United Kingdom, economic concerns was not significantly improved during the pandemic ^16^. Even though it was documented that the pandemic was associated with a sharp increase in mental health problems ^17^ and inequalities generated by school closures, job type and sector of activity ^18,19^, associations between both dimension has been clearly identified. More recently, the cost of living crisis that was fuelled by inflation due to post-COVID-19 global consumer demand, supply chain disruption and soaring energy priced due to the Russian invasion of Ukraine may have had an effect on financial wellbeing as inflation leads to higher poverty levels, greater income inequalities, debt problems and other financial difficulties, all of which are associated with worse physical and mental health ^20^.

The UK was characterized by a large number of policy interventions during the pandemic that somehow contrasts with the lack of intervention during the following cost-of-living crisis^21^. For instance, the COVID-19 job retention scheme (furlough) has contributed to maintaining a large part of the population in employment, minimize unemployment during the pandemic and, consequently, protect workers’ mental health ^22^. Similarly, the Credit Holiday scheme was implemented for borrowers to be able to request a delay in repaying their financial or mortgage debts, resulting in mental health benefits among those who used the scheme ^23^. Yet, whilst these interventions have mitigated the economic costs of the COVID-19 crisis and financial insecurity for (most) people, population mental health was drastically affected. Social isolation ^24^ and home working ^25^ have, for instance, contributed to explain such a trend. Starting in 2022. The UK has experienced the highest surge in prices over the past 30 years, exceeding the increase seen after the 2008 financial crisis and surpassing other comparable countries. Mechanisms that explain poor mental health outcomes are mainly related to insecurity to meet basic needs. This comes after a decade of austerity that led to social security cuts with, for instance, caps on social benefits ^26^ or a reform of the universal tax credit ^27^. In comparison with the pandemic period, the policy response to the cost of living crisis has been seen as minimal with tax rises and cuts to public spending amidst great uncertainty ^28^, leading to a potential public health crisis ^29^.

In such a context and based on previous findings, three hypotheses can be drawn. First, (**hypothesis 1**) the COVID-19 and cost-of-living crisis have generated an economic impact that translated into increased financial hardship and uncertainty that have contributed to higher levels of financial distress within the population. Second, (**hypothesis 2**) the accumulation of financial hardship and uncertainty over time are linked to greater levels of mental health problems in comparison to single events for which mental strains resolve over time. Third, (**hypothesis 3**) financial hardship and financial uncertainty correspond to different concepts that translate into poor mental wellbeing in different ways. To address these three hypotheses, this study takes a parallel approach comparing two cohorts of respondents aged 16 and over in the United Kingdom over two different time periods (2016-2019 & 2019-2022). First, we address how contextual changes have affected rates of financial hardship and financial uncertainty as well as psychological distress across these two different periods in the United Kingdom. Second, we estimate whether the intensity of the relationships between changes in psychological distress and financial hardship and uncertainty trajectories are different across periods.

## Methods

### Data

We use individual panel data from the UK Household Longitudinal Survey (UKHLS), i.e., Understanding Society (USoc), that is a nationally representative dataset of the UK including respondents in England, Wales, Scotland and Northern Ireland. The study uses USoc waves 7 to 13 (the most recent). Wave 7 was collected between January 2015 and May 2018. Wave 8 was collected between January 2016 and May 2019. Wave 9 was collected between January 2017 and May 2019. Wave 10 was collected between January 2018 and May 2020. Wave 11 was collected between January 2019 and May 2021. Wave 12 was collected between January 2020 and May 2022. Wave 13 was collected between January 2021 and May 2023. For each wave, the data collection process spans over 2 years (see <u>supplementary file S.1.</u>). To simplify the reading, wave 7 will be referred to year 2016, wave 8 to year 2017, wave 9 to year 2018, wave 10 to year 2019, wave 11 to year 2020, wave 12 to year 2021 and wave 12 to year 2022. To address how changes in financial hardship and financial wellbeing has affected psychological distress during the recent period, we extracted two 4-years-long cohorts. The first cohort (**cohort 1**) includes wave 7 as the baseline and waves 8, 9 and 10 as follow-up waves. The second cohort (**cohort 2**) includes wave 10 as the baseline and waves 11, 12 and 13 as follow-up waves. Cohort 2 specifically includes waves collected over the recent period with waves 2020 and 2021 collected during the COVID-19 pandemic and wave 2022 collected at the start of the so-called cost-of-living crisis with a pre-pandemic baseline (data collection in wave 10 stopped at the early stage of the pandemic). We include all respondents aged 16 and over at each baseline with a sample 41,855 (28,291 after restriction to complete cases) respondents in cohort 1 and 33,588 (23,246) respondents in cohort 2.

### Outcome

We use a binary version of the General Health Questionnaire composed of 36 items (GHQ-36) ^30^ that converts valid answers to 12 questions of the General Health Questionnaire (GHQ) to a single scale and then summing, giving a scale running from 0 (the least distressed) to 36 (the most distressed). These items include concentration, loss of sleep, playing a useful role, capable of making decisions, constantly under strain, problem overcoming difficulties, enjoy day-to-day activities, ability to face problems, unhappy or depressed, losing confidence, believe worthless and general happiness ^31^. To avoid issues related to the use of binary outcomes in Structural Equation Modelling, we kept the variable linear on its original scale (i.e, from 1 to 36).

### Exposures

We focus on two exposure variables. <u>Financial hardship</u> is derived from a financial wellbeing variable (i.e., how well at managing financially these days?) that is coded over five modalities from 1 (living comfortably) to 5 (finding it very difficult) and is transformed into a binary variable where ‘0’ is attributed to those reporting living comfortably, doing alright or just about getting by and ‘1’ to those reporting finding it quite difficult or very difficult. <u>Financial uncertainty</u> is derived from a variable measuring poor financial prospect for the year ahead that originally contains three categories (better, worse, same) and is recoded as binary, distinguishing those reporting that their situation will be worse (coded ‘1’) from those reporting that their situation will be better similar or similar (coded ‘0’). Financial hardship and uncertainty were collected at each wave. We generated four main variables to address financial trajectories over time for each cohort. First, we generated a baseline financial hardship and uncertainty variable for wave 2016 in cohort 1 and wave 2019 in cohort 2. Second, we generate a subsequent change in financial hardship and uncertainty variable combining information on follow-up trajectories including the following modalities: constant hardship/uncertainty, no hardship/uncertainty, hardship/uncertainty at b (baseline) +1, hardship/uncertainty at b +2, hardship/uncertainty at b +3, hardship/uncertainty at b +1 and b +2, hardship/uncertainty at b +1 +3, hardship/uncertainty at b +2 and b +3, hardship/uncertainty at b +3.

### Control variables and adjustment layers

Modelling is repeated based on four layers of adjustment to address the specific effect of sets of covariates on the relationship between mental health and psychological distress and avoid potential overadjustment including: (a) unadjusted, (b) adjusted for socio-demographic covariates, (c) adjusted for socio-economic covariates and (d) adjusted for the change in employment status. <u>Socio-demographic covariates</u> are collected at baseline and include age, gender (male is the reference category), the highest level of education (higher education degree *versus* no-degree (reference)), the country of residence (distinguishing Scotland, Wales, Northern Ireland, North of England (ref.) and South of England including Greater London) and household composition (couple without child(ren) (ref.), couple with child(ren), single with child(ren) and single) and the presence of one or more chronic health condition (yes, no (ref.)). <u>Socio-economic covariates</u> include baseline information on housing tenure (renting, owning with mortgage or owning without mortgage (=reference)), ability to save money on a regular basis (yes, no (ref.)) and an area-based Index of Material Deprivation ^32^ provided by USoc (IMD) that measures area of residence deprivation across seven domains including income, employment, education, health, crime, access to services and housing environment with a final deprivation score that is the weighted sum of these domains and converted into quintiles (reference: third quintile). IMD is not present in the individual panel and was imputed from the USoc household panel. Saving capacity was not collected at each wave, the reason why we only used baseline (2019) values from cohort 2 and wave 2017 for cohort 1. Finally, the final layer of adjustment includes the employment status of each time points and includes the following categories: employed (ref.), self-employed, student, retired, on sick leave or on maternity/paternity leave.

### Analyses

We first produce descriptive statistics on change in the financial situation within each cohort. We provide the percentages of financial hardship and uncertainty by time points in each cohort for both male and female respondents. Then, we provide the rates of follow-up trajectories by baseline financial status in cohort 1 and cohort 2 for both financial hardship and financial uncertainty. Finally, produce density plots on GHQ-36 distribution across time points.

We then apply parallel analysis on each cohort using a Latent Growth Modelling (LGM) technique within the Structural Equation Modelling (SEM) framework ^33^ to construct a latent curve representing GHQ-36 scores across four time points, including an intercept (at waves 2016 in cohort 1 and 2019 in cohort 2) and a slope tracking subsequent changes (i.e., years 2017-2019 in cohort 1 and 2020-2022 in cohort 2). LGMs are sophisticated statistical tools employed in longitudinal analysis to address latent developmental trajectories underlying observed variables across multiple measurement occasions. These models assume that the observed data result from a combination of systematic growth patterns and random errors. Through decomposition, LGMs estimate latent parameters representing the intercept (initial status) and slope (rate of change) of the growth trajectory, potentially encompassing additional factors such as quadratic trends or group disparities. By elucidating individual variations in growth patterns within a population, LGMs facilitate the examination of predictors or covariates influencing these trajectories and to assess heterogeneity across subgroups. The model is replicated separately for financial hardship and financial uncertainty and a final model include both variables. In each model, baseline financial status is used to explain both the intercept and the slope whilst the slope is also explained by subsequent changes trajectories, as can be seen in figure 1 where ‘i’ and ‘s’ are the intercept and slope and ‘C’ corresponds to the covariates, i.e. the different layers adjustment explaining both the intercept and slope.

**Figure 1.**
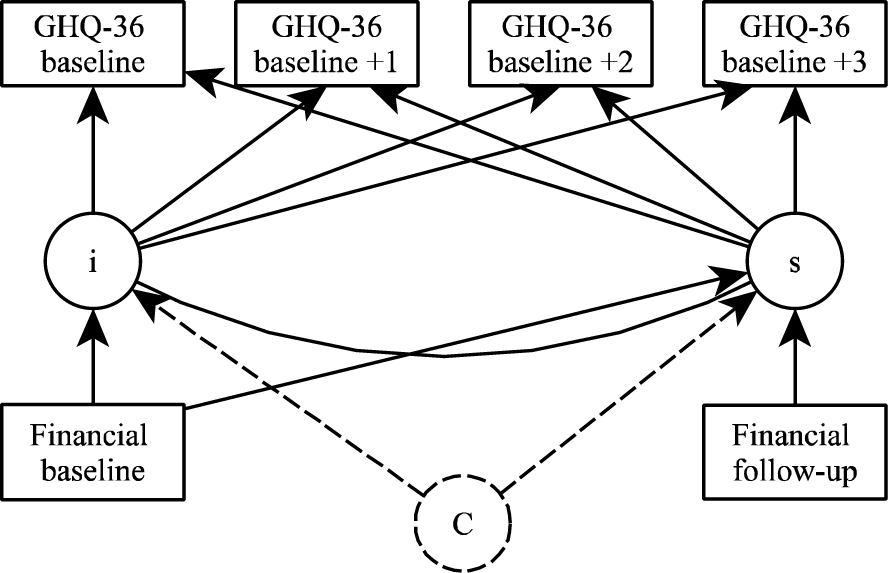
Latent Growth Modelling (LGM) design.

### Weights and missing data

We use complete-case analysis including all baseline cohort 1 and cohort 2 respondents respectively at wave 2016 and 2019. Data are weighted using USoc-provided baseline cross-sectional weight. Attrition over subsequent waves was addressed for both cohorts. We first ran a binary logit model using our baseline exposure, outcome and control variables to explain subsequent attrition with no relevant patterns observed, indicating that, based on our selected set of variables, attrition is more likely to be at random than due to our variables of interest (see <u>supplementary files 2 and 3</u>). We then generate an inverse probability weight by assigning weights to the remaining participants based on the inverse of their probability of being observed at each time point, given their observed characteristics and prior attrition patterns to mitigate potential biases due to attrition and reduce potential selection bias. Models are estimated using both cross-sectional weights and the combination of cross-sectional and inverse probability weights with no major differences observed across estimates.

## Results

### Descriptive statistics

Table 1 shows the percentage of respondents reporting financial hardship and financial uncertainly across waves for the full population by gender including 95 percent confidence intervals. Data are weighted using a combination cross-section baseline weights and inverse probability weights for attrition, explaining why results from 2019 are not exactly similar across cohort (i.e., they represent percentages for different selected populations). We observed slightly higher percentages of financial hardship among female in both cohorts but no clear pattern is observed for financial uncertainty. Percentages of respondents reporting financial hardship are stable across cohorts with between 25.7 and 27.8 percent in cohort 1 and 24.7 and 28.2 percent in cohort 2. By contrast, financial uncertainty is between 10-13 percent for both cohorts except in 2022 for cohort 2 where it reaches 22.9 percent, i.e., 10.6 percentage points more than at the same period in cohort 1. At population level, the COVID-19 pandemic and the subsequent cost-of-living crisis are not associated with a substantial increase in financial hardship but the 2022 crisis translates into much higher rates of financial uncertainty.

**Table 1.** Percentages of financial hardship and financial uncertainly across waves 2019 to 2022 and 95%CI.

| **Cohort 1 (2016-2019)** |  |  |  |  |
| --- | --- | --- | --- | --- |
|  | 2016 | 2017 | 2018 | 2019 |
|  | **Financial hardship** |  |  |  |
| Female | 26.8<br>(25.7-27.9) | 26.6<br>(25.5-27.7) | 29.5<br>(28.3-30.6) | 29.0<br>(27.9-30.0) |
| Male | 25.5<br>(24.4-26.7) | 25.5<br>(24.3-26.7) | 27.0<br>(25.8-28.3) | 26.6<br>(25.4-27.8) |
| Total | 25.7 | 25.7 | 27.3 | 27.8 |
|  | **Financial uncertainty** |  |  |  |
| Female | 10.9<br>(10.2-10.7) | 12.8<br>(11.9-13.6) | 13.0<br>(12.2-13.9) | 12.5<br>(11.7-13.3) |
| Male | 10.3<br>(9.49-11.2) | 11.9<br>(11.0-12.9) | 13.4<br>(12.4-14.3) | 12.1<br>(11.2-13.0) |
| Total | 10.7 | 12.3 | 13.3 | 12.3 |

| **Cohort 2 (2019-2022)** |  |  |  |  |
| --- | --- | --- | --- | --- |
|  | 2019 | 2020 | 2021 | 2022 |
|  | **Financial hardship** |  |  |  |
| Female | 29.3<br>(28.2 – 30.2) | 28.1<br>(27.0 – 29.2) | 25.8<br>(24.6 – 26.9) | 26.5<br>(25.3 – 27.7) |
| Male | 27.3<br>(26.2-28.5) | 27.0<br>(25.7 – 28.2) | 24.2<br>(22.9 -25.4) | 25.3<br>(23.9 – 26.6) |
|  | 28.2 | 27.4 | 24.7 | 26.1 |
|  | **Financial uncertainty** |  |  |  |
| Female | 12.5<br>(11.7 – 13.2) | 13.6<br>(12.7 – 14.4) | 13.7<br>(12.8 – 14.6) | 22.9<br>(21.7 – 24.0) |
| Male | 11.7<br>(10.9 – 12.6) | 14.1<br>(13.2 – 15.1) | 14.4<br>(13.4 – 15.4) | 22.8<br>(21.5 – 24.1) |
|  | 12.1 | 14.8 | 14.1 | 22.9 |

Note: data are weighted using a mixture of baseline cross-sectional weight and inverse probability weight to adjust for subsequent attrition.

The overall distribution of financial hardship and uncertainty across the four time points does not translate well individuals’ trajectories. Table 2 shows the distribution of financial hardship and uncertainly trajectories based on baseline (2016 and 2019) financial hardship and wellbeing. Looking at respondents reporting baseline financial hardship, we observe that 42.8 (95%CI= 40.4; 44.7) and 41.4 (95%CI= 39.4; 43.5) experienced constant hardship at follow-up, respectively in cohort 1 and 2. Among those who reported financial wellbeing, respectively 74.2 (95%CI=73.2; 75.2) and 77.9 (95%CI= 76.8; 78.9) percent reported constant wellbeing in cohort 1 and 2. Looking at those who reported baseline financial uncertainty, the percentage of respondents reporting constant uncertainty at follow-up is 14 percent (95%CI= 12.0; 16.1) in cohort 1 and 19.3 percent (95%CI= 17.0; 21.7) in cohort 2. When looking within the same group those reporting financial uncertainty three years late (i.e. in 2016 for cohort 1 and 2022 for cohort 2), percentages are respectively 6.9 (95%CI=5.4; 8.5) and 11.6 (95%CI= 10.0; 13.9). The same pattern is observed for those who reported no financial uncertainty at baseline as the percentage of respondents who reported no uncertainty at all over subsequent waves is 78.2 percent (95%CI= 77.4; 79.1) in cohort 1 and 69.8 percent (95%CI= 68.7; 70.8) in cohort 2, a difference of about 10 percentage points that is mainly due to increased financial uncertainty in 2022.

**Table 2.** Percentages of financial hardship and financial uncertainty trajectories in cohorts 1 (2016-2019) and 2 (2019-2022) by baseline financial hardship and uncertainty and 95%CI.

| **Cohort 1 (2016-2019)** |  |  |  |  |  |  |  |  |
| --- | --- | --- | --- | --- | --- | --- | --- | --- |
|  | Financial hardship trajectories |  |  |  |  |  |  |  |
|  | Constant hardship | Constant wellbeing | 2017 | 2018 | 2017-18 | 2019 | 2018 and 2019 | 2017 and 2019 |
| Baseline financial hardship (25.7%) | 42.8<br>(40.9-44.7) | 14.9<br>(13.5-16.3) | 7.6<br>(6.2-8.7) | 5.0<br>(4.2-5.9) | 9.0<br>(7.89-10.1) | 5.1<br>(4.3-6.0) | 8.8<br>(7.7-9.9) | 6.7<br>(5.7-7.6) |
| Baseline financial wellbeing (74.3%) | 4.1<br>(3.7-4.5) | 74.2<br>(73.2-75.2) | 4.0<br>(3.6-4.4) | 4.6<br>(4.2-5.1) | 2.1<br>(1.8-2.4) | 5.2<br>(4.7-5.7) | 4.1<br>(3.6-4.5) | 1.7<br>(1.4-2.0) |
|  | Financial uncertainty trajectories |  |  |  |  |  |  |  |
|  | Constant uncertainty | Constant non uncertainty | 2017 | 2018 | 2017-18 | 2019 | 2018 and 2019 | 2017 and 2019 |
| Baseline financial uncertainty (10.7%) | 14.0<br>(12.0-16.1) | 41.9<br>(39.0-44.9) | 12.2<br>(10.2-14.1) | 8.25<br>(6.6-9.9) | 7.0<br>(5.5-8.6) | 6.9<br>(5.4-8.5) | 4.5<br>(3.3-5.8) | 5.1<br>(3.8-6.4) |
| Baseline no financial uncertainty (89.3%) | 1.4<br>(1.2-1.7) | 78.2<br>(77.4-79.1) | 4.7<br>(4.3-5.2) | 5.4<br>(4.9-5.9) | 1.6<br>(1.3-1.8) | 5.0<br>(4.5-5.4) | 2.3<br>(2.0-2.6) | 1.4<br>(1.2-1.6) |

| **Cohort 2 (2019-2022)** |  |  |  |  |  |  |  |  |
| --- | --- | --- | --- | --- | --- | --- | --- | --- |
|  | Financial hardship trajectories |  |  |  |  |  |  |  |
|  | Constant hardship | Constant wellbeing | 2020 | 2021 | 2020-21 | 2022 | 2021 and 2022 | 2020 and 2022 |
| Baseline financial hardship (28.2%) | 41.4<br>(39.4-43.5) | 16.3<br>(14.7-17.9) | 10.5<br>(9.2-12.0) | 3.5<br>(2.7-4.29) | 11.8<br>(10.4-13.01) | 5.4<br>(4.4-6.4) | 5.05<br>(4.1-6.0) | 6.1<br>(5.0-7.1) |
| Baseline financial wellbeing (71.8%) | 2.9<br>(2.5-3.4) | 77.9<br>(76.8-78.9) | 4.6<br>(4.0-5.1) | 3.4<br>(2.9-3.8) | 1.5<br>(1.2-1.8) | 5.2<br>(4.7-5.7) | 2.6<br>(2.2-3.0) | 1.87<br>(1.5-2.2) |
|  | Financial uncertainty trajectories |  |  |  |  |  |  |  |
|  | Constant uncertainty | Constant non uncertainty | 2020 | 2021 | 2020-21 | 2022 | 2021 and 2022 | 2020 and 2022 |
| Baseline financial uncertainty (12.1%) | 19.3<br>(17.0-21.7) | 30.6<br>(27.9-33.4) | 9.8<br>(8.0-11.5) | 6.1<br>(4.7-7.5) | 6.1<br>(4.7-4.5) | 11.9<br>(10.0-13.9) | 8.0<br>(6.4-9.6) | 8.16<br>(6.5-9.8) |
| Baseline no financial uncertainty (87.9%) | 1.9<br>(1.6-2.2) | 69.8<br>(68.7-70.8) | 4.8<br>(4.3-5.3) | 4.4<br>(4.0-4.9) | 1.18<br>(0.9-1.4) | 12.4<br>(11.6-13.1) | 3.0<br>(2.6-3.4) | 2.4<br>(2.0-2.8) |

Note: data are weighted using a mixture of baseline cross-sectional weight and inverse probability weight to adjust for subsequent attrition.

Density plots of GHQ-36 distribution is shown in <u>supplementary file 4</u> for both cohort 1 (figure S.3.1) and cohort 2 (figure S.3.2). The baseline mean is 10.8 for cohort 1 and 11.4 for cohort 2. GHQ-36 means are 11, 11.1 and 11.3 over the subsequent waves in cohort 1 and 11.7, 11.9 and 11.7 in cohort 2. These results corresponds to what is observed within the full population. Figure S.4.3. shows the percentage of respondents aged 16 and over reporting GHQ-caseness (i.e., those reporting a GHQ-36 equal or above 9) in the UK from 1991 to 2021 based on USoc data. GHS-caseness has indeed constantly increased since 2015, finding that we find using a cohort approach as well.

### Latent growth modelling

Results from the LGM are shown in <u>supplementary files 5 and 6</u>, respectively for cohort 1 and cohort 2. They include the four layers of adjustment as well as cross-sectionally weighed estimates and the combination of cross-sectional and inverse probability weights to adjust for attrition. To ease the interpretation of the intercepts and slopes from the LGM, we have plotted the baseline and follow-up differences in GHQ-36 by both financial hardship (figure 1) and financial uncertainty (figure 2) for both cohort 1 (1.a. and 2.a.) and cohort 2 (1.b. and 2.b.). We only represent the most comment trajectories, other estimates and 95% confidence intervals are in supplementary files.

**Figure 2.**
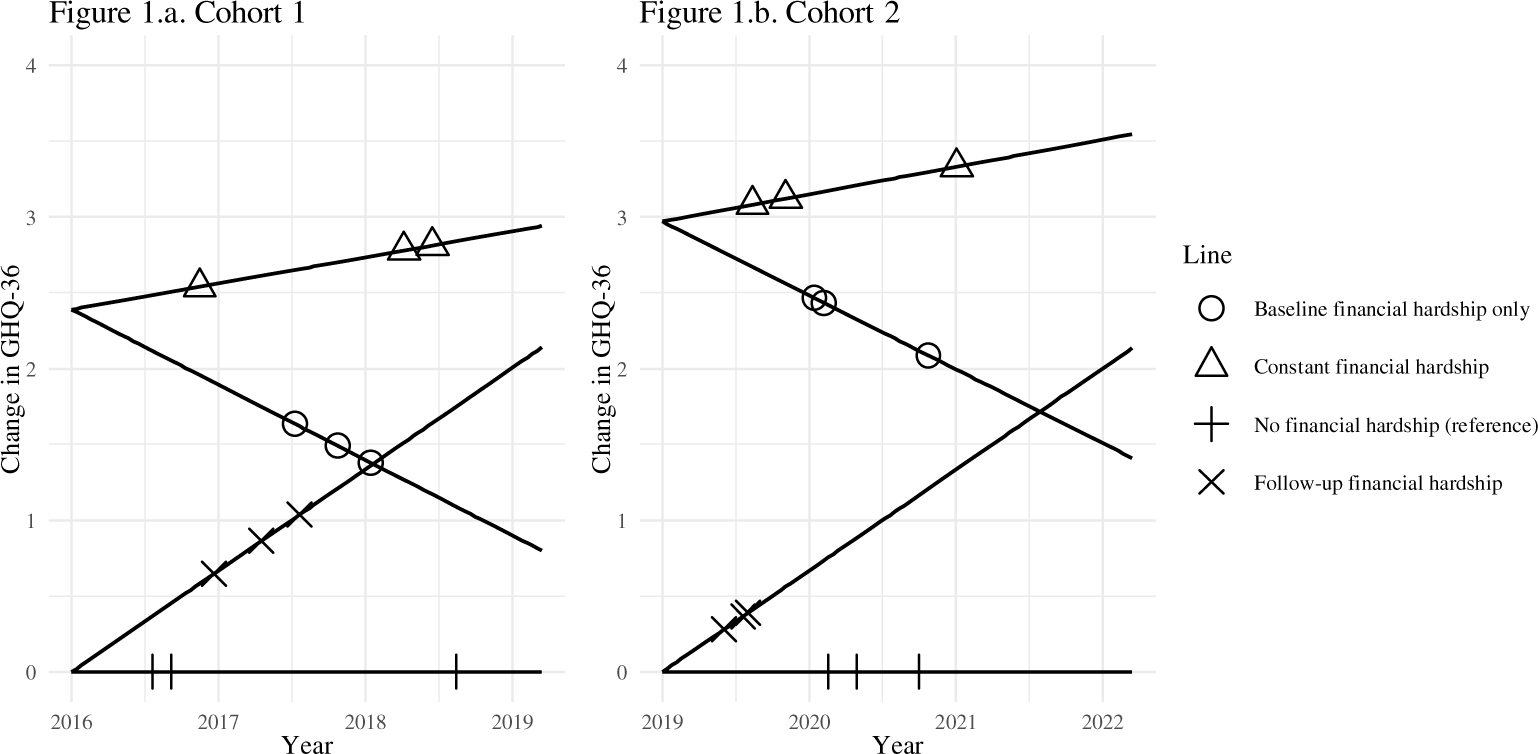
Differences in mental health (GHQ-36) scores by baseline (intercept) and follow-up (slope) financial wellbeing, fully adjusted model including cross-sectional and inverse probability of attrition weights for cohort 1 and cohort 2.

Figure 2 shows that those reporting financial hardship at baseline in 2019 (cohort 2) have experienced a greater level of psychological distress – although not significant – during the pandemic and subsequent cost-of-living crisis with an intercept of 2.97 (95%CI= 2.65; 3.29) in comparison to the 2016 cohort (intercept: 2.39, 95%CI= 2.11; 2.67). Follow-up changes in financial situation show little difference between cohort 1 and cohort 2 with a slope of respectively -0.66 (95%CI: -0.78; -0.55) and -0.65 (95%CI= -0.79, -0.51) for those who experience no financial hardship in the follow-up waves and, similarly, 0.89 (95%CI= 0.76; 1.02 and) 0.89 (95%CI= 0.73; 1.05) for those who experienced constant financial hardship throughout the following waves.

The pattern is slightly different for financial uncertainty as both the intercepts and slopes are different across cohorts as shown in figure 3. In cohort 1, the intercept was 1.74 (95%CI= 1.38, 2.10) against 2.12 (95%CI= 1.75; 2.49) for cohort 2, indicating higher GHQ-36 scores across the pandemic and the cost-of-living crisis for those who have reported financial uncertainty at baseline, although this is not statistically significant as the confidence intervals overlap. Not reporting financial uncertainty at follow-up is associated with a similar slope in cohort 1 (-0.327, (95%CI= -0.45, -0.20)) and cohort 2 (-0.339 (95%CI=-0.47; -0.18)) but reporting constant financial uncertainty at follow-up is associated with higher GHQ-36 scores for cohort 2 (1.037, 95%CI= 0.82; 1.25) in comparison to cohort 1 (0.955, 95%CI= 0.74; 1.17), but this, again is not significant.

**Figure 3.**
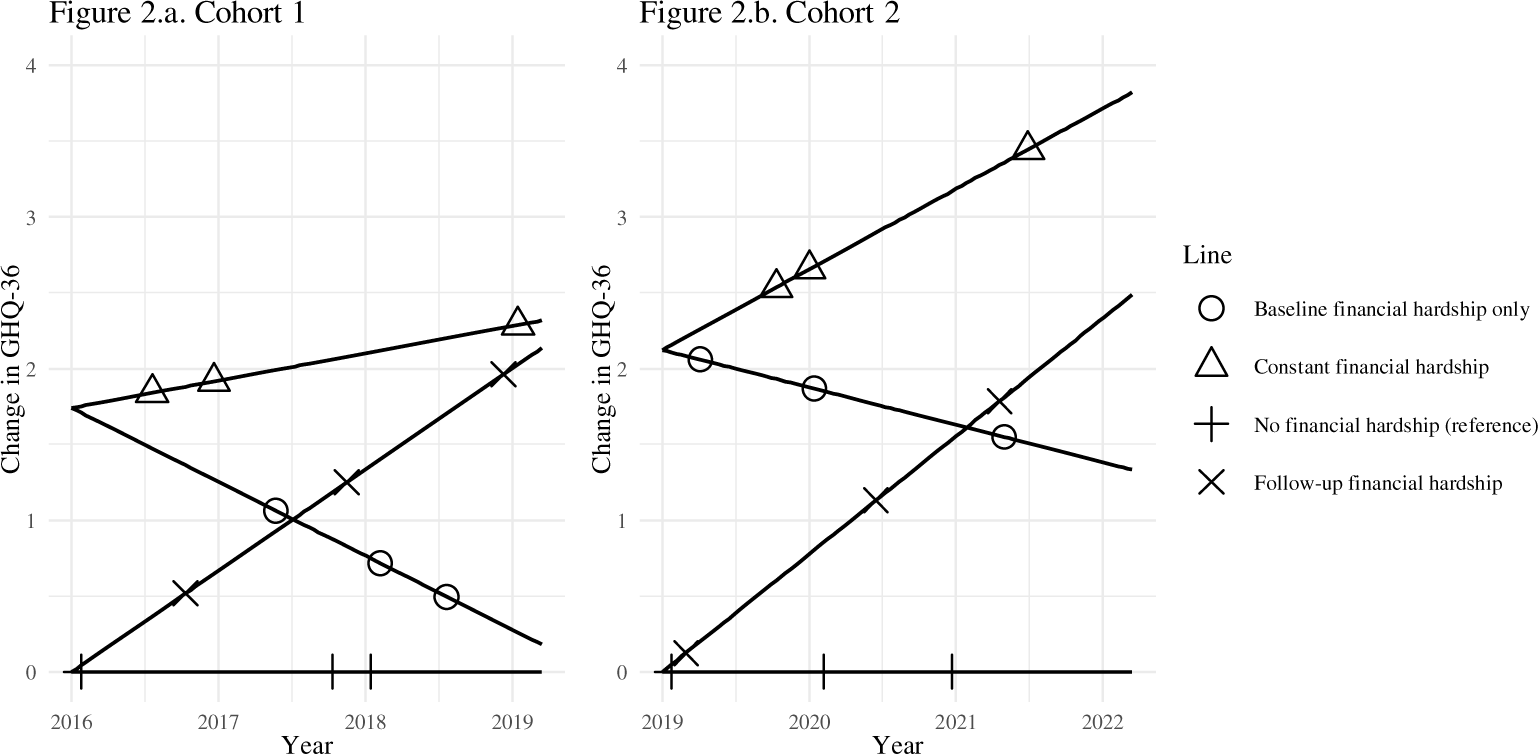
Differences in mental health (GHQ-36) scores by baseline (intercept) and follow-up (slope) financial uncertainty, fully adjusted model including cross-sectional and inverse probability of attrition weights for cohort 1 and cohort 2.

Focusing on baseline + 3, i.e. financial hardship and financial uncertainty in 2019 for cohort 1 and 2022 for cohort 2, estimates for the slope (independently of baseline values) are similar. Financial hardship is associated with a slope of 0.73 (95%CI= 0.56; 0.91) in cohort 1 and 0.73 (95%CI= 0.54; 0.92) in cohort 2 whilst financial uncertainty is associated with a slope of 0.51 (95%CI= 0.36; 0.67) in cohort 1 and 0.47 (95%CI= 0.36; 0.59) in cohort 2. For financial uncertainty (that has drastically increased in baseline + 3 for cohort 2), this corresponds to a difference of 1.53 units (=0.51*3 years) in cohort 1 and 1.41 (=0.47*3 years) in cohort 2 on the GHQ-36 scale.

The same can be found when looking at financial hardship and uncertainty in years 2017-2018 in cohort 1 and years 2020-2021 (during the COVID-19 pandemic) in cohort 2 as estimates are very similar.

What these results show is that, even though small differences can be observed across cohorts, they are not statistically significant meaning that associations between financial hardship and uncertainty trajectories and psychological distress as somehow stable over time.

Financial hardship and, to a lesser extent, financial uncertainty explain psychological distress but the nature of these relationships remained unchanged.

Looking at the different levels of adjustment, we observe that coefficients are higher in the unadjusted model and that each level of adjustment slightly reduces their intensity. For instance, the intercept of financial hardship in cohort 1 (fully weighted model) is 3.08 (95%CI= 2.80; 3.36) in the unadjusted model, 2.93 (95%CI= 2.65; 3.21) in the model adjusting for demographic characteristics (including gender), 2.68 (95%CI= 2.39; 2.96) in the model adjusting for socio-demographic characteristics and, finally, 2.38 (95%CI= 2.11; 2.67) in the model controlling for baseline employment status. However, no adjustment level changes the nature of the relationship that is observed for the different exposures of interest.

We also ran an additional model including both financial hardship and uncertainty (fully adjusted only) to estimate the specific relationship between each exposure and GHQ-36. Estimates for baseline and slope exposures confirm what was observed before, i.e. that financial hardship has a stronger effect on GHQ-36 compared to financial uncertainty but the effect of financial uncertainty is not null. For instance, in the fully weighted model, baseline financial hardship is associated with higher GHQ-36 of 2.33 units (95%CI= 2.05; 2.62) in cohort 1 and 2.83 units (95%CI= 2.51; 3.16) in cohort 2 whilst the coefficients for financial uncertainty are respectively 1.59 (95%CI= 1.24; 1.95) and 1.68 (95%CI= 1.31; 2.05). Adjusting for both variables at the same time is of interest because it shows that the association between financial hardship and GHQ-36 is somehow independent of financial uncertainty and, similarly, financial uncertainty is associated with GHQ-39 independently of financial hardship.

## Limitations

This study is one of the first to provide empirical evidence on the recent cost-of-living crisis in the UK and its implication for mental health. However, it is not without limitations.

A first limitation is about data collection. Recent data are only available until 2023 and collected between January 2021 and May 2023, which corresponds to the very early stage of the cost-of-living crisis. This might explain why financial uncertainty appears to be increased in 2022 but not financial hardship. It might be expected that next waves will show a potential increase in financial hardship in response to inflation. This should be addressed further when available.

A second limitation is about the covariates included in the models. We have used several layers of adjustment to avoid over adjusting the model and observed little variations the levels (the financial hardship and uncertainty coefficients are only slightly reduced) but some limitations should be mentioned. First, data on housing tenure are not replicated yearly and had to be used as fixed. Because of this, they were collected at baseline in cohort 2 and in the second wave of cohort 1. Second, data on IMD was imputed from the USoc household dataset to the individual dataset and only baseline information is used. Third, change in employment is measured across different waves but does not contain information on furlough (the COVID-19 job retention scheme) because furlough was implemented over a limited number of months and for some specific types of occupation. Furlough workers are therefore included within the employed category. This does not raise a major methodological concern because furlough workers experienced a much lower declined in mental health compared to newly unemployed respondents during the pandemic ^22^.

A third limitation is about attrition. Maximum attrition between the baseline and the last selected waves is 32 percent in cohort 1 and 30.8 percent in cohort 2. We have checked attrition patterns using binary logit models to address associations between our set of outcome, exposures and control variables and follow-up attrition with no evidence of strong associations meaning that baseline mental health, financial status or employment do not substantially explain subsequent drop out. We have used inverse probability weight using the same variable to correct our sample for attrition assuming a missing at random pattern. Although some unmeasured cofounders might explain attrition patterns, similar results found for cohort 1 and cohort 2 are reassuring.

Finally, a fourth limitation is about the use of the GHQ-36 scale. We selected this variable for two reasons. Firstly, because binary outcomes (e.g., GHQ-caseness ^34^) are not easy to use within the structural equation modelling (SEM) framework with coefficients somehow hard to interpret. Secondly because GHQ-36 contains a maximum amount of information and results are very similar to those obtained with restricted measures such a GHQ-12 ^31^.

## Discussion

Addressing the potential impact of the COVID-19 pandemic and the subsequent cost of living crisis on financial hardship and uncertainty (**hypothesis 1**), the study provides some nuances. We demonstrate that the COVID-19 pandemic was not associated with a drastic increase in financial hardship nor was it associated with a profound change in financial uncertainty among the UK population. In some way, the implementation of protective schemes such as the COVID-19 job retention scheme ^22^ and the credit holiday ^23^ might have contributed to protecting the UK population against greater financial distress. This is not true when it comes to the subsequent cost-of-living crisis. 2022 data clearly indicate a 10 percentage points increase in financial uncertainty within the 16+ population compared to previous times. Associations between poor mental health and financial hardship and uncertainty are – although with some tiny variations over time – constant: the onset of financial hardship and uncertainty is associated with increased psychological distress. An increase of 10 percentage points in financial uncertainty therefore leads to greater psychological distress among the population. However, what happened during the cost-of-living crisis should not hide a more stable phenomenon: in the UK, more than 25 percent of the population experienced financial hardship and this is independent of contextual factors such as the pandemic or the cost of living crisis. These people report an average psychological distress that is higher by about 2.3 units compared to the rest of the population, corresponding to a difference of 6.3 percentage points. Our study also shows that the accumulation of financial hardship and financial uncertainty increases psychological distress. For instance, we estimate that 28.2 percent of the 2019-2022 cohort experienced financial hardship at baseline. Among those, 41.4 percent experienced constant financial hardship. These people have experienced GHQ-36 increased by 3.5 units in comparison to those who never experienced financial hardship.

Addressing the potential accumulation of financial strains over time and their relationship with poor mental health (**hypothesis 2**), we estimated that contextual events offer little explanations on individual mental health trajectories. Baseline hardship and uncertainty are associated with greater degrees of psychological distress but follow-up financial wellbeing is associated with a sharp decline in psychological distress. Although the cohorts only focus on a 4-year period, it can be extrapolated that it would take between five and six years for respondents’ mental health to fully recover after financial hardship or uncertainty ends, confirming what has been found in previous studies ^4,5^. We do not find any difference between cohort 1 and 2 on how accumulation of financial hardship over the life course would translate into greater mental health issues in recent years. However, both descriptive statistics and the models’ intercepts indicate that psychological distress has sharply increased in the UK since the mid 2010s, indicating a broader structural context that needs to be addressed further.

Finally, the study demonstrates that financial hardship and financial uncertainty are associated with higher levels of psychological distress, independent on baseline mental health, indicating a possible causal relationship. However, the extent to which these variables affect mental health varies (**hypothesis 3**). The impact of financial hardship on psychological distress is approximately twice as pronounced as that of financial uncertainty. However, as mentioned earlier, this does not imply that uncertainty is insignificant; it actually plays a key role in explaining psychological distress discrepancies across the population.

## Data Availability

The study uses Understanding Society data that can be accessed through the UK data archive portal: https://www.data-archive.ac.uk.

## Funding

JW is the PI of the Horizon Europe ERC Starting Grant UHealth (Trade Unions and Workers’ Health), the PI of the “Negotiating Health: addressing the role of trade unions in explaining workers’ physical and mental health” (NegHealth) Incentive Grant for Scientific Research funded by the Belgian National Scientific fund (FNRS) and the PI of the “Trade unions, job satisfaction and workers’ health: an international comparison of panel data” Joint Usage and Research Center Program at the Institute of Economic Research, Hitotsubashi University, Japan.

## Data access and ethical statement

The University of Essex Ethics Committee has approved all data collection on Understanding Society main study and innovation panel waves. Participant consent was given during data collection. The overall mechanism for gaining consent for participation in Understanding Society is oral. Participants are sent details about the study in advance letters, information leaflets and are given information by interviewers if taking part in a face-to-face or telephone interview. USoc data can be accessed through the UK data archive portal: https://www.data-archive.ac.uk.

## Conflict of interest

The authors report no conflict of interest. JW is a member of the Belgian Health Data Agency (HDA) user committee.

## Notes

### Competing Interest Statement

The authors report no conflict of interest. Jacques Wes is a member of the Belgian Health Data Agency (HDA) user committee.

### Summary of Updates

This version of the manuscript has been revised to update the following: 1/ Develop a LGM framework that compares two Understanding Society cohorts in capture potential changes in associations between financial hardship and uncertainty and psychological distress within two different contexts (pre-Covid and during the COVID-19 pandemic and the cost of living crisis). 2/ Use GHQ-36 as a linear variable and not on a binary basis. 3/ Use the two cohorts to provide descriptives statistics on financial hardship and uncertainty in 2016-2019 (cohort 1) and 2019-2022 (cohort 2). 4/ Move to the appendix the longer descriptive trends in GHQ-caseeness, financial hardship and financial uncertainty that at the start of the result section.

