## Supplementary files S.1. to S.6. for "Examining the relationships between psychological distress and financial hardship and uncertainty during the COVID-19 pandemic and the subsequent cost-of-living crisis in the United Kingdom"

### Supplementary file S.1. Data collection periods, waves and selected cohorts

<table>
<tr><td rowspan="7">Data collection</td><td colspan="2">Jan 2015 - May 2017</td><td></td><td></td><td></td><td></td><td></td><td></td></tr>
<tr><td></td><td colspan="2">Jan 2016 - May 2018</td><td></td><td></td><td></td><td></td><td></td></tr>
<tr><td></td><td></td><td colspan="2">Jan 2017 - May 2019</td><td></td><td></td><td></td><td></td></tr>
<tr><td></td><td></td><td></td><td colspan="2">Jan 2018 - May 2020</td><td></td><td></td><td></td></tr>
<tr><td></td><td></td><td></td><td></td><td colspan="2">Jan 2019 - May 2021</td><td></td><td></td></tr>
<tr><td></td><td></td><td></td><td></td><td></td><td colspan="2">Jan 2020 - May 2022</td><td></td></tr>
<tr><td></td><td></td><td></td><td></td><td></td><td></td><td colspan="2">Jan 2021 - May 2023</td></tr>
<tr><td rowspan="2">Waves</td><td colspan="2">Wave g</td><td colspan="2">Wave i</td><td colspan="2">Wave j</td><td colspan="2">Wave m</td></tr>
<tr><td></td><td colspan="2">Wave h</td><td colspan="2">Wave j</td><td colspan="2">Wave l</td><td></td></tr>
<tr><td rowspan="2">Cohorts</td><td colspan="5">Cohort 1</td><td></td><td></td><td></td></tr>
<tr><td></td><td></td><td></td><td colspan="5">Cohort 2</td></tr>
</table>

### Supplementary file S2. Attrition pattern in cohort 1

Baseline sample: 41.855
Attrition in 2020: 8,466
Attrition in 2021: 11,897
Attrition in 2022: 13,564

**Ordered logit regression of attrition in subsequent waves by baseline variables**

| | Estimate | Std. Error | t value | Pr(>\|t\|) |
|---|---|---|---|---|
| **(Intercept)** | 0.250 | 0.016 | 15.331 | 0.000 |
| **FinancialHardship2016** | 0.014 | 0.006 | 2.370 | 0.018 |
| **FinancialUncertainty2016** | -0.012 | 0.008 | -1.506 | 0.132 |
| **ghq** | 0.001 | 0.000 | 1.822 | 0.068 |
| **sex_female** | -0.002 | 0.005 | -0.357 | 0.721 |
| **age** | -0.003 | 0.000 | -11.785 | 0.000 |
| **isced** | -0.009 | 0.007 | -1.178 | 0.239 |
| **country_LondonandSouthEngland** | 0.003 | 0.005 | 0.573 | 0.567 |
| **coupleandchildren** | 0.012 | 0.007 | 1.749 | 0.080 |
| **singleandchildren** | 0.029 | 0.011 | 2.578 | 0.010 |
| **single** | 0.014 | 0.007 | 2.120 | 0.034 |
| **own_mortgage** | -0.007 | 0.007 | -1.018 | 0.309 |
| **own_rent** | 0.011 | 0.008 | 1.490 | 0.136 |
| **save** | -0.022 | 0.005 | -4.337 | 0.000 |
| **IMD_1** | 0.027 | 0.007 | 3.870 | 0.000 |
| **IMD_2** | 0.001 | 0.007 | 0.143 | 0.886 |
| **IMD_5** | -0.002 | 0.006 | -0.383 | 0.702 |
| **empl_16_unemployed** | 0.028 | 0.014 | 2.038 | 0.042 |
| **empl_16_student** | 0.021 | 0.012 | 1.772 | 0.076 |
| **empl_16_retired** | 0.004 | 0.009 | 0.518 | 0.604 |
| **empl_16_self** | 0.002 | 0.009 | 0.256 | 0.798 |
| **empl_16_sick** | 0.005 | 0.017 | 0.324 | 0.746 |
| **empl_16_maternity** | 0.003 | 0.011 | 0.237 | 0.813 |

(Dispersion parameter for gaussian family taken to be 0.1020826)
Null deviance: 1887.4 on 17859 degrees of freedom
Residual deviance: 1820.8 on 17837 degrees of freedom

### Supplementary file S.3. Attrition pattern in cohort 2

Baseline sample: 33.588
Attrition in 2020: 5,953
Attrition in 2021: 8,649
Attrition in 2022: 10,342

**Ordered logit regression of attrition in subsequent waves by baseline variables**

| | Estimate | Std. Error | t value | Pr(>\|t\|) |
|---|---|---|---|---|
| **(Intercept)** | 0.462 | 0.020 | 23.513 | 0.000 |
| **FinancialHardship2019** | 0.004 | 0.008 | 0.552 | 0.581 |
| **FinancialUncertainty2019** | -0.048 | 0.010 | -4.832 | 0.000 |
| **ghq** | 0.002 | 0.001 | 3.181 | 0.001 |
| **sex_female** | -0.031 | 0.007 | -4.644 | 0.000 |
| **age** | -0.003 | 0.000 | -11.355 | 0.000 |
| **isced** | -0.017 | 0.010 | -1.701 | 0.089 |
| **country_LondonandSouthEngland** | -0.002 | 0.007 | -0.301 | 0.764 |
| **coupleandchildren** | 0.048 | 0.009 | 5.617 | 0.000 |
| **singleandchildren** | 0.117 | 0.014 | 8.260 | 0.000 |
| **single** | 0.042 | 0.009 | 4.763 | 0.000 |
| **own_mortgage** | 0.009 | 0.009 | 0.964 | 0.335 |
| **own_rent** | 0.093 | 0.010 | 9.431 | 0.000 |
| **save** | -0.066 | 0.007 | -9.539 | 0.000 |
| **IMD_1** | 0.060 | 0.009 | 6.495 | 0.000 |
| **IMD_2** | 0.027 | 0.009 | 2.999 | 0.003 |
| **IMD_5** | -0.046 | 0.009 | -5.249 | 0.000 |
| **empl_unemployed** | 0.075 | 0.018 | 4.210 | 0.000 |
| **empl_student** | 0.070 | 0.015 | 4.608 | 0.000 |
| **empl_retired** | 0.078 | 0.012 | 6.607 | 0.000 |
| **empl_self** | 0.049 | 0.012 | 4.022 | 0.000 |
| **empl_sick** | 0.018 | 0.020 | 0.862 | 0.388 |
| **empl_maternity** | 0.048 | 0.017 | 2.917 | 0.004 |

(Dispersion parameter for gaussian family taken to be 0.2167649)
Null deviance: 4971.5 on 21482 degrees of freedom
Residual deviance: 4651.8 on 21460 degrees of freedom

### Supplementary file S.4. GHQ-36, density plots and cross-sectional change

#### Figure S.4.1. GHQ-36 density plot by year in cohort 1 (including cross-sectional and inverse probability weights)

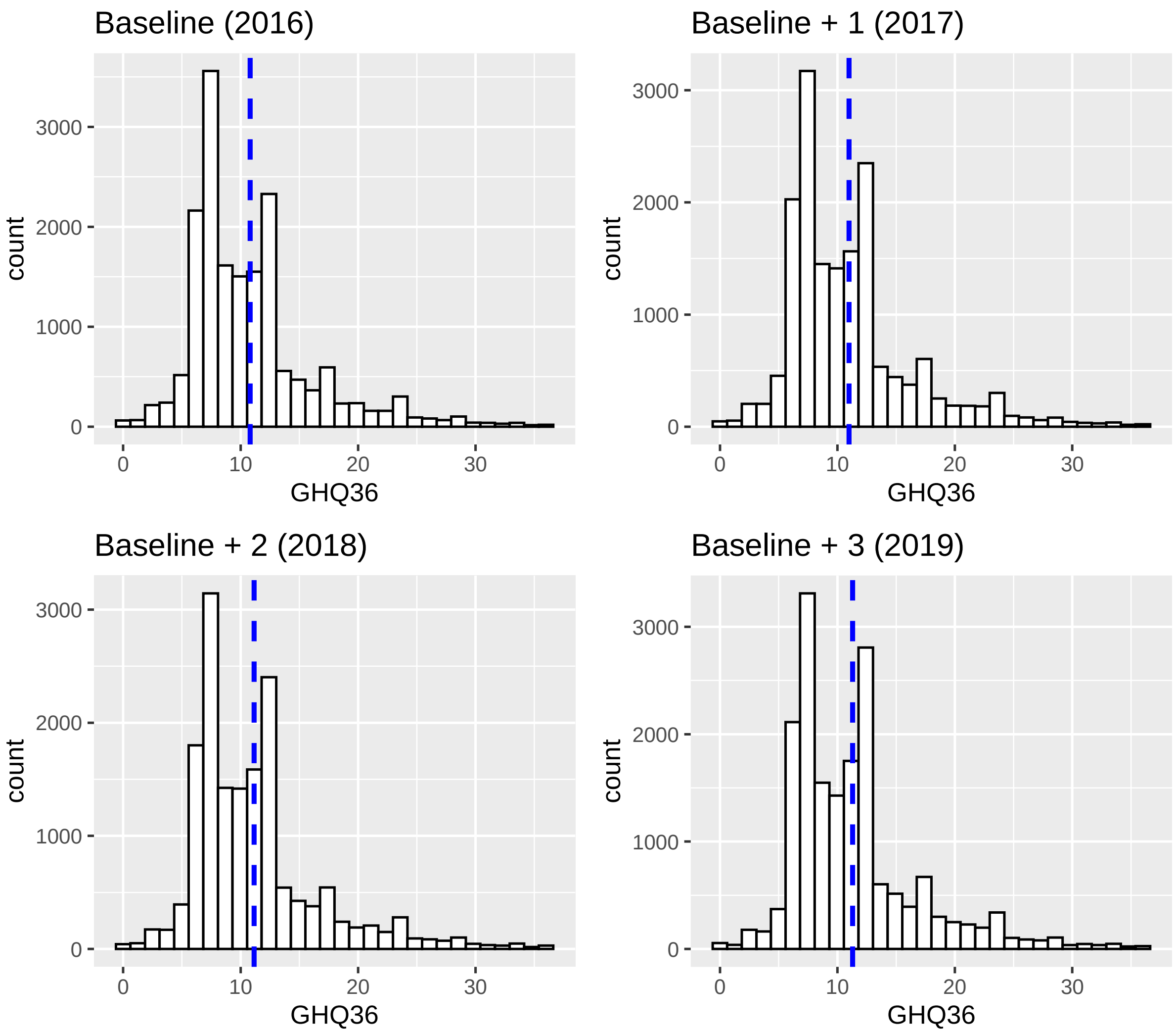

| time | GHQ-36 means |
|---|---|
| 1 2016. | 10.80600 |
| 2 2017 | 10.99273 |
| 3 2018 | 11.14514 |
| 4 2019 | 11.28897 |

#### Figure S.4.2. GHQ-36 density plot by year in cohort 2 (including cross-sectional and inverse probability weights)

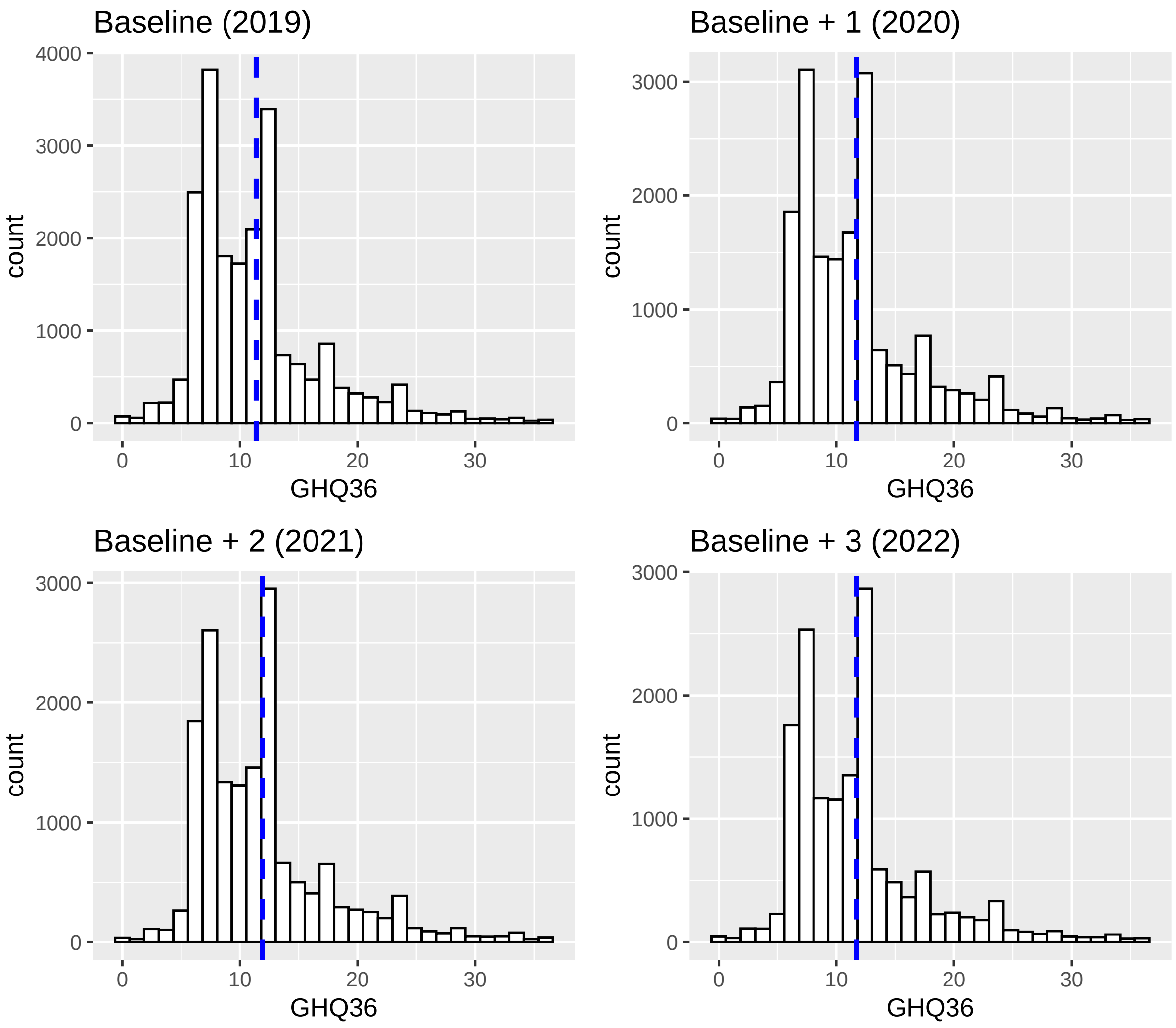

| time | GHQ-36 means |
|---|---|
| 2019 | 11.37476 |
| 2020 | 11.69447 |
| 2021 | 11.88569 |
| 2022 | 11.68791 |

Figure S.4.3. Percentages of GHQ caseness based on GHQ-36 by year), financial uncertainty, financial hardship and consumer price inflation (z-axis) from 1991 to 2022

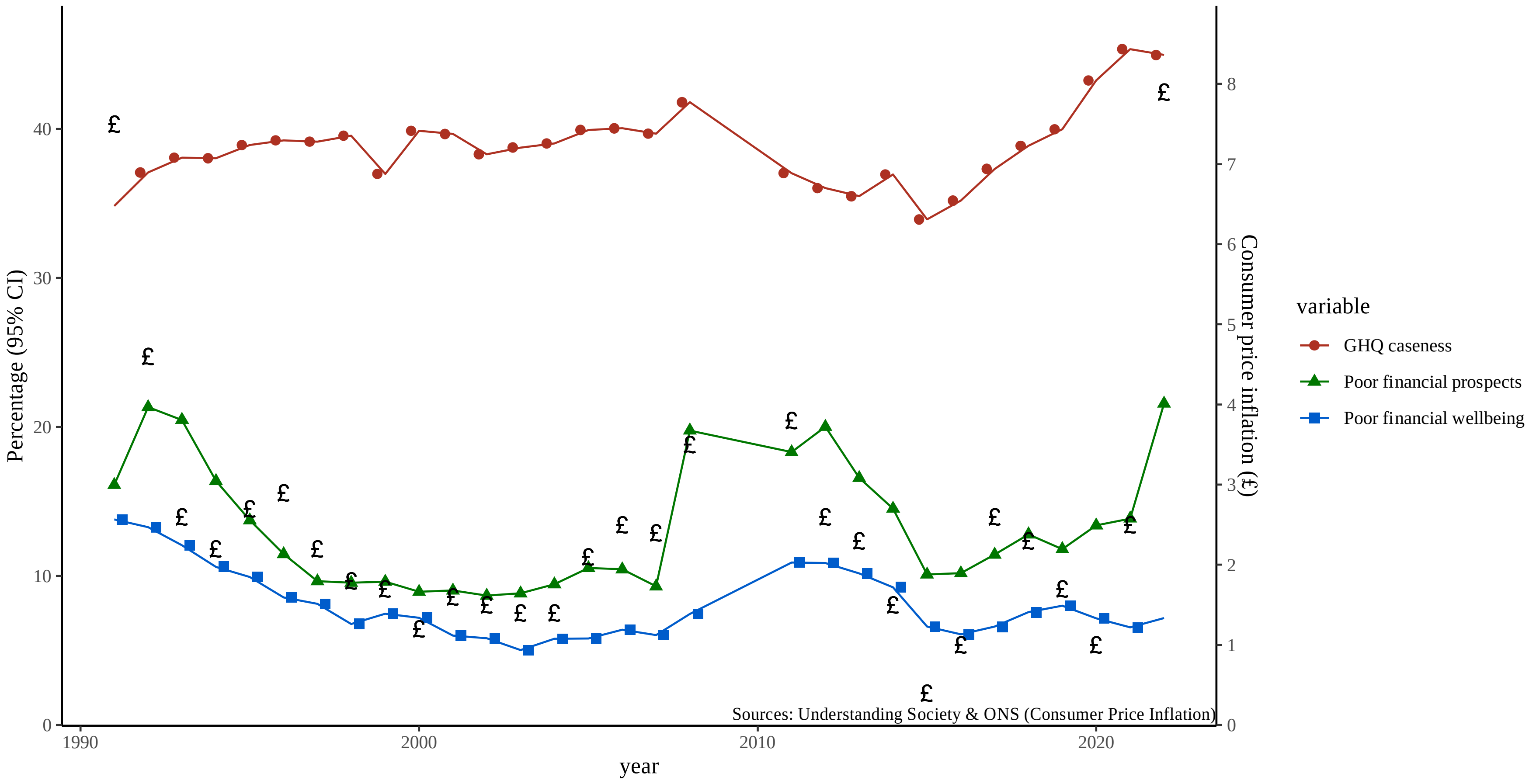

Note: Percentages and 95%CI calculated yearly based on the combination of the British Household Panel Survey (BHPS) and the UK Longitudinal Survey (UKHLS) for the full population aged 16 and over. GHQ-caseness is calculated based on GHQ-36 with a cut-point at 9. The consumer price inflation is derived from Office for National Statistics (2024). Ref.: Office for National Statistics. Consumer price inflation time series. Published 2024. Accessed January 3, 2024. https://www.ons.gov.uk/economy/inflationandpriceindices/datasets/consumerpriceindices. Data are weighted using USoc-provided yearly cross-sectional weights.

### Supplementary file S.5. LGM results for cohort 1 (2016-2019)

#### Financial hardship (cross-sectional baseline weight)

| | Employment adjustment | | | | Socio-economic adjustment | | | | Demographic adjustment | | | | Unadjusted | | | |
|---|---|---|---|---|---|---|---|---|---|---|---|---|---|---|---|---|
| **term** | **Estimate** | **SE** | **CI lower** | **CI upper** | **Estimate** | **SE** | **CI lower** | **CI upper** | **Estimate** | **SE** | **CI lower** | **CI upper** | **Estimate** | **SE** | **CI lower** | **CI upper** |
| **ghq_16 ~~ ghq_16** | 11.957 | 0.468 | 11.039 | 12.875 | 11.979 | 0.472 | 11.054 | 12.904 | 11.854 | 0.431 | 11.010 | 12.698 | 11.722 | 0.399 | 10.940 | 12.504 |
| **ghq_17 ~~ ghq_17** | 12.924 | 0.383 | 12.173 | 13.674 | 12.896 | 0.382 | 12.147 | 13.646 | 13.273 | 0.370 | 12.548 | 13.997 | 13.062 | 0.343 | 12.389 | 13.734 |
| **ghq_18 ~~ ghq_18** | 13.419 | 0.391 | 12.652 | 14.186 | 13.434 | 0.393 | 12.663 | 14.205 | 13.113 | 0.349 | 12.428 | 13.798 | 13.037 | 0.323 | 12.404 | 13.670 |
| **ghq_19 ~~ ghq_19** | 11.529 | 0.471 | 10.606 | 12.452 | 11.525 | 0.476 | 10.592 | 12.458 | 11.483 | 0.431 | 10.637 | 12.329 | 11.305 | 0.399 | 10.524 | 12.087 |
| **i ~ age** | -0.010 | 0.005 | -0.019 | 0.000 | -0.015 | 0.004 | -0.022 | -0.008 | -0.020 | 0.003 | -0.025 | -0.014 | | | | |
| **i ~ country_LondonandSouthEngland** | -0.065 | 0.097 | -0.256 | 0.126 | -0.085 | 0.100 | -0.280 | 0.110 | -0.090 | 0.094 | -0.275 | 0.094 | | | | |
| **i ~ coupleandchildren** | -0.310 | 0.129 | -0.564 | -0.056 | -0.373 | 0.131 | -0.629 | -0.116 | -0.343 | 0.116 | -0.570 | -0.116 | | | | |
| **i ~ empl_16_maternity** | 0.514 | 0.268 | -0.012 | 1.040 | | | | | | | | | | | | |
| **i ~ empl_16_retired** | -0.253 | 0.145 | -0.537 | 0.032 | | | | | | | | | | | | |
| **i ~ empl_16_self** | -0.270 | 0.154 | -0.571 | 0.032 | | | | | | | | | | | | |
| **i ~ empl_16_sick** | 5.179 | 0.461 | 4.276 | 6.082 | | | | | | | | | | | | |
| **i ~ empl_16_student** | 0.171 | 0.266 | -0.350 | 0.692 | | | | | | | | | | | | |
| **i ~ empl_16_unemployed** | 1.002 | 0.359 | 0.300 | 1.705 | | | | | | | | | | | | |
| **i ~ empl_maternity** | 0.113 | 0.317 | -0.509 | 0.735 | 0.346 | 0.310 | -0.260 | 0.953 | 0.394 | 0.279 | -0.152 | 0.941 | | | | |
| **i ~ FinancialHardship2016** | 2.378 | 0.138 | 2.108 | 2.648 | 2.670 | 0.140 | 2.395 | 2.945 | 2.974 | 0.126 | 2.727 | 3.222 | 3.169 | 0.117 | 2.940 | 3.399 |
| **i ~ IMD_1** | -0.155 | 0.180 | -0.507 | 0.197 | 0.024 | 0.184 | -0.338 | 0.385 | | | | | | | | |
| **i ~ IMD_2** | -0.037 | 0.157 | -0.345 | 0.271 | -0.022 | 0.159 | -0.334 | 0.289 | | | | | | | | |
| **i ~ IMD_4** | -0.165 | 0.139 | -0.436 | 0.107 | -0.185 | 0.142 | -0.463 | 0.093 | | | | | | | | |
| **i ~ IMD_5** | -0.229 | 0.134 | -0.492 | 0.034 | -0.225 | 0.136 | -0.490 | 0.041 | | | | | | | | |
| **i ~ isced** | 0.361 | 0.130 | 0.107 | 0.615 | 0.296 | 0.130 | 0.040 | 0.552 | 0.161 | 0.120 | -0.075 | 0.397 | | | | |
| **i ~ own_mortgage** | 0.175 | 0.123 | -0.066 | 0.415 | 0.229 | 0.120 | -0.007 | 0.465 | | | | | | | | |
| **i ~ own_rent** | 0.438 | 0.156 | 0.133 | 0.743 | 0.738 | 0.157 | 0.430 | 1.047 | | | | | | | | |
| **i ~ save** | -0.177 | 0.098 | -0.370 | 0.016 | -0.259 | 0.100 | -0.454 | -0.064 | | | | | | | | |
| **i ~ sex_female** | 0.997 | 0.097 | 0.808 | 1.187 | 0.989 | 0.099 | 0.795 | 1.182 | 1.024 | 0.090 | 0.848 | 1.201 | | | | |
| **i ~ single** | 0.402 | 0.127 | 0.152 | 0.652 | 0.568 | 0.132 | 0.310 | 0.826 | 0.697 | 0.119 | 0.464 | 0.930 | | | | |
| **i ~ singleandchildren** | 0.169 | 0.283 | -0.387 | 0.724 | 0.053 | 0.285 | -0.507 | 0.612 | 0.356 | 0.261 | -0.155 | 0.867 | | | | |
| **i ~~ i** | 13.932 | 0.491 | 12.970 | 14.893 | 14.897 | 0.518 | 13.882 | 15.911 | 15.119 | 0.480 | 14.178 | 16.061 | 15.608 | 0.454 | 14.718 | 16.498 |
| **i ~~ s** | -0.965 | 0.160 | -1.279 | -0.651 | -0.976 | 0.163 | -1.295 | -0.657 | -1.051 | 0.151 | -1.347 | -0.756 | -1.008 | 0.140 | -1.283 | -0.733 |
| **s ~ age** | -0.006 | 0.002 | -0.010 | -0.002 | -0.005 | 0.001 | -0.008 | -0.003 | -0.004 | 0.001 | -0.006 | -0.002 | | | | |
| **s ~ country_LondonandSouthEngland** | 0.060 | 0.036 | -0.011 | 0.131 | 0.053 | 0.036 | -0.018 | 0.125 | 0.063 | 0.034 | -0.004 | 0.131 | | | | |
| **s ~ coupleandchildren** | 0.053 | 0.050 | -0.046 | 0.152 | 0.026 | 0.050 | -0.072 | 0.124 | 0.038 | 0.044 | -0.048 | 0.125 | | | | |
| **s ~ empl_17_maternity** | -0.468 | 0.118 | -0.699 | -0.237 | | | | | | | | | | | | |
| **s ~ empl_17_retired** | 0.011 | 0.088 | -0.162 | 0.184 | | | | | | | | | | | | |
| **s ~ empl_17_self** | -0.105 | 0.093 | -0.287 | 0.077 | | | | | | | | | | | | |
| **s ~ empl_17_sick** | -0.799 | 0.216 | -1.223 | -0.376 | | | | | | | | | | | | |
| **s ~ empl_17_student** | -0.091 | 0.132 | -0.350 | 0.169 | | | | | | | | | | | | |
| **s ~ empl_17_unemployed** | -0.502 | 0.134 | -0.765 | -0.240 | | | | | | | | | | | | |
| **s ~ empl_18_maternity** | 0.284 | 0.124 | 0.042 | 0.526 | | | | | | | | | | | | |
| **s ~ empl_18_retired** | 0.050 | 0.098 | -0.142 | 0.243 | | | | | | | | | | | | |
| **s ~ empl_18_self** | 0.119 | 0.117 | -0.110 | 0.347 | | | | | | | | | | | | |
| **s ~ empl_18_sick** | -0.055 | 0.223 | -0.492 | 0.383 | | | | | | | | | | | | |
| **s ~ empl_18_student** | 0.225 | 0.170 | -0.107 | 0.558 | | | | | | | | | | | | |
| **s ~ empl_18_unemployed** | 0.509 | 0.156 | 0.203 | 0.816 | | | | | | | | | | | | |
| **s ~ empl_maternity** | 0.102 | 0.129 | -0.152 | 0.355 | -0.030 | 0.099 | -0.223 | 0.164 | -0.009 | 0.089 | -0.183 | 0.165 | | | | |
| **s ~ empl_retired** | 0.004 | 0.086 | -0.163 | 0.172 | | | | | | | | | | | | |
| **s ~ empl_self** | -0.010 | 0.100 | -0.207 | 0.187 | | | | | | | | | | | | |
| **s ~ empl_sick** | 1.444 | 0.215 | 1.022 | 1.865 | | | | | | | | | | | | |
| **s ~ empl_student** | -0.274 | 0.161 | -0.589 | 0.040 | | | | | | | | | | | | |
| **s ~ empl_unemployed** | 0.381 | 0.148 | 0.091 | 0.672 | | | | | | | | | | | | |

| | | | | | | | | | | | | | | | | |
|---|---|---|---|---|---|---|---|---|---|---|---|---|---|---|---|---|
| **s ~ FinancialHardship2016** | -0.654 | 0.056 | -0.764 | -0.544 | -0.651 | 0.056 | -0.762 | -0.541 | -0.672 | 0.051 | -0.772 | -0.572 | -0.671 | 0.047 | -0.763 | -0.579 |
| **s ~ FinancialHardship2017to2019_Constant_hardship** | 0.870 | 0.066 | 0.741 | 1.000 | 0.927 | 0.068 | 0.794 | 1.060 | 0.981 | 0.062 | 0.860 | 1.103 | 1.013 | 0.057 | 0.901 | 1.124 |
| **s ~ FinancialHardship2017to2019_b+3 hardship** | 0.740 | 0.086 | 0.572 | 0.907 | 0.765 | 0.087 | 0.595 | 0.936 | 0.729 | 0.077 | 0.578 | 0.880 | 0.782 | 0.072 | 0.640 | 0.924 |
| **s ~ FinancialHardship2017to2019_b+1 and b+3 hardship** | 0.463 | 0.105 | 0.258 | 0.668 | 0.531 | 0.101 | 0.332 | 0.729 | 0.559 | 0.091 | 0.380 | 0.737 | 0.632 | 0.087 | 0.462 | 0.803 |
| **s ~ FinancialHardship2017to2019_b+1 hardship** | 0.119 | 0.065 | -0.009 | 0.246 | 0.097 | 0.066 | -0.032 | 0.226 | 0.101 | 0.059 | -0.016 | 0.217 | 0.142 | 0.054 | 0.035 | 0.249 |
| **s ~ FinancialHardship2017to2019_b+1 and b+2 hardship** | 0.222 | 0.093 | 0.040 | 0.403 | 0.234 | 0.094 | 0.049 | 0.420 | 0.261 | 0.084 | 0.096 | 0.427 | 0.289 | 0.078 | 0.137 | 0.442 |
| **s ~ FinancialHardship2017to2019_b+2 and b+3 hardshop** | 0.803 | 0.082 | 0.642 | 0.963 | 0.872 | 0.085 | 0.706 | 1.039 | 0.949 | 0.078 | 0.796 | 1.102 | 1.002 | 0.074 | 0.857 | 1.147 |
| **s ~ FinancialHardship2017to2019_b+2 hardship** | 0.394 | 0.070 | 0.257 | 0.531 | 0.428 | 0.067 | 0.296 | 0.560 | 0.420 | 0.061 | 0.300 | 0.539 | 0.466 | 0.057 | 0.354 | 0.578 |
| **s ~ IMD_1** | -0.010 | 0.062 | -0.133 | 0.112 | 0.007 | 0.063 | -0.117 | 0.131 | | | | | | | | |
| **s ~ IMD_2** | -0.020 | 0.056 | -0.130 | 0.090 | -0.019 | 0.056 | -0.130 | 0.091 | | | | | | | | |
| **s ~ IMD_4** | 0.077 | 0.051 | -0.023 | 0.177 | 0.075 | 0.051 | -0.025 | 0.176 | | | | | | | | |
| **s ~ IMD_5** | 0.063 | 0.050 | -0.034 | 0.161 | 0.061 | 0.050 | -0.036 | 0.159 | | | | | | | | |
| **s ~ isced** | -0.141 | 0.048 | -0.235 | -0.047 | -0.133 | 0.049 | -0.228 | -0.038 | -0.135 | 0.047 | -0.227 | -0.043 | | | | |
| **s ~ own_mortgage** | -0.029 | 0.046 | -0.120 | 0.062 | -0.039 | 0.045 | -0.127 | 0.050 | | | | | | | | |
| **s ~ own_rent** | -0.012 | 0.057 | -0.124 | 0.100 | 0.016 | 0.057 | -0.095 | 0.128 | | | | | | | | |
| **s ~ save** | -0.046 | 0.036 | -0.117 | 0.025 | -0.057 | 0.036 | -0.128 | 0.013 | | | | | | | | |
| **s ~ sex_female** | -0.055 | 0.035 | -0.124 | 0.015 | -0.061 | 0.036 | -0.130 | 0.009 | -0.057 | 0.033 | -0.121 | 0.006 | | | | |
| **s ~ single** | -0.077 | 0.046 | -0.167 | 0.014 | -0.054 | 0.046 | -0.145 | 0.036 | -0.043 | 0.041 | -0.124 | 0.038 | | | | |
| **s ~ singleandchildren** | -0.033 | 0.102 | -0.233 | 0.168 | -0.063 | 0.102 | -0.263 | 0.137 | -0.030 | 0.093 | -0.212 | 0.152 | | | | |
| **s ~~ s** | 0.681 | 0.086 | 0.513 | 0.848 | 0.712 | 0.087 | 0.542 | 0.883 | 0.750 | 0.080 | 0.592 | 0.907 | 0.741 | 0.075 | 0.595 | 0.887 |

### Financial hardship (cross-sectional baseline weight and ipw for attrition)

| | Employment adjustment | | | | Socio-economic adjustment | | | | Demographic adjustment | | | | Unadjusted | | | |
|---|---|---|---|---|---|---|---|---|---|---|---|---|---|---|---|---|
| **term** | **Estimate** | **SE** | **CI lower** | **CI upper** | **Estimate** | **SE** | **CI lower** | **CI upper** | **Estimate** | **SE** | **CI lower** | **CI upper** | **Estimate** | **SE** | **CI lower** | **CI upper** |
| **ghq_16 ~~ ghq_16** | 12.033 | 0.491 | 11.071 | 12.994 | 12.065 | 0.494 | 11.096 | 13.034 | 12.066 | 0.496 | 11.095 | 13.038 | 12.103 | 0.499 | 11.124 | 13.082 |
| **ghq_17 ~~ ghq_17** | 13.186 | 0.407 | 12.389 | 13.983 | 13.150 | 0.406 | 12.355 | 13.946 | 13.144 | 0.406 | 12.348 | 13.940 | 13.120 | 0.407 | 12.322 | 13.918 |
| **ghq_18 ~~ ghq_18** | 13.694 | 0.418 | 12.875 | 14.513 | 13.700 | 0.420 | 12.877 | 14.522 | 13.701 | 0.421 | 12.876 | 14.526 | 13.694 | 0.421 | 12.867 | 14.520 |
| **ghq_19 ~~ ghq_19** | 11.885 | 0.499 | 10.907 | 12.864 | 11.910 | 0.507 | 10.915 | 12.904 | 11.919 | 0.511 | 10.919 | 12.920 | 11.939 | 0.517 | 10.926 | 12.952 |
| **i ~ age** | -0.009 | 0.005 | -0.019 | 0.001 | -0.014 | 0.004 | -0.022 | -0.007 | -0.019 | 0.003 | -0.026 | -0.012 | | | | |
| **i ~ country_LondonandSouthEngland** | -0.099 | 0.101 | -0.297 | 0.098 | -0.114 | 0.103 | -0.317 | 0.088 | -0.138 | 0.104 | -0.342 | 0.065 | | | | |
| **i ~ coupleandchildren** | -0.324 | 0.134 | -0.587 | -0.062 | -0.382 | 0.135 | -0.647 | -0.118 | -0.310 | 0.130 | -0.566 | -0.054 | | | | |
| **i ~ empl_16_maternity** | 0.485 | 0.281 | -0.065 | 1.035 | | | | | | | | | | | | |
| **i ~ empl_16_retired** | -0.280 | 0.149 | -0.571 | 0.011 | | | | | | | | | | | | |
| **i ~ empl_16_self** | -0.236 | 0.157 | -0.544 | 0.072 | | | | | | | | | | | | |
| **i ~ empl_16_sick** | 5.240 | 0.479 | 4.302 | 6.179 | | | | | | | | | | | | |
| **i ~ empl_16_student** | 0.214 | 0.269 | -0.314 | 0.742 | | | | | | | | | | | | |
| **i ~ empl_16_unemployed** | 0.969 | 0.378 | 0.228 | 1.710 | | | | | | | | | | | | |
| **i ~ empl_maternity** | 0.143 | 0.334 | -0.512 | 0.799 | 0.360 | 0.326 | -0.280 | 0.999 | 0.494 | 0.324 | -0.142 | 1.130 | | | | |
| **i ~ FinancialHardship2016** | 2.389 | 0.143 | 2.109 | 2.669 | 2.676 | 0.145 | 2.391 | 2.961 | 2.932 | 0.143 | 2.652 | 3.212 | 3.085 | 0.143 | 2.805 | 3.365 |
| **i ~ IMD_1** | -0.116 | 0.186 | -0.480 | 0.247 | 0.070 | 0.191 | -0.304 | 0.443 | | | | | | | | |
| **i ~ IMD_2** | 0.026 | 0.162 | -0.292 | 0.344 | 0.036 | 0.164 | -0.286 | 0.357 | | | | | | | | |
| **i ~ IMD_4** | -0.150 | 0.143 | -0.430 | 0.130 | -0.176 | 0.146 | -0.463 | 0.111 | | | | | | | | |
| **i ~ IMD_5** | -0.205 | 0.138 | -0.476 | 0.066 | -0.201 | 0.140 | -0.475 | 0.073 | | | | | | | | |
| **i ~ isced** | 0.369 | 0.134 | 0.106 | 0.631 | 0.304 | 0.135 | 0.040 | 0.568 | 0.198 | 0.134 | -0.065 | 0.462 | | | | |
| **i ~ own_mortgage** | 0.165 | 0.126 | -0.083 | 0.412 | 0.230 | 0.124 | -0.013 | 0.472 | | | | | | | | |
| **i ~ own_rent** | 0.441 | 0.159 | 0.129 | 0.752 | 0.745 | 0.161 | 0.430 | 1.061 | | | | | | | | |
| **i ~ save** | -0.134 | 0.102 | -0.335 | 0.067 | -0.219 | 0.104 | -0.422 | -0.016 | | | | | | | | |
| **i ~ sex_female** | 1.023 | 0.101 | 0.826 | 1.221 | 1.007 | 0.103 | 0.806 | 1.209 | 1.007 | 0.103 | 0.805 | 1.209 | | | | |
| **i ~ single** | 0.353 | 0.132 | 0.094 | 0.612 | 0.527 | 0.137 | 0.258 | 0.797 | 0.730 | 0.138 | 0.459 | 1.001 | | | | |
| **i ~ singleandchildren** | 0.140 | 0.294 | -0.437 | 0.716 | 0.022 | 0.297 | -0.559 | 0.604 | 0.298 | 0.294 | -0.279 | 0.875 | | | | |
| **i ~~ i** | 14.075 | 0.517 | 13.061 | 15.089 | 15.057 | 0.548 | 13.984 | 16.131 | 15.169 | 0.553 | 14.084 | 16.254 | 15.665 | 0.567 | 14.554 | 16.777 |
| **i ~~ s** | -0.973 | 0.168 | -1.302 | -0.644 | -0.972 | 0.171 | -1.308 | -0.637 | -0.983 | 0.172 | -1.320 | -0.646 | -0.994 | 0.174 | -1.334 | -0.654 |
| **s ~ age** | -0.007 | 0.002 | -0.010 | -0.003 | -0.006 | 0.001 | -0.008 | -0.003 | -0.005 | 0.001 | -0.008 | -0.003 | | | | |
| **s ~ country_LondonandSouthEngland** | 0.075 | 0.038 | 0.001 | 0.148 | 0.068 | 0.038 | -0.007 | 0.142 | 0.070 | 0.038 | -0.004 | 0.144 | | | | |
| **s ~ coupleandchildren** | 0.052 | 0.052 | -0.051 | 0.154 | 0.021 | 0.052 | -0.080 | 0.123 | 0.015 | 0.050 | -0.084 | 0.114 | | | | |
| **s ~ empl_17_maternity** | -0.504 | 0.123 | -0.745 | -0.262 | | | | | | | | | | | | |
| **s ~ empl_17_retired** | -0.003 | 0.090 | -0.179 | 0.174 | | | | | | | | | | | | |

| | | | | | | | | | | | | | | | | |
|---|---|---|---|---|---|---|---|---|---|---|---|---|---|---|---|---|
| **s ~ empl_17_self** | -0.105 | 0.094 | -0.288 | 0.079 | | | | | | | | | | | | |
| **s ~ empl_17_sick** | -0.814 | 0.226 | -1.258 | -0.371 | | | | | | | | | | | | |
| **s ~ empl_17_student** | -0.095 | 0.136 | -0.361 | 0.171 | | | | | | | | | | | | |
| **s ~ empl_17_unemployed** | -0.519 | 0.143 | -0.800 | -0.239 | | | | | | | | | | | | |
| **s ~ empl_18_maternity** | 0.253 | 0.128 | 0.002 | 0.504 | | | | | | | | | | | | |
| **s ~ empl_18_retired** | 0.072 | 0.100 | -0.124 | 0.269 | | | | | | | | | | | | |
| **s ~ empl_18_self** | 0.101 | 0.124 | -0.142 | 0.344 | | | | | | | | | | | | |
| **s ~ empl_18_sick** | -0.125 | 0.238 | -0.591 | 0.341 | | | | | | | | | | | | |
| **s ~ empl_18_student** | 0.227 | 0.176 | -0.118 | 0.571 | | | | | | | | | | | | |
| **s ~ empl_18_unemployed** | 0.537 | 0.168 | 0.208 | 0.867 | | | | | | | | | | | | |
| **s ~ empl_maternity** | 0.147 | 0.135 | -0.118 | 0.413 | -0.017 | 0.105 | -0.222 | 0.188 | -0.006 | 0.104 | -0.210 | 0.198 | | | | |
| **s ~ empl_retired** | -0.003 | 0.086 | -0.172 | 0.166 | | | | | | | | | | | | |
| **s ~ empl_self** | -0.014 | 0.105 | -0.221 | 0.193 | | | | | | | | | | | | |
| **s ~ empl_sick** | 1.534 | 0.228 | 1.087 | 1.981 | | | | | | | | | | | | |
| **s ~ empl_student** | -0.304 | 0.166 | -0.630 | 0.022 | | | | | | | | | | | | |
| **s ~ empl_unemployed** | 0.368 | 0.155 | 0.065 | 0.671 | | | | | | | | | | | | |
| **s ~ FinancialHardship2016** | -0.662 | 0.058 | -0.776 | -0.547 | -0.658 | 0.059 | -0.773 | -0.543 | -0.674 | 0.058 | -0.787 | -0.561 | -0.690 | 0.058 | -0.803 | -0.576 |
| **s ~ FinancialHardship2017to2019_Constant_hardship** | 0.891 | 0.068 | 0.757 | 1.025 | 0.950 | 0.070 | 0.812 | 1.088 | 1.021 | 0.071 | 0.883 | 1.159 | 1.068 | 0.071 | 0.929 | 1.207 |
| **s ~ FinancialHardship2017to2019_b+3 hardship** | 0.735 | 0.088 | 0.564 | 0.907 | 0.763 | 0.089 | 0.588 | 0.938 | 0.808 | 0.089 | 0.633 | 0.982 | 0.894 | 0.090 | 0.719 | 1.070 |
| **s ~ FinancialHardship2017to2019_b+1 and b+3 hardship** | 0.502 | 0.111 | 0.284 | 0.721 | 0.567 | 0.108 | 0.355 | 0.779 | 0.629 | 0.108 | 0.417 | 0.840 | 0.686 | 0.110 | 0.469 | 0.902 |
| **s ~ FinancialHardship2017to2019_b+1 hardship** | 0.109 | 0.066 | -0.021 | 0.239 | 0.081 | 0.067 | -0.051 | 0.212 | 0.118 | 0.067 | -0.013 | 0.249 | 0.163 | 0.066 | 0.034 | 0.292 |
| **s ~ FinancialHardship2017to2019_b+1 and b+2 hardship** | 0.269 | 0.096 | 0.082 | 0.457 | 0.284 | 0.097 | 0.094 | 0.474 | 0.316 | 0.097 | 0.126 | 0.506 | 0.366 | 0.097 | 0.176 | 0.557 |
| **s ~ FinancialHardship2017to2019_b+2 and b+3 hardshop** | 0.817 | 0.085 | 0.649 | 0.984 | 0.891 | 0.089 | 0.717 | 1.065 | 0.951 | 0.089 | 0.777 | 1.124 | 1.020 | 0.089 | 0.845 | 1.194 |
| **s ~ FinancialHardship2017to2019_b+2 hardship** | 0.400 | 0.073 | 0.257 | 0.543 | 0.436 | 0.070 | 0.299 | 0.573 | 0.474 | 0.070 | 0.337 | 0.611 | 0.518 | 0.069 | 0.381 | 0.654 |
| **s ~ IMD_1** | -0.012 | 0.064 | -0.138 | 0.114 | 0.006 | 0.065 | -0.121 | 0.133 | | | | | | | | |
| **s ~ IMD_2** | -0.025 | 0.059 | -0.140 | 0.090 | -0.026 | 0.059 | -0.141 | 0.090 | | | | | | | | |
| **s ~ IMD_4** | 0.085 | 0.053 | -0.019 | 0.190 | 0.083 | 0.053 | -0.022 | 0.187 | | | | | | | | |
| **s ~ IMD_5** | 0.065 | 0.051 | -0.035 | 0.166 | 0.062 | 0.051 | -0.039 | 0.163 | | | | | | | | |
| **s ~ isced** | -0.144 | 0.050 | -0.241 | -0.046 | -0.135 | 0.050 | -0.234 | -0.036 | -0.139 | 0.050 | -0.237 | -0.040 | | | | |
| **s ~ own_mortgage** | -0.031 | 0.048 | -0.125 | 0.062 | -0.037 | 0.047 | -0.129 | 0.054 | | | | | | | | |
| **s ~ own_rent** | 0.001 | 0.059 | -0.114 | 0.116 | 0.033 | 0.058 | -0.081 | 0.148 | | | | | | | | |
| **s ~ save** | -0.059 | 0.038 | -0.133 | 0.016 | -0.070 | 0.038 | -0.143 | 0.004 | | | | | | | | |
| **s ~ sex_female** | -0.064 | 0.037 | -0.136 | 0.009 | -0.073 | 0.037 | -0.146 | 0.000 | -0.070 | 0.037 | -0.143 | 0.003 | | | | |
| **s ~ single** | -0.064 | 0.048 | -0.158 | 0.031 | -0.035 | 0.048 | -0.129 | 0.059 | -0.035 | 0.048 | -0.128 | 0.059 | | | | |
| **s ~ singleandchildren** | -0.024 | 0.105 | -0.231 | 0.183 | -0.062 | 0.105 | -0.268 | 0.144 | -0.061 | 0.105 | -0.267 | 0.144 | | | | |
| **s ~~ s** | 0.681 | 0.090 | 0.505 | 0.858 | 0.712 | 0.092 | 0.532 | 0.893 | 0.720 | 0.092 | 0.539 | 0.901 | 0.732 | 0.094 | 0.548 | 0.915 |

### Financial uncertainty (cross-sectional baseline weight)

| | Employment adjustment | | | | Socio-economic adjustment | | | | Demographic adjustment | | | | Unadjusted | | | |
|---|---|---|---|---|---|---|---|---|---|---|---|---|---|---|---|---|
| **term** | **Estimate** | **SE** | **CI lower** | **CI upper** | **Estimate** | **SE** | **CI lower** | **CI upper** | **Estimate** | **SE** | **CI lower** | **CI upper** | **Estimate** | **SE** | **CI lower** | **CI upper** |
| **ghq_16 ~~ ghq_16** | 11.650 | 0.479 | 10.711 | 12.588 | 11.670 | 0.481 | 10.727 | 12.613 | 11.579 | 0.442 | 10.714 | 12.444 | 11.464 | 0.410 | 10.661 | 12.267 |
| **ghq_17 ~~ ghq_17** | 12.816 | 0.393 | 12.046 | 13.587 | 12.788 | 0.393 | 12.018 | 13.557 | 13.157 | 0.383 | 12.406 | 13.907 | 12.958 | 0.356 | 12.261 | 13.655 |
| **ghq_18 ~~ ghq_18** | 13.261 | 0.400 | 12.477 | 14.045 | 13.270 | 0.402 | 12.483 | 14.057 | 12.949 | 0.359 | 12.246 | 13.653 | 12.887 | 0.333 | 12.233 | 13.540 |
| **ghq_19 ~~ ghq_19** | 11.401 | 0.483 | 10.455 | 12.347 | 11.415 | 0.488 | 10.459 | 12.371 | 11.367 | 0.445 | 10.494 | 12.239 | 11.231 | 0.415 | 10.418 | 12.044 |
| **i ~ age** | -0.008 | 0.005 | -0.017 | 0.002 | -0.015 | 0.004 | -0.023 | -0.008 | -0.024 | 0.003 | -0.030 | -0.018 | | | | |
| **i ~ country_LondonandSouthEngland** | -0.048 | 0.099 | -0.243 | 0.147 | -0.061 | 0.102 | -0.260 | 0.139 | -0.072 | 0.097 | -0.263 | 0.119 | | | | |
| **i ~ coupleandchildren** | -0.215 | 0.134 | -0.478 | 0.047 | -0.250 | 0.135 | -0.515 | 0.015 | -0.111 | 0.120 | -0.346 | 0.124 | | | | |
| **i ~ empl_16_maternity** | 0.615 | 0.278 | 0.070 | 1.161 | | | | | | | | | | | | |
| **i ~ empl_16_retired** | -0.432 | 0.149 | -0.724 | -0.140 | | | | | | | | | | | | |
| **i ~ empl_16_self** | -0.165 | 0.156 | -0.471 | 0.140 | | | | | | | | | | | | |
| **i ~ empl_16_sick** | 5.557 | 0.504 | 4.569 | 6.545 | | | | | | | | | | | | |
| **i ~ empl_16_student** | 0.157 | 0.278 | -0.388 | 0.702 | | | | | | | | | | | | |
| **i ~ empl_16_unemployed** | 1.266 | 0.371 | 0.539 | 1.993 | | | | | | | | | | | | |
| **i ~ empl_maternity** | 0.081 | 0.319 | -0.544 | 0.705 | 0.377 | 0.305 | -0.220 | 0.975 | 0.569 | 0.273 | 0.034 | 1.104 | | | | |
| **i ~ FinancialUncertainty2016** | 1.725 | 0.177 | 1.379 | 2.071 | 1.904 | 0.183 | 1.544 | 2.263 | 1.931 | 0.172 | 1.593 | 2.268 | 2.003 | 0.164 | 1.682 | 2.325 |
| **i ~ IMD_1** | 0.093 | 0.183 | -0.265 | 0.451 | 0.304 | 0.188 | -0.065 | 0.673 | | | | | | | | |
| **i ~ IMD_2** | 0.104 | 0.161 | -0.211 | 0.419 | 0.125 | 0.163 | -0.195 | 0.446 | | | | | | | | |
| **i ~ IMD_4** | -0.156 | 0.140 | -0.431 | 0.118 | -0.190 | 0.144 | -0.472 | 0.092 | | | | | | | | |

| | | | | | | | | | | | | | | | | | | | |
|---|---|---|---|---|---|---|---|---|---|---|---|---|---|---|---|---|---|---|---|
| **i ~ IMD_5** | -0.242 | 0.136 | -0.508 | 0.024 | | -0.252 | 0.138 | -0.522 | 0.019 | | | | | | | | | | |
| **i ~ isced** | 0.267 | 0.133 | 0.006 | 0.528 | | 0.184 | 0.134 | -0.079 | 0.447 | | -0.107 | 0.125 | -0.352 | 0.137 | | | | | |
| **i ~ own_mortgage** | 0.353 | 0.125 | 0.107 | 0.598 | | 0.469 | 0.123 | 0.227 | 0.711 | | | | | | | | | | |
| **i ~ own_rent** | 0.922 | 0.161 | 0.606 | 1.237 | | 1.329 | 0.165 | 1.006 | 1.652 | | | | | | | | | | |
| **i ~ save** | -0.417 | 0.100 | -0.612 | -0.222 | | -0.550 | 0.101 | -0.748 | -0.353 | | | | | | | | | | |
| **i ~ sex_female** | 0.969 | 0.099 | 0.774 | 1.164 | | 0.943 | 0.102 | 0.744 | 1.143 | | 0.979 | 0.094 | 0.795 | 1.163 | | | | | |
| **i ~ single** | 0.450 | 0.131 | 0.193 | 0.706 | | 0.645 | 0.136 | 0.378 | 0.912 | | 0.983 | 0.126 | 0.735 | 1.230 | | | | | |
| **i ~ singleandchildren** | 0.445 | 0.292 | -0.128 | 1.017 | | 0.370 | 0.294 | -0.205 | 0.946 | | 1.027 | 0.264 | 0.509 | 1.545 | | | | | |
| **i ~~ i** | 14.229 | 0.512 | 13.226 | 15.231 | | 15.329 | 0.548 | 14.255 | 16.403 | | 15.914 | 0.523 | 14.890 | 16.939 | | 16.657 | 0.501 | 15.675 | 17.639 |
| **i ~~ s** | -1.054 | 0.165 | -1.377 | -0.731 | | -1.071 | 0.168 | -1.401 | -0.741 | | -1.086 | 0.157 | -1.394 | -0.778 | | -0.979 | 0.147 | -1.266 | -0.692 |
| **s ~ age** | -0.008 | 0.002 | -0.011 | -0.004 | | -0.007 | 0.001 | -0.010 | -0.004 | | -0.006 | 0.001 | -0.009 | -0.004 | | | | | |
| **s ~ country_LondonandSouthEngland** | 0.069 | 0.037 | -0.003 | 0.141 | | 0.064 | 0.037 | -0.009 | 0.136 | | 0.068 | 0.035 | -0.001 | 0.136 | | | | | |
| **s ~ coupleandchildren** | 0.083 | 0.051 | -0.017 | 0.183 | | 0.059 | 0.051 | -0.040 | 0.159 | | 0.086 | 0.045 | -0.001 | 0.174 | | | | | |
| **s ~ empl_17_maternity** | -0.487 | 0.123 | -0.728 | -0.247 | | | | | | | | | | | | | | | |
| **s ~ empl_17_retired** | 0.081 | 0.090 | -0.094 | 0.257 | | | | | | | | | | | | | | | |
| **s ~ empl_17_self** | -0.054 | 0.093 | -0.237 | 0.129 | | | | | | | | | | | | | | | |
| **s ~ empl_17_sick** | -0.775 | 0.219 | -1.205 | -0.346 | | | | | | | | | | | | | | | |
| **s ~ empl_17_student** | -0.055 | 0.143 | -0.334 | 0.224 | | | | | | | | | | | | | | | |
| **s ~ empl_17_unemployed** | -0.512 | 0.139 | -0.784 | -0.239 | | | | | | | | | | | | | | | |
| **s ~ empl_18_maternity** | 0.294 | 0.130 | 0.040 | 0.548 | | | | | | | | | | | | | | | |
| **s ~ empl_18_retired** | 0.036 | 0.100 | -0.160 | 0.231 | | | | | | | | | | | | | | | |
| **s ~ empl_18_self** | 0.103 | 0.121 | -0.134 | 0.341 | | | | | | | | | | | | | | | |
| **s ~ empl_18_sick** | 0.016 | 0.233 | -0.441 | 0.474 | | | | | | | | | | | | | | | |
| **s ~ empl_18_student** | 0.133 | 0.182 | -0.224 | 0.490 | | | | | | | | | | | | | | | |
| **s ~ empl_18_unemployed** | 0.552 | 0.166 | 0.227 | 0.877 | | | | | | | | | | | | | | | |
| **s ~ empl_maternity** | 0.063 | 0.132 | -0.196 | 0.322 | | -0.057 | 0.101 | -0.256 | 0.142 | | -0.007 | 0.091 | -0.184 | 0.171 | | | | | |
| **s ~ empl_retired** | -0.079 | 0.086 | -0.248 | 0.089 | | | | | | | | | | | | | | | |
| **s ~ empl_self** | -0.040 | 0.101 | -0.238 | 0.159 | | | | | | | | | | | | | | | |
| **s ~ empl_sick** | 1.311 | 0.238 | 0.845 | 1.777 | | | | | | | | | | | | | | | |
| **s ~ empl_student** | -0.324 | 0.172 | -0.660 | 0.013 | | | | | | | | | | | | | | | |
| **s ~ empl_unemployed** | 0.448 | 0.157 | 0.141 | 0.756 | | | | | | | | | | | | | | | |
| **s ~ FinancialUncertainty2016** | -0.322 | 0.063 | -0.446 | -0.199 | | -0.322 | 0.062 | -0.444 | -0.199 | | -0.301 | 0.057 | -0.414 | -0.189 | | -0.316 | 0.053 | -0.420 | -0.211 |
| **s ~ FinancialHardship2017to2019_Constant_uncertainty** | 0.933 | 0.104 | 0.728 | 1.137 | | 0.998 | 0.113 | 0.775 | 1.220 | | 0.949 | 0.105 | 0.743 | 1.156 | | 0.833 | 0.095 | 0.646 | 1.020 |
| **s ~ FinancialHardship2017to2019_b+3 uncertainty** | 0.508 | 0.075 | 0.360 | 0.656 | | 0.551 | 0.076 | 0.402 | 0.700 | | 0.544 | 0.072 | 0.403 | 0.686 | | 0.538 | 0.067 | 0.408 | 0.669 |
| **s ~ FinancialHardship2017to2019_b+1 and b+3 uncertainty** | 0.437 | 0.133 | 0.177 | 0.697 | | 0.468 | 0.139 | 0.195 | 0.741 | | 0.443 | 0.125 | 0.198 | 0.688 | | 0.457 | 0.116 | 0.231 | 0.684 |
| **s ~ FinancialHardship2017to2019_b+1 uncertainty** | 0.207 | 0.068 | 0.073 | 0.341 | | 0.205 | 0.070 | 0.067 | 0.343 | | 0.184 | 0.064 | 0.058 | 0.310 | | 0.202 | 0.061 | 0.083 | 0.321 |
| **s ~ FinancialHardship2017to2019_b+1 and b+2 uncertainty** | 0.289 | 0.101 | 0.092 | 0.487 | | 0.333 | 0.104 | 0.129 | 0.536 | | 0.392 | 0.099 | 0.198 | 0.586 | | 0.335 | 0.098 | 0.144 | 0.527 |
| **s ~ FinancialHardship2017to2019_b+2 and b+3 uncertainty** | 0.689 | 0.111 | 0.471 | 0.906 | | 0.723 | 0.114 | 0.499 | 0.946 | | 0.732 | 0.102 | 0.532 | 0.932 | | 0.747 | 0.094 | 0.563 | 0.930 |
| **s ~ FinancialHardship2017to2019_b+2 uncertainty** | 0.329 | 0.071 | 0.191 | 0.468 | | 0.402 | 0.072 | 0.260 | 0.544 | | 0.393 | 0.067 | 0.262 | 0.525 | | 0.368 | 0.063 | 0.246 | 0.491 |
| **s ~ IMD_1** | -0.036 | 0.064 | -0.161 | 0.089 | | -0.012 | 0.064 | -0.137 | 0.113 | | | | | | | | | | |
| **s ~ IMD_2** | -0.038 | 0.058 | -0.152 | 0.076 | | -0.036 | 0.058 | -0.150 | 0.078 | | | | | | | | | | |
| **s ~ IMD_4** | 0.062 | 0.053 | -0.042 | 0.165 | | 0.060 | 0.053 | -0.044 | 0.163 | | | | | | | | | | |
| **s ~ IMD_5** | 0.019 | 0.051 | -0.080 | 0.119 | | 0.013 | 0.051 | -0.087 | 0.113 | | | | | | | | | | |
| **s ~ isced** | -0.165 | 0.049 | -0.261 | -0.069 | | -0.158 | 0.050 | -0.256 | -0.061 | | -0.176 | 0.047 | -0.269 | -0.084 | | | | | |
| **s ~ own_mortgage** | -0.015 | 0.047 | -0.107 | 0.077 | | -0.009 | 0.046 | -0.099 | 0.082 | | | | | | | | | | |
| **s ~ own_rent** | 0.033 | 0.057 | -0.079 | 0.145 | | 0.082 | 0.057 | -0.029 | 0.194 | | | | | | | | | | |
| **s ~ save** | -0.120 | 0.037 | -0.193 | -0.048 | | -0.144 | 0.037 | -0.216 | -0.072 | | | | | | | | | | |
| **s ~ sex_female** | -0.056 | 0.036 | -0.127 | 0.015 | | -0.067 | 0.036 | -0.138 | 0.004 | | -0.059 | 0.033 | -0.124 | 0.006 | | | | | |
| **s ~ single** | -0.043 | 0.047 | -0.135 | 0.050 | | -0.015 | 0.047 | -0.108 | 0.077 | | 0.019 | 0.043 | -0.065 | 0.103 | | | | | |
| **s ~ singleandchildren** | -0.019 | 0.105 | -0.224 | 0.186 | | -0.051 | 0.104 | -0.255 | 0.152 | | 0.028 | 0.094 | -0.156 | 0.212 | | | | | |
| **s ~~ s** | 0.714 | 0.087 | 0.542 | 0.885 | | 0.743 | 0.089 | 0.569 | 0.918 | | 0.767 | 0.082 | 0.606 | 0.929 | | 0.750 | 0.077 | 0.599 | 0.900 |

### Financial uncertainty (cross-sectional baseline weight and ipw for attrition)

| | Employment adjustment | | | | | Socio-economic adjustment | | | | | Demographic adjustment | | | | | Unadjusted | | | | |
|---|---|---|---|---|---|---|---|---|---|---|---|---|---|---|---|---|---|---|---|---|
| **term** | **Estimate** | **SE** | **CI lower** | **CI upper** | | **Estimate** | **SE** | **CI lower** | **CI upper** | | **Estimate** | **SE** | **CI lower** | **CI upper** | | **Estimate** | **SE** | **CI lower** | **CI upper** | **model.pit1** |
| **ghq_16 ~~ ghq_16** | 11.885 | 0.501 | 10.903 | 12.867 | | 11.913 | 0.505 | 10.923 | 12.902 | | 11.923 | 0.508 | 10.927 | 12.918 | | 11.953 | 0.511 | 10.951 | 12.956 | model5w |
| **ghq_17 ~~ ghq_17** | 13.164 | 0.416 | 12.349 | 13.980 | | 13.129 | 0.415 | 12.315 | 13.943 | | 13.113 | 0.417 | 12.295 | 13.931 | | 13.084 | 0.419 | 12.263 | 13.905 | model5w |

| | | | | | | | | | | | | | | | | | | | | |
|---|---|---|---|---|---|---|---|---|---|---|---|---|---|---|---|---|---|---|---|---|
| **ghq_18 ~~ ghq_18** | 13.641 | 0.425 | 12.808 | 14.474 | | 13.653 | 0.427 | 12.817 | 14.489 | | 13.656 | 0.430 | 12.813 | 14.499 | | 13.659 | 0.433 | 12.810 | 14.507 | model5w |
| **ghq_19 ~~ ghq_19** | 11.717 | 0.508 | 10.721 | 12.713 | | 11.731 | 0.516 | 10.720 | 12.742 | | 11.749 | 0.525 | 10.720 | 12.778 | | 11.763 | 0.537 | 10.711 | 12.815 | model5w |
| **i ~ age** | -0.007 | 0.005 | -0.017 | 0.003 | | -0.014 | 0.004 | -0.022 | -0.007 | | -0.023 | 0.003 | -0.030 | -0.017 | | | | | | |
| **i ~ country_LondonandSouthEngland** | -0.048 | 0.103 | -0.249 | 0.154 | | -0.061 | 0.106 | -0.267 | 0.146 | | -0.112 | 0.107 | -0.322 | 0.097 | | | | | | |
| **i ~ coupleandchildren** | -0.218 | 0.138 | -0.488 | 0.052 | | -0.249 | 0.139 | -0.521 | 0.024 | | -0.049 | 0.135 | -0.313 | 0.215 | | | | | | |
| **i ~ empl_16_maternity** | 0.635 | 0.288 | 0.071 | 1.198 | | | | | | | | | | | | | | | | |
| **i ~ empl_16_retired** | -0.456 | 0.153 | -0.756 | -0.157 | | | | | | | | | | | | | | | | |
| **i ~ empl_16_self** | -0.151 | 0.163 | -0.470 | 0.169 | | | | | | | | | | | | | | | | |
| **i ~ empl_16_sick** | 5.589 | 0.504 | 4.601 | 6.577 | | | | | | | | | | | | | | | | |
| **i ~ empl_16_student** | 0.134 | 0.285 | -0.424 | 0.693 | | | | | | | | | | | | | | | | |
| **i ~ empl_16_unemployed** | 1.305 | 0.387 | 0.546 | 2.064 | | | | | | | | | | | | | | | | |
| **i ~ empl_maternity** | 0.067 | 0.330 | -0.580 | 0.714 | | 0.376 | 0.317 | -0.244 | 0.997 | | 0.682 | 0.315 | 0.064 | 1.300 | | | | | | |
| **i ~ FinancialUncertainty2016** | 1.741 | 0.182 | 1.383 | 2.098 | | 1.927 | 0.189 | 1.556 | 2.298 | | 1.968 | 0.193 | 1.589 | 2.347 | | 1.965 | 0.198 | 1.577 | 2.353 | model5w |
| **i ~ IMD_1** | 0.075 | 0.188 | -0.294 | 0.445 | | 0.289 | 0.194 | -0.092 | 0.670 | | | | | | | | | | | |
| **i ~ IMD_2** | 0.116 | 0.167 | -0.210 | 0.443 | | 0.134 | 0.169 | -0.197 | 0.466 | | | | | | | | | | | |
| **i ~ IMD_4** | -0.178 | 0.145 | -0.461 | 0.106 | | -0.214 | 0.149 | -0.506 | 0.077 | | | | | | | | | | | |
| **i ~ IMD_5** | -0.251 | 0.140 | -0.526 | 0.023 | | -0.260 | 0.142 | -0.540 | 0.019 | | | | | | | | | | | |
| **i ~ isced** | 0.268 | 0.137 | 0.000 | 0.536 | | 0.183 | 0.138 | -0.087 | 0.453 | | -0.070 | 0.139 | -0.342 | 0.201 | | | | | | |
| **i ~ own_mortgage** | 0.337 | 0.128 | 0.087 | 0.588 | | 0.466 | 0.126 | 0.218 | 0.714 | | | | | | | | | | | |
| **i ~ own_rent** | 0.922 | 0.163 | 0.601 | 1.242 | | 1.345 | 0.167 | 1.018 | 1.673 | | | | | | | | | | | |
| **i ~ save** | -0.431 | 0.103 | -0.633 | -0.229 | | -0.571 | 0.104 | -0.775 | -0.366 | | | | | | | | | | | |
| **i ~ sex_female** | 0.997 | 0.103 | 0.794 | 1.200 | | 0.970 | 0.106 | 0.762 | 1.178 | | 0.969 | 0.107 | 0.759 | 1.179 | | | | | | |
| **i ~ single** | 0.449 | 0.135 | 0.184 | 0.714 | | 0.654 | 0.141 | 0.377 | 0.931 | | 1.108 | 0.145 | 0.825 | 1.392 | | | | | | |
| **i ~ singleandchildren** | 0.389 | 0.300 | -0.200 | 0.978 | | 0.316 | 0.302 | -0.276 | 0.908 | | 0.964 | 0.298 | 0.380 | 1.548 | | | | | | |
| **i ~~ i** | 14.491 | 0.538 | 13.437 | 15.545 | | 15.643 | 0.574 | 14.517 | 16.769 | | 16.098 | 0.596 | 14.930 | 17.267 | | 16.819 | 0.620 | 15.604 | 18.034 | model5w |
| **i ~~ s** | -1.060 | 0.173 | -1.400 | -0.720 | | -1.073 | 0.177 | -1.421 | -0.726 | | -1.028 | 0.180 | -1.380 | -0.677 | | -0.973 | 0.183 | -1.331 | -0.615 | model5w |
| **s ~ age** | -0.008 | 0.002 | -0.012 | -0.005 | | -0.008 | 0.001 | -0.011 | -0.005 | | -0.008 | 0.001 | -0.010 | -0.005 | | | | | | |
| **s ~ country_LondonandSouthEngland** | 0.071 | 0.038 | -0.004 | 0.146 | | 0.064 | 0.039 | -0.011 | 0.140 | | 0.062 | 0.038 | -0.013 | 0.137 | | | | | | |
| **s ~ coupleandchildren** | 0.079 | 0.053 | -0.025 | 0.182 | | 0.053 | 0.053 | -0.050 | 0.156 | | 0.067 | 0.051 | -0.032 | 0.167 | | | | | | |
| **s ~ empl_17_maternity** | -0.519 | 0.128 | -0.769 | -0.269 | | | | | | | | | | | | | | | | |
| **s ~ empl_17_retired** | 0.076 | 0.092 | -0.105 | 0.256 | | | | | | | | | | | | | | | | |
| **s ~ empl_17_self** | -0.063 | 0.098 | -0.256 | 0.129 | | | | | | | | | | | | | | | | |
| **s ~ empl_17_sick** | -0.816 | 0.228 | -1.263 | -0.368 | | | | | | | | | | | | | | | | |
| **s ~ empl_17_student** | -0.066 | 0.147 | -0.353 | 0.221 | | | | | | | | | | | | | | | | |
| **s ~ empl_17_unemployed** | -0.519 | 0.147 | -0.807 | -0.231 | | | | | | | | | | | | | | | | |
| **s ~ empl_18_maternity** | 0.313 | 0.134 | 0.049 | 0.576 | | | | | | | | | | | | | | | | |
| **s ~ empl_18_retired** | 0.045 | 0.103 | -0.156 | 0.246 | | | | | | | | | | | | | | | | |
| **s ~ empl_18_self** | 0.114 | 0.129 | -0.139 | 0.366 | | | | | | | | | | | | | | | | |
| **s ~ empl_18_sick** | 0.017 | 0.242 | -0.458 | 0.492 | | | | | | | | | | | | | | | | |
| **s ~ empl_18_student** | 0.163 | 0.188 | -0.206 | 0.532 | | | | | | | | | | | | | | | | |
| **s ~ empl_18_unemployed** | 0.578 | 0.176 | 0.232 | 0.923 | | | | | | | | | | | | | | | | |
| **s ~ empl_maternity** | 0.080 | 0.136 | -0.187 | 0.348 | | -0.047 | 0.106 | -0.254 | 0.161 | | -0.008 | 0.105 | -0.215 | 0.199 | | | | | | |
| **s ~ empl_retired** | -0.074 | 0.088 | -0.247 | 0.099 | | | | | | | | | | | | | | | | |
| **s ~ empl_self** | -0.042 | 0.106 | -0.249 | 0.166 | | | | | | | | | | | | | | | | |
| **s ~ empl_sick** | 1.364 | 0.244 | 0.885 | 1.843 | | | | | | | | | | | | | | | | |
| **s ~ empl_student** | -0.350 | 0.176 | -0.696 | -0.004 | | | | | | | | | | | | | | | | |
| **s ~ empl_unemployed** | 0.442 | 0.161 | 0.126 | 0.759 | | | | | | | | | | | | | | | | |
| **s ~ FinancialUncertainty2016** | -0.327 | 0.065 | -0.454 | -0.200 | | -0.328 | 0.064 | -0.453 | -0.202 | | -0.317 | 0.064 | -0.443 | -0.191 | | -0.311 | 0.065 | -0.438 | -0.184 | model5w |
| **s ~ FinancialHardship2017to2019_Constant_uncertainty** | 0.955 | 0.108 | 0.743 | 1.167 | | 1.028 | 0.120 | 0.793 | 1.263 | | 1.028 | 0.123 | 0.787 | 1.269 | | 0.903 | 0.128 | 0.652 | 1.153 | model5w |
| **s ~ FinancialHardship2017to2019_b+3 uncertainty** | 0.514 | 0.078 | 0.362 | 0.666 | | 0.561 | 0.079 | 0.406 | 0.715 | | 0.553 | 0.080 | 0.396 | 0.711 | | 0.527 | 0.080 | 0.369 | 0.685 | model5w |
| **s ~ FinancialHardship2017to2019_b+1 and b+3 uncertainty** | 0.442 | 0.138 | 0.171 | 0.714 | | 0.474 | 0.145 | 0.189 | 0.758 | | 0.461 | 0.143 | 0.182 | 0.741 | | 0.448 | 0.143 | 0.167 | 0.729 | model5w |
| **s ~ FinancialHardship2017to2019_b+1 uncertainty** | 0.203 | 0.070 | 0.066 | 0.341 | | 0.201 | 0.072 | 0.060 | 0.343 | | 0.187 | 0.073 | 0.044 | 0.331 | | 0.184 | 0.073 | 0.041 | 0.327 | model5w |
| **s ~ FinancialHardship2017to2019_b+1 and b+2 uncertainty** | 0.301 | 0.103 | 0.099 | 0.504 | | 0.349 | 0.107 | 0.139 | 0.559 | | 0.355 | 0.109 | 0.142 | 0.569 | | 0.288 | 0.114 | 0.064 | 0.511 | model5w |
| **s ~ FinancialHardship2017to2019_b+2 and b+3 uncertainty** | 0.681 | 0.115 | 0.455 | 0.907 | | 0.717 | 0.119 | 0.484 | 0.951 | | 0.709 | 0.117 | 0.479 | 0.939 | | 0.667 | 0.117 | 0.437 | 0.897 | model5w |
| **s ~ FinancialHardship2017to2019_b+2 uncertainty** | 0.337 | 0.073 | 0.194 | 0.479 | | 0.414 | 0.076 | 0.266 | 0.562 | | 0.418 | 0.078 | 0.265 | 0.570 | | 0.417 | 0.079 | 0.261 | 0.572 | model5w |
| **s ~ IMD_1** | -0.030 | 0.066 | -0.159 | 0.099 | | -0.004 | 0.066 | -0.133 | 0.126 | | | | | | | | | | | |
| **s ~ IMD_2** | -0.031 | 0.060 | -0.149 | 0.087 | | -0.029 | 0.061 | -0.148 | 0.090 | | | | | | | | | | | |
| **s ~ IMD_4** | 0.068 | 0.055 | -0.040 | 0.175 | | 0.065 | 0.055 | -0.042 | 0.173 | | | | | | | | | | | |
| **s ~ IMD_5** | 0.025 | 0.053 | -0.078 | 0.128 | | 0.019 | 0.053 | -0.085 | 0.122 | | | | | | | | | | | |
| **s ~ isced** | -0.171 | 0.051 | -0.270 | -0.071 | | -0.165 | 0.051 | -0.265 | -0.064 | | -0.191 | 0.051 | -0.291 | -0.091 | | | | | | |

| | | | | | | | | | | | | | | | | | | | | |
|---|---|---|---|---|---|---|---|---|---|---|---|---|---|---|---|---|---|---|---|---|
| **s ~ own_mortgage** | -0.017 | 0.048 | -0.112 | 0.078 | | -0.011 | 0.047 | -0.104 | 0.082 | | | | | | | | | | | |
| **s ~ own_rent** | 0.031 | 0.059 | -0.084 | 0.146 | | 0.083 | 0.058 | -0.031 | 0.197 | | | | | | | | | | | |
| **s ~ save** | -0.127 | 0.039 | -0.203 | -0.052 | | -0.153 | 0.038 | -0.227 | -0.078 | | | | | | | | | | | |
| **s ~ sex_female** | -0.064 | 0.038 | -0.138 | 0.010 | | -0.077 | 0.038 | -0.151 | -0.003 | | -0.074 | 0.038 | -0.148 | 0.000 | | | | | | |
| **s ~ single** | -0.035 | 0.049 | -0.132 | 0.061 | | -0.005 | 0.049 | -0.101 | 0.091 | | 0.027 | 0.049 | -0.069 | 0.123 | | | | | | |
| **s ~ singleandchildren** | -0.013 | 0.107 | -0.222 | 0.197 | | -0.048 | 0.106 | -0.256 | 0.160 | | 0.005 | 0.105 | -0.200 | 0.210 | | | | | | |
| **s ~~ s** | 0.729 | 0.092 | 0.548 | 0.910 | | 0.762 | 0.095 | 0.576 | 0.947 | | 0.767 | 0.095 | 0.580 | 0.955 | | 0.785 | 0.097 | 0.595 | 0.976 | model5w |

### Financial hardship and uncertainty (cross-sectional baseline)

| | Employment adjustment | | | |
|---|---|---|---|---|
| **term** | **Estimate** | **SE** | **CI lower** | **CI upper** |
| **ghq_16 ~~ ghq_16** | 11.604 | 0.473 | 10.678 | 12.531 |
| **ghq_17 ~~ ghq_17** | 12.832 | 0.389 | 12.069 | 13.596 |
| **ghq_18 ~~ ghq_18** | 13.229 | 0.397 | 12.451 | 14.006 |
| **ghq_19 ~~ ghq_19** | 11.437 | 0.479 | 10.499 | 12.376 |
| **i ~ age** | -0.012 | 0.005 | -0.022 | -0.003 |
| **i ~ country_LondonandSouthEngland** | -0.076 | 0.097 | -0.266 | 0.115 |
| **i ~ coupleandchildren** | -0.329 | 0.131 | -0.586 | -0.072 |
| **i ~ empl_16_maternity** | 0.467 | 0.272 | -0.066 | 1.000 |
| **i ~ empl_16_retired** | -0.225 | 0.145 | -0.509 | 0.059 |
| **i ~ empl_16_self** | -0.217 | 0.154 | -0.519 | 0.084 |
| **i ~ empl_16_sick** | 5.020 | 0.493 | 4.054 | 5.987 |
| **i ~ empl_16_student** | 0.226 | 0.267 | -0.298 | 0.749 |
| **i ~ empl_16_unemployed** | 0.846 | 0.365 | 0.130 | 1.562 |
| **i ~ empl_maternity** | 0.122 | 0.315 | -0.495 | 0.739 |
| **i ~ FinancialUncertainty2016** | 1.572 | 0.176 | 1.228 | 1.917 |
| **i ~ FinancialHardship2016** | 2.314 | 0.141 | 2.037 | 2.590 |
| **i ~ IMD_1** | -0.057 | 0.180 | -0.410 | 0.296 |
| **i ~ IMD_2** | 0.066 | 0.156 | -0.241 | 0.372 |
| **i ~ IMD_4** | -0.143 | 0.137 | -0.412 | 0.126 |
| **i ~ IMD_5** | -0.174 | 0.134 | -0.437 | 0.088 |
| **i ~ isced** | 0.379 | 0.131 | 0.121 | 0.636 |
| **i ~ own_mortgage** | 0.166 | 0.123 | -0.075 | 0.407 |
| **i ~ own_rent** | 0.431 | 0.158 | 0.122 | 0.740 |
| **i ~ save** | -0.092 | 0.098 | -0.284 | 0.101 |
| **i ~ sex_female** | 0.994 | 0.097 | 0.803 | 1.185 |
| **i ~ single** | 0.305 | 0.128 | 0.054 | 0.556 |
| **i ~ singleandchildren** | 0.150 | 0.292 | -0.423 | 0.722 |
| **i ~~ i** | 13.449 | 0.492 | 12.485 | 14.414 |
| **i ~~ s** | -1.037 | 0.161 | -1.352 | -0.722 |
| **s ~ age** | -0.006 | 0.002 | -0.010 | -0.003 |
| **s ~ country_LondonandSouthEngland** | 0.077 | 0.036 | 0.006 | 0.148 |
| **s ~ coupleandchildren** | 0.063 | 0.051 | -0.037 | 0.164 |
| **s ~ empl_17_maternity** | -0.465 | 0.121 | -0.702 | -0.228 |
| **s ~ empl_17_retired** | 0.047 | 0.089 | -0.128 | 0.221 |
| **s ~ empl_17_self** | -0.068 | 0.091 | -0.248 | 0.111 |
| **s ~ empl_17_sick** | -0.764 | 0.216 | -1.187 | -0.340 |
| **s ~ empl_17_student** | -0.067 | 0.139 | -0.340 | 0.206 |
| **s ~ empl_17_unemployed** | -0.484 | 0.137 | -0.752 | -0.216 |
| **s ~ empl_18_maternity** | 0.290 | 0.128 | 0.040 | 0.540 |
| **s ~ empl_18_retired** | 0.051 | 0.098 | -0.142 | 0.244 |
| **s ~ empl_18_self** | 0.092 | 0.120 | -0.143 | 0.328 |
| **s ~ empl_18_sick** | -0.008 | 0.228 | -0.454 | 0.438 |
| **s ~ empl_18_student** | 0.151 | 0.175 | -0.193 | 0.495 |
| **s ~ empl_18_unemployed** | 0.509 | 0.162 | 0.191 | 0.826 |
| **s ~ empl_maternity** | 0.068 | 0.134 | -0.193 | 0.330 |
| **s ~ empl_retired** | -0.045 | 0.085 | -0.211 | 0.120 |
| **s ~ empl_self** | -0.020 | 0.102 | -0.220 | 0.181 |

| | | | | |
|---|---|---|---|---|
| **s ~ empl_sick** | 1.327 | 0.228 | 0.879 | 1.775 |
| **s ~ empl_student** | -0.348 | 0.164 | -0.669 | -0.027 |
| **s ~ empl_unemployed** | 0.343 | 0.157 | 0.036 | 0.651 |
| **s ~ FinancialUncertainty2016** | -0.314 | 0.064 | -0.440 | -0.189 |
| **s ~ FinancialHardship2017to2019_Constant_uncertainty** | 0.795 | 0.103 | 0.592 | 0.998 |
| **s ~ FinancialHardship2017to2019_b+3 uncertainty** | 0.431 | 0.074 | 0.287 | 0.576 |
| **s ~ FinancialHardship2017to2019_b+1 and b+3 uncertainty** | 0.344 | 0.133 | 0.083 | 0.605 |
| **s ~ FinancialHardship2017to2019_b+1 uncertainty** | 0.156 | 0.068 | 0.024 | 0.289 |
| **s ~ FinancialHardship2017to2019_b+1 and b+2 uncertainty** | 0.202 | 0.103 | 0.000 | 0.403 |
| **s ~ FinancialHardship2017to2019_b+2 and b+3 uncertainty** | 0.556 | 0.109 | 0.343 | 0.769 |
| **s ~ FinancialHardship2017to2019_b+2 uncertainty** | 0.246 | 0.068 | 0.112 | 0.380 |
| **s ~ FinancialHardship2016** | -0.673 | 0.058 | -0.787 | -0.560 |
| **s ~ FinancialHardship2017to2019_Constant_hardship** | 0.784 | 0.067 | 0.653 | 0.915 |
| **s ~ FinancialHardship2017to2019_b+3 hardship** | 0.688 | 0.087 | 0.518 | 0.858 |
| **s ~ FinancialHardship2017to2019_b+1 and b+3 hardship** | 0.460 | 0.110 | 0.243 | 0.676 |
| **s ~ FinancialHardship2017to2019_b+1 hardship** | 0.116 | 0.067 | -0.015 | 0.247 |
| **s ~ FinancialHardship2017to2019_b+1 and b+2 hardship** | 0.260 | 0.093 | 0.078 | 0.443 |
| **s ~ FinancialHardship2017to2019_b+2 and b+3 hardship** | 0.748 | 0.084 | 0.583 | 0.913 |
| **s ~ FinancialHardship2017to2019_b+2 hardship** | 0.351 | 0.072 | 0.210 | 0.493 |
| **s ~ IMD_1** | -0.019 | 0.063 | -0.143 | 0.105 |
| **s ~ IMD_2** | -0.039 | 0.057 | -0.152 | 0.073 |
| **s ~ IMD_4** | 0.074 | 0.052 | -0.029 | 0.176 |
| **s ~ IMD_5** | 0.033 | 0.050 | -0.066 | 0.131 |
| **s ~ isced** | -0.158 | 0.049 | -0.254 | -0.062 |
| **s ~ own_mortgage** | -0.020 | 0.047 | -0.112 | 0.072 |
| **s ~ own_rent** | 0.041 | 0.059 | -0.074 | 0.156 |
| **s ~ save** | -0.059 | 0.037 | -0.131 | 0.014 |
| **s ~ sex_female** | -0.059 | 0.036 | -0.129 | 0.012 |
| **s ~ single** | -0.050 | 0.047 | -0.142 | 0.042 |
| **s ~ singleandchildren** | -0.060 | 0.105 | -0.266 | 0.146 |
| **s ~~ s** | 0.673 | 0.086 | 0.506 | 0.841 |

### Financial hardship and uncertainty (cross-sectional baseline and ipw for attrition)

| | Employment adjustment | | | |
|---|---|---|---|---|
| **term** | **Estimate** | **SE** | **CI lower** | **CI upper** |
| **ghq_16 ~~ ghq_16** | 11.845 | 0.496 | 10.873 | 12.817 |
| **ghq_17 ~~ ghq_17** | 13.175 | 0.412 | 12.368 | 13.982 |
| **ghq_18 ~~ ghq_18** | 13.607 | 0.421 | 12.782 | 14.432 |
| **ghq_19 ~~ ghq_19** | 11.774 | 0.505 | 10.785 | 12.763 |
| **i ~ age** | -0.012 | 0.005 | -0.022 | -0.002 |
| **i ~ country_LondonandSouthEngland** | -0.080 | 0.101 | -0.277 | 0.117 |
| **i ~ coupleandchildren** | -0.333 | 0.135 | -0.597 | -0.069 |
| **i ~ empl_16_maternity** | 0.480 | 0.281 | -0.071 | 1.032 |
| **i ~ empl_16_retired** | -0.241 | 0.148 | -0.531 | 0.050 |
| **i ~ empl_16_self** | -0.208 | 0.160 | -0.521 | 0.106 |
| **i ~ empl_16_sick** | 5.048 | 0.494 | 4.080 | 6.016 |
| **i ~ empl_16_student** | 0.203 | 0.272 | -0.330 | 0.737 |
| **i ~ empl_16_unemployed** | 0.851 | 0.381 | 0.104 | 1.597 |
| **i ~ empl_maternity** | 0.108 | 0.325 | -0.529 | 0.744 |
| **i ~ FinancialUncertainty2016** | 1.593 | 0.182 | 1.237 | 1.950 |
| **i ~ FinancialHardship2016** | 2.334 | 0.145 | 2.050 | 2.619 |
| **i ~ IMD_1** | -0.072 | 0.186 | -0.437 | 0.292 |
| **i ~ IMD_2** | 0.066 | 0.162 | -0.251 | 0.384 |
| **i ~ IMD_4** | -0.170 | 0.141 | -0.447 | 0.107 |
| **i ~ IMD_5** | -0.187 | 0.138 | -0.457 | 0.084 |
| **i ~ isced** | 0.382 | 0.135 | 0.118 | 0.646 |
| **i ~ own_mortgage** | 0.154 | 0.126 | -0.093 | 0.400 |
| **i ~ own_rent** | 0.427 | 0.160 | 0.113 | 0.741 |

| | | | | |
|---|---|---|---|---|
| **i ~ save** | -0.090 | 0.102 | -0.289 | 0.109 |
| **i ~ sex_female** | 1.016 | 0.101 | 0.817 | 1.214 |
| **i ~ single** | 0.301 | 0.133 | 0.041 | 0.561 |
| **i ~ singleandchildren** | 0.108 | 0.300 | -0.479 | 0.695 |
| **i ~~ i** | 13.697 | 0.518 | 12.682 | 14.711 |
| **i ~~ s** | -1.045 | 0.169 | -1.376 | -0.715 |
| **s ~ age** | -0.007 | 0.002 | -0.011 | -0.004 |
| **s ~ country_LondonandSouthEngland** | 0.079 | 0.038 | 0.005 | 0.153 |
| **s ~ coupleandchildren** | 0.060 | 0.053 | -0.043 | 0.164 |
| **s ~ empl_17_maternity** | -0.492 | 0.126 | -0.738 | -0.245 |
| **s ~ empl_17_retired** | 0.044 | 0.091 | -0.135 | 0.223 |
| **s ~ empl_17_self** | -0.077 | 0.096 | -0.264 | 0.111 |
| **s ~ empl_17_sick** | -0.800 | 0.225 | -1.240 | -0.360 |
| **s ~ empl_17_student** | -0.067 | 0.143 | -0.347 | 0.212 |
| **s ~ empl_17_unemployed** | -0.482 | 0.145 | -0.765 | -0.198 |
| **s ~ empl_18_maternity** | 0.303 | 0.132 | 0.044 | 0.562 |
| **s ~ empl_18_retired** | 0.058 | 0.101 | -0.140 | 0.256 |
| **s ~ empl_18_self** | 0.100 | 0.128 | -0.150 | 0.350 |
| **s ~ empl_18_sick** | -0.011 | 0.235 | -0.472 | 0.451 |
| **s ~ empl_18_student** | 0.157 | 0.180 | -0.196 | 0.510 |
| **s ~ empl_18_unemployed** | 0.528 | 0.172 | 0.191 | 0.864 |
| **s ~ empl_maternity** | 0.090 | 0.138 | -0.180 | 0.359 |
| **s ~ empl_retired** | -0.039 | 0.087 | -0.208 | 0.131 |
| **s ~ empl_self** | -0.019 | 0.107 | -0.228 | 0.191 |
| **s ~ empl_sick** | 1.378 | 0.234 | 0.918 | 1.838 |
| **s ~ empl_student** | -0.362 | 0.167 | -0.690 | -0.035 |
| **s ~ empl_unemployed** | 0.337 | 0.161 | 0.021 | 0.653 |
| **s ~ FinancialUncertainty2016** | -0.321 | 0.066 | -0.450 | -0.192 |
| **s ~ FinancialHardship2017to2019_Constant_uncertainty** | 0.813 | 0.107 | 0.604 | 1.023 |
| **s ~ FinancialHardship2017to2019_b+3 uncertainty** | 0.436 | 0.076 | 0.287 | 0.584 |
| **s ~ FinancialHardship2017to2019_b+1 and b+3 uncertainty** | 0.346 | 0.139 | 0.073 | 0.618 |
| **s ~ FinancialHardship2017to2019_b+1 uncertainty** | 0.154 | 0.069 | 0.018 | 0.289 |
| **s ~ FinancialHardship2017to2019_b+1 and b+2 uncertainty** | 0.216 | 0.106 | 0.008 | 0.423 |
| **s ~ FinancialHardship2017to2019_b+2 and b+3 uncertainty** | 0.544 | 0.113 | 0.322 | 0.766 |
| **s ~ FinancialHardship2017to2019_b+2 uncertainty** | 0.249 | 0.070 | 0.111 | 0.387 |
| **s ~ FinancialHardship2016** | -0.677 | 0.060 | -0.794 | -0.560 |
| **s ~ FinancialHardship2017to2019_Constant_hardship** | 0.802 | 0.069 | 0.668 | 0.937 |
| **s ~ FinancialHardship2017to2019_b+3 hardship** | 0.703 | 0.089 | 0.528 | 0.878 |
| **s ~ FinancialHardship2017to2019_b+1 and b+3 hardship** | 0.474 | 0.115 | 0.249 | 0.698 |
| **s ~ FinancialHardship2017to2019_b+1 hardship** | 0.111 | 0.068 | -0.022 | 0.244 |
| **s ~ FinancialHardship2017to2019_b+1 and b+2 hardship** | 0.258 | 0.097 | 0.069 | 0.447 |
| **s ~ FinancialHardship2017to2019_b+2 and b+3 hardship** | 0.756 | 0.087 | 0.586 | 0.926 |
| **s ~ FinancialHardship2017to2019_b+2 hardship** | 0.350 | 0.074 | 0.204 | 0.496 |
| **s ~ IMD_1** | -0.011 | 0.065 | -0.140 | 0.117 |
| **s ~ IMD_2** | -0.030 | 0.059 | -0.146 | 0.087 |
| **s ~ IMD_4** | 0.083 | 0.054 | -0.023 | 0.188 |
| **s ~ IMD_5** | 0.041 | 0.052 | -0.061 | 0.143 |
| **s ~ isced** | -0.163 | 0.051 | -0.262 | -0.064 |
| **s ~ own_mortgage** | -0.023 | 0.048 | -0.117 | 0.071 |
| **s ~ own_rent** | 0.040 | 0.060 | -0.078 | 0.158 |
| **s ~ save** | -0.062 | 0.038 | -0.137 | 0.014 |
| **s ~ sex_female** | -0.065 | 0.037 | -0.138 | 0.009 |
| **s ~ single** | -0.044 | 0.049 | -0.140 | 0.051 |
| **s ~ singleandchildren** | -0.055 | 0.107 | -0.265 | 0.156 |
| **s ~~ s** | 0.685 | 0.090 | 0.508 | 0.862 |

### Supplementary file S.6. LGM results for cohort 2 (2019-2022)

#### Financial hardship (cross-sectional baseline weight)

| | Employment adjustment | | | | Socio-economic adjustment | | | | Demographic adjustment | | | | Unadjusted | | | |
|---|---|---|---|---|---|---|---|---|---|---|---|---|---|---|---|---|
| **term** | **Estimate** | **SE** | **CI lower** | **CI upper** | **Estimate** | **SE** | **CI lower** | **CI upper** | **Estimate** | **SE** | **CI lower** | **CI upper** | **Estimate** | **SE** | **CI lower** | **CI upper** |
| **ghq ~~ ghq** | 10.953 | 0.483 | 10.006 | 11.900 | 11.185 | 0.499 | 10.208 | 12.162 | 11.334 | 0.455 | 10.442 | 12.226 | 11.201 | 0.426 | 10.365 | 12.036 |
| **ghq_20 ~~ ghq_20** | 14.117 | 0.425 | 13.283 | 14.950 | 14.043 | 0.426 | 13.208 | 14.878 | 13.983 | 0.383 | 13.232 | 14.734 | 13.685 | 0.361 | 12.979 | 14.392 |
| **ghq_21 ~~ ghq_21** | 14.240 | 0.393 | 13.469 | 15.011 | 14.152 | 0.394 | 13.379 | 14.925 | 14.317 | 0.390 | 13.552 | 15.081 | 14.285 | 0.365 | 13.571 | 15.000 |
| **ghq_22 ~~ ghq_22** | 10.961 | 0.501 | 9.978 | 11.943 | 11.186 | 0.510 | 10.187 | 12.185 | 11.566 | 0.468 | 10.648 | 12.484 | 11.292 | 0.435 | 10.439 | 12.146 |
| **i ~ age** | -0.034 | 0.005 | -0.043 | -0.024 | -0.036 | 0.004 | -0.043 | -0.028 | -0.037 | 0.003 | -0.043 | -0.031 | | | | |
| **i ~ country_LondonandSouthEngland** | 0.166 | 0.103 | -0.036 | 0.368 | 0.140 | 0.106 | -0.068 | 0.348 | 0.182 | 0.100 | -0.014 | 0.378 | | | | |
| **i ~ coupleandchildren** | -0.310 | 0.133 | -0.571 | -0.049 | -0.384 | 0.136 | -0.651 | -0.117 | -0.380 | 0.120 | -0.615 | -0.144 | | | | |
| **i ~ empl_maternity** | 0.589 | 0.328 | -0.055 | 1.233 | 0.324 | 0.325 | -0.313 | 0.962 | 0.470 | 0.299 | -0.117 | 1.056 | | | | |
| **i ~ empl_retired** | -0.101 | 0.147 | -0.389 | 0.187 | | | | | | | | | | | | |
| **i ~ empl_self** | -0.203 | 0.180 | -0.556 | 0.151 | | | | | | | | | | | | |
| **i ~ empl_sick** | 5.469 | 0.480 | 4.528 | 6.410 | | | | | | | | | | | | |
| **i ~ empl_student** | -0.064 | 0.268 | -0.590 | 0.462 | | | | | | | | | | | | |
| **i ~ empl_unemployed** | 1.359 | 0.371 | 0.632 | 2.085 | | | | | | | | | | | | |
| **i ~ FinancialHardship2019** | 2.939 | 0.143 | 2.658 | 3.220 | 3.236 | 0.146 | 2.950 | 3.523 | 3.475 | 0.131 | 3.219 | 3.731 | 3.682 | 0.122 | 3.443 | 3.921 |
| **i ~ IMD_1** | -0.229 | 0.184 | -0.591 | 0.133 | -0.068 | 0.194 | -0.447 | 0.312 | | | | | | | | |
| **i ~ IMD_2** | 0.129 | 0.162 | -0.189 | 0.447 | 0.173 | 0.167 | -0.154 | 0.499 | | | | | | | | |
| **i ~ IMD_4** | 0.091 | 0.143 | -0.191 | 0.372 | 0.103 | 0.147 | -0.185 | 0.390 | | | | | | | | |
| **i ~ IMD_5** | 0.048 | 0.138 | -0.222 | 0.319 | 0.058 | 0.140 | -0.217 | 0.333 | | | | | | | | |
| **i ~ isced** | 0.120 | 0.135 | -0.145 | 0.385 | 0.083 | 0.136 | -0.183 | 0.349 | -0.067 | 0.125 | -0.312 | 0.179 | | | | |
| **i ~ own_mortgage** | -0.024 | 0.129 | -0.276 | 0.229 | 0.008 | 0.128 | -0.243 | 0.258 | | | | | | | | |
| **i ~ own_rent** | 0.444 | 0.167 | 0.116 | 0.772 | 0.796 | 0.171 | 0.460 | 1.132 | | | | | | | | |
| **i ~ save** | -0.093 | 0.102 | -0.294 | 0.108 | -0.188 | 0.103 | -0.391 | 0.014 | | | | | | | | |
| **i ~ sex_female** | 1.004 | 0.101 | 0.806 | 1.201 | 1.003 | 0.103 | 0.800 | 1.205 | 1.065 | 0.094 | 0.880 | 1.250 | | | | |
| **i ~ single** | 0.132 | 0.139 | -0.140 | 0.405 | 0.273 | 0.144 | -0.008 | 0.554 | 0.490 | 0.129 | 0.237 | 0.742 | | | | |
| **i ~ singleandchildren** | -0.248 | 0.306 | -0.847 | 0.352 | -0.204 | 0.317 | -0.826 | 0.418 | 0.024 | 0.280 | -0.524 | 0.573 | | | | |
| **i ~~ i** | 13.344 | 0.495 | 12.374 | 14.315 | 14.292 | 0.528 | 13.258 | 15.326 | 14.425 | 0.486 | 13.472 | 15.378 | 15.307 | 0.463 | 14.400 | 16.214 |
| **i ~~ s** | -0.709 | 0.174 | -1.050 | -0.368 | -0.770 | 0.181 | -1.125 | -0.415 | -0.791 | 0.166 | -1.117 | -0.466 | -0.803 | 0.156 | -1.108 | -0.497 |
| **s ~ age** | 0.000 | 0.002 | -0.003 | 0.004 | 0.000 | 0.001 | -0.003 | 0.003 | 0.000 | 0.001 | -0.002 | 0.003 | | | | |
| **s ~ country_LondonandSouthEngland** | -0.034 | 0.039 | -0.111 | 0.043 | -0.034 | 0.040 | -0.112 | 0.044 | -0.031 | 0.037 | -0.104 | 0.042 | | | | |
| **s ~ coupleandchildren** | 0.093 | 0.052 | -0.009 | 0.196 | 0.098 | 0.052 | -0.005 | 0.201 | 0.068 | 0.047 | -0.024 | 0.160 | | | | |
| **s ~ empl_20_maternity** | -0.077 | 0.146 | -0.364 | 0.209 | | | | | | | | | | | | |
| **s ~ empl_20_retired** | 0.050 | 0.086 | -0.118 | 0.218 | | | | | | | | | | | | |
| **s ~ empl_20_self** | -0.106 | 0.128 | -0.357 | 0.145 | | | | | | | | | | | | |
| **s ~ empl_20_sick** | -0.885 | 0.218 | -1.311 | -0.458 | | | | | | | | | | | | |
| **s ~ empl_20_student** | 0.003 | 0.139 | -0.270 | 0.276 | | | | | | | | | | | | |
| **s ~ empl_20_unemployed** | -0.306 | 0.139 | -0.580 | -0.033 | | | | | | | | | | | | |
| **s ~ empl_21_maternity** | 0.190 | 0.149 | -0.103 | 0.483 | | | | | | | | | | | | |
| **s ~ empl_21_retired** | -0.007 | 0.109 | -0.220 | 0.207 | | | | | | | | | | | | |
| **s ~ empl_21_self** | 0.252 | 0.140 | -0.023 | 0.526 | | | | | | | | | | | | |
| **s ~ empl_21_sick** | -0.111 | 0.259 | -0.619 | 0.398 | | | | | | | | | | | | |
| **s ~ empl_21_student** | 0.044 | 0.188 | -0.324 | 0.411 | | | | | | | | | | | | |
| **s ~ empl_21_unemployed** | -0.067 | 0.130 | -0.322 | 0.188 | | | | | | | | | | | | |
| **s ~ empl_22_maternity** | 0.221 | 0.193 | -0.157 | 0.598 | | | | | | | | | | | | |
| **s ~ empl_22_retired** | 0.026 | 0.103 | -0.175 | 0.228 | | | | | | | | | | | | |
| **s ~ empl_22_self** | -0.093 | 0.128 | -0.343 | 0.158 | | | | | | | | | | | | |
| **s ~ empl_22_sick** | 0.940 | 0.278 | 0.395 | 1.486 | | | | | | | | | | | | |

| | | | | | | | | | | | | | | | | |
|---|---|---|---|---|---|---|---|---|---|---|---|---|---|---|---|---|
| **s ~ empl_22_student** | 0.332 | 0.191 | -0.043 | 0.707 | | | | | | | | | | | | |
| **s ~ empl_22_unemployed** | 0.653 | 0.152 | 0.355 | 0.951 | | | | | | | | | | | | |
| **s ~ empl_maternity** | -0.282 | 0.145 | -0.566 | 0.002 | -0.143 | 0.115 | -0.369 | 0.083 | -0.179 | 0.106 | -0.387 | 0.029 | | | | |
| **s ~ FinancialHardship2019** | -0.643 | 0.065 | -0.770 | -0.515 | -0.669 | 0.065 | -0.796 | -0.541 | -0.745 | 0.058 | -0.858 | -0.631 | -0.731 | 0.054 | -0.836 | -0.626 |
| **s ~ FinancialHardship2020to2022_Constant_hardship** | 0.875 | 0.076 | 0.727 | 1.024 | 0.933 | 0.077 | 0.782 | 1.084 | 0.972 | 0.071 | 0.833 | 1.110 | 0.950 | 0.066 | 0.820 | 1.080 |
| **s ~ FinancialHardship2020to2022_b+3_hardship** | 0.687 | 0.087 | 0.516 | 0.858 | 0.688 | 0.087 | 0.517 | 0.860 | 0.691 | 0.080 | 0.535 | 0.848 | 0.756 | 0.077 | 0.605 | 0.908 |
| **s ~ FinancialHardship2020to2022_b+1 and b+3_hardship** | 0.553 | 0.114 | 0.329 | 0.776 | 0.609 | 0.116 | 0.382 | 0.836 | 0.712 | 0.110 | 0.496 | 0.929 | 0.752 | 0.105 | 0.547 | 0.957 |
| **s ~ FinancialHardship2020to2022_b+1 hardship** | 0.217 | 0.081 | 0.058 | 0.376 | 0.214 | 0.080 | 0.057 | 0.370 | 0.225 | 0.071 | 0.087 | 0.363 | 0.233 | 0.065 | 0.105 | 0.361 |
| **s ~ FinancialHardship2020to2022_b+1 and b+2 hardship** | 0.259 | 0.100 | 0.064 | 0.455 | 0.223 | 0.098 | 0.030 | 0.415 | 0.242 | 0.089 | 0.068 | 0.417 | 0.309 | 0.083 | 0.147 | 0.471 |
| **s ~ FinancialHardship2020to2022_b+2 and b+3 hardship** | 1.009 | 0.119 | 0.775 | 1.242 | 1.043 | 0.119 | 0.810 | 1.277 | 1.072 | 0.112 | 0.852 | 1.292 | 1.085 | 0.105 | 0.878 | 1.291 |
| **s ~ FinancialHardship2020to2022_b+2 hardship** | 0.293 | 0.107 | 0.082 | 0.503 | 0.281 | 0.107 | 0.071 | 0.491 | 0.287 | 0.098 | 0.096 | 0.478 | 0.380 | 0.092 | 0.201 | 0.560 |
| **s ~ IMD_1** | 0.073 | 0.069 | -0.062 | 0.208 | 0.068 | 0.070 | -0.069 | 0.205 | | | | | | | | |
| **s ~ IMD_2** | 0.087 | 0.064 | -0.039 | 0.212 | 0.090 | 0.064 | -0.036 | 0.216 | | | | | | | | |
| **s ~ IMD_4** | -0.015 | 0.055 | -0.122 | 0.092 | -0.023 | 0.055 | -0.130 | 0.085 | | | | | | | | |
| **s ~ IMD_5** | -0.011 | 0.053 | -0.115 | 0.092 | -0.009 | 0.053 | -0.113 | 0.095 | | | | | | | | |
| **s ~ isced** | 0.109 | 0.056 | -0.001 | 0.220 | 0.095 | 0.057 | -0.016 | 0.206 | 0.121 | 0.052 | 0.020 | 0.223 | | | | |
| **s ~ own_mortgage** | -0.037 | 0.048 | -0.131 | 0.057 | -0.070 | 0.047 | -0.163 | 0.022 | | | | | | | | |
| **s ~ own_rent** | -0.123 | 0.064 | -0.247 | 0.002 | -0.147 | 0.063 | -0.270 | -0.024 | | | | | | | | |
| **s ~ save** | 0.123 | 0.039 | 0.046 | 0.200 | 0.118 | 0.039 | 0.041 | 0.195 | | | | | | | | |
| **s ~ sex_female** | 0.102 | 0.038 | 0.027 | 0.177 | 0.110 | 0.038 | 0.035 | 0.185 | 0.091 | 0.035 | 0.023 | 0.159 | | | | |
| **s ~ single** | -0.034 | 0.051 | -0.134 | 0.066 | -0.041 | 0.051 | -0.141 | 0.058 | -0.089 | 0.045 | -0.178 | 0.000 | | | | |
| **s ~ singleandchildren** | -0.005 | 0.119 | -0.239 | 0.228 | 0.013 | 0.119 | -0.220 | 0.247 | -0.006 | 0.107 | -0.216 | 0.204 | | | | |
| **s ~~ s** | 0.842 | 0.094 | 0.658 | 1.026 | 0.831 | 0.096 | 0.643 | 1.019 | 0.818 | 0.087 | 0.647 | 0.989 | 0.824 | 0.081 | 0.664 | 0.983 |

### Financial hardship (cross-sectional baseline weight and ipw for attrition)

| | Employment adjustment | | | | Socio-economic adjustment | | | | Demographic adjustment | | | | Unadjusted | | | |
|---|---|---|---|---|---|---|---|---|---|---|---|---|---|---|---|---|
| **term** | **Estimate** | **SE** | **CI lower** | **CI upper** | **Estimate** | **SE** | **CI lower** | **CI upper** | **Estimate** | **SE** | **CI lower** | **CI upper** | **Estimate** | **SE** | **CI lower** | **CI upper** |
| **ghq ~~ ghq** | 11.954 | 0.629 | 10.721 | 13.187 | 12.229 | 0.647 | 10.961 | 13.497 | 12.237 | 0.653 | 10.957 | 13.517 | 12.265 | 0.668 | 10.954 | 13.575 |
| **ghq_20 ~~ ghq_20** | 15.001 | 0.508 | 14.005 | 15.996 | 14.912 | 0.507 | 13.918 | 15.906 | 14.923 | 0.510 | 13.923 | 15.923 | 14.900 | 0.511 | 13.898 | 15.902 |
| **ghq_21 ~~ ghq_21** | 14.872 | 0.437 | 14.015 | 15.728 | 14.754 | 0.440 | 13.891 | 15.617 | 14.756 | 0.441 | 13.892 | 15.620 | 14.783 | 0.443 | 13.915 | 15.652 |
| **ghq_22 ~~ ghq_22** | 11.802 | 0.579 | 10.666 | 12.937 | 12.084 | 0.591 | 10.926 | 13.242 | 12.049 | 0.592 | 10.889 | 13.210 | 12.012 | 0.598 | 10.839 | 13.186 |
| **i ~ age** | -0.035 | 0.005 | -0.046 | -0.025 | -0.037 | 0.004 | -0.045 | -0.028 | -0.040 | 0.004 | -0.047 | -0.032 | | | | |
| **i ~ country_LondonandSouthEngland** | 0.188 | 0.119 | -0.046 | 0.422 | 0.155 | 0.124 | -0.087 | 0.397 | 0.175 | 0.124 | -0.068 | 0.417 | | | | |
| **i ~ coupleandchildren** | -0.341 | 0.148 | -0.631 | -0.051 | -0.415 | 0.152 | -0.713 | -0.117 | -0.405 | 0.149 | -0.696 | -0.113 | | | | |
| **i ~ empl_maternity** | 0.832 | 0.410 | 0.029 | 1.635 | 0.518 | 0.407 | -0.280 | 1.315 | 0.690 | 0.410 | -0.113 | 1.493 | | | | |
| **i ~ empl_retired** | -0.144 | 0.161 | -0.459 | 0.171 | | | | | | | | | | | | |
| **i ~ empl_self** | -0.179 | 0.202 | -0.575 | 0.217 | | | | | | | | | | | | |
| **i ~ empl_sick** | 5.557 | 0.527 | 4.523 | 6.591 | | | | | | | | | | | | |
| **i ~ empl_student** | -0.086 | 0.305 | -0.684 | 0.513 | | | | | | | | | | | | |
| **i ~ empl_unemployed** | 1.316 | 0.421 | 0.491 | 2.141 | | | | | | | | | | | | |
| **i ~ FinancialHardship2019** | 2.968 | 0.162 | 2.650 | 3.286 | 3.289 | 0.165 | 2.966 | 3.611 | 3.552 | 0.164 | 3.231 | 3.873 | 3.773 | 0.165 | 3.448 | 4.097 |
| **i ~ IMD_1** | -0.226 | 0.209 | -0.635 | 0.183 | -0.073 | 0.219 | -0.501 | 0.356 | | | | | | | | |
| **i ~ IMD_2** | 0.199 | 0.186 | -0.164 | 0.563 | 0.243 | 0.191 | -0.131 | 0.618 | | | | | | | | |
| **i ~ IMD_4** | 0.026 | 0.159 | -0.286 | 0.338 | 0.037 | 0.164 | -0.285 | 0.359 | | | | | | | | |
| **i ~ IMD_5** | 0.054 | 0.151 | -0.242 | 0.350 | 0.067 | 0.154 | -0.235 | 0.369 | | | | | | | | |
| **i ~ isced** | 0.093 | 0.145 | -0.191 | 0.378 | 0.047 | 0.146 | -0.240 | 0.333 | -0.070 | 0.147 | -0.358 | 0.218 | | | | |
| **i ~ own_mortgage** | -0.099 | 0.139 | -0.371 | 0.174 | -0.050 | 0.139 | -0.323 | 0.223 | | | | | | | | |
| **i ~ own_rent** | 0.483 | 0.177 | 0.136 | 0.831 | 0.863 | 0.181 | 0.509 | 1.218 | | | | | | | | |
| **i ~ save** | -0.148 | 0.114 | -0.372 | 0.076 | -0.249 | 0.115 | -0.475 | -0.023 | | | | | | | | |
| **i ~ sex_female** | 1.051 | 0.117 | 0.821 | 1.280 | 1.043 | 0.121 | 0.806 | 1.279 | 1.039 | 0.121 | 0.802 | 1.277 | | | | |
| **i ~ single** | 0.146 | 0.157 | -0.161 | 0.453 | 0.304 | 0.162 | -0.013 | 0.621 | 0.546 | 0.161 | 0.230 | 0.862 | | | | |
| **i ~ singleandchildren** | -0.284 | 0.345 | -0.961 | 0.393 | -0.249 | 0.360 | -0.954 | 0.456 | 0.066 | 0.358 | -0.636 | 0.768 | | | | |
| **i ~~ i** | 14.181 | 0.593 | 13.019 | 15.343 | 15.302 | 0.623 | 14.081 | 16.523 | 15.463 | 0.635 | 14.219 | 16.707 | 16.286 | 0.648 | 15.016 | 17.555 |
| **i ~~ s** | -0.774 | 0.223 | -1.210 | -0.337 | -0.836 | 0.228 | -1.283 | -0.390 | -0.860 | 0.231 | -1.313 | -0.406 | -0.855 | 0.234 | -1.313 | -0.396 |

| | | | | | | | | | | | | | | | | |
|---|---|---|---|---|---|---|---|---|---|---|---|---|---|---|---|---|
| **s ~ age** | 0.000 | 0.002 | -0.004 | 0.004 | 0.000 | 0.002 | -0.003 | 0.003 | 0.001 | 0.001 | -0.002 | 0.003 | | | | |
| **s ~ country_LondonandSouthEngland** | -0.033 | 0.046 | -0.122 | 0.057 | -0.031 | 0.046 | -0.122 | 0.059 | -0.035 | 0.046 | -0.124 | 0.055 | | | | |
| **s ~ coupleandchildren** | 0.100 | 0.058 | -0.012 | 0.213 | 0.103 | 0.058 | -0.011 | 0.216 | 0.078 | 0.056 | -0.032 | 0.189 | | | | |
| **s ~ empl_20_maternity** | -0.181 | 0.165 | -0.505 | 0.143 | | | | | | | | | | | | |
| **s ~ empl_20_retired** | 0.014 | 0.095 | -0.172 | 0.201 | | | | | | | | | | | | |
| **s ~ empl_20_self** | -0.145 | 0.147 | -0.433 | 0.143 | | | | | | | | | | | | |
| **s ~ empl_20_sick** | -1.039 | 0.243 | -1.516 | -0.562 | | | | | | | | | | | | |
| **s ~ empl_20_student** | -0.030 | 0.143 | -0.310 | 0.250 | | | | | | | | | | | | |
| **s ~ empl_20_unemployed** | -0.225 | 0.153 | -0.526 | 0.075 | | | | | | | | | | | | |
| **s ~ empl_21_maternity** | 0.209 | 0.168 | -0.119 | 0.538 | | | | | | | | | | | | |
| **s ~ empl_21_retired** | 0.040 | 0.128 | -0.211 | 0.291 | | | | | | | | | | | | |
| **s ~ empl_21_self** | 0.316 | 0.157 | 0.009 | 0.623 | | | | | | | | | | | | |
| **s ~ empl_21_sick** | -0.018 | 0.301 | -0.608 | 0.572 | | | | | | | | | | | | |
| **s ~ empl_21_student** | -0.013 | 0.192 | -0.390 | 0.364 | | | | | | | | | | | | |
| **s ~ empl_21_unemployed** | -0.055 | 0.141 | -0.331 | 0.222 | | | | | | | | | | | | |
| **s ~ empl_22_maternity** | 0.225 | 0.217 | -0.201 | 0.652 | | | | | | | | | | | | |
| **s ~ empl_22_retired** | 0.034 | 0.115 | -0.191 | 0.259 | | | | | | | | | | | | |
| **s ~ empl_22_self** | -0.122 | 0.141 | -0.398 | 0.155 | | | | | | | | | | | | |
| **s ~ empl_22_sick** | 1.045 | 0.332 | 0.394 | 1.697 | | | | | | | | | | | | |
| **s ~ empl_22_student** | 0.318 | 0.199 | -0.072 | 0.707 | | | | | | | | | | | | |
| **s ~ empl_22_unemployed** | 0.602 | 0.162 | 0.284 | 0.921 | | | | | | | | | | | | |
| **s ~ empl_maternity** | -0.330 | 0.174 | -0.671 | 0.010 | -0.228 | 0.142 | -0.506 | 0.049 | -0.263 | 0.142 | -0.542 | 0.016 | | | | |
| **s ~ FinancialHardship2019** | -0.650 | 0.072 | -0.792 | -0.508 | -0.675 | 0.072 | -0.816 | -0.534 | -0.743 | 0.071 | -0.882 | -0.604 | -0.757 | 0.071 | -0.895 | -0.619 |
| **s ~ FinancialHardship2020to2022_Constant_hardship** | 0.890 | 0.082 | 0.728 | 1.051 | 0.956 | 0.084 | 0.791 | 1.122 | 0.949 | 0.086 | 0.782 | 1.117 | 0.935 | 0.086 | 0.765 | 1.104 |
| **s ~ FinancialHardship2020to2022_b+3_hardship** | 0.731 | 0.095 | 0.545 | 0.916 | 0.727 | 0.094 | 0.542 | 0.912 | 0.725 | 0.094 | 0.540 | 0.909 | 0.771 | 0.097 | 0.581 | 0.961 |
| **s ~ FinancialHardship2020to2022_b+1 and b+3_hardship** | 0.547 | 0.124 | 0.303 | 0.791 | 0.610 | 0.125 | 0.365 | 0.854 | 0.604 | 0.126 | 0.357 | 0.850 | 0.614 | 0.125 | 0.368 | 0.860 |
| **s ~ FinancialHardship2020to2022_b+1 hardship** | 0.237 | 0.089 | 0.062 | 0.412 | 0.242 | 0.088 | 0.070 | 0.414 | 0.233 | 0.088 | 0.061 | 0.404 | 0.267 | 0.090 | 0.091 | 0.443 |
| **s ~ FinancialHardship2020to2022_b+1 and b+2 hardship** | 0.270 | 0.113 | 0.049 | 0.492 | 0.242 | 0.111 | 0.025 | 0.460 | 0.235 | 0.112 | 0.017 | 0.454 | 0.249 | 0.113 | 0.028 | 0.470 |
| **s ~ FinancialHardship2020to2022_b+2 and b+3 hardship** | 1.078 | 0.129 | 0.826 | 1.330 | 1.118 | 0.128 | 0.866 | 1.370 | 1.115 | 0.129 | 0.863 | 1.368 | 1.152 | 0.128 | 0.901 | 1.403 |
| **s ~ FinancialHardship2020to2022_b+2 hardship** | 0.274 | 0.116 | 0.047 | 0.501 | 0.256 | 0.117 | 0.027 | 0.484 | 0.245 | 0.117 | 0.016 | 0.473 | 0.300 | 0.118 | 0.068 | 0.531 |
| **s ~ IMD_1** | 0.090 | 0.075 | -0.058 | 0.238 | 0.086 | 0.077 | -0.064 | 0.236 | | | | | | | | |
| **s ~ IMD_2** | 0.082 | 0.073 | -0.061 | 0.225 | 0.089 | 0.073 | -0.054 | 0.232 | | | | | | | | |
| **s ~ IMD_4** | -0.012 | 0.060 | -0.129 | 0.106 | -0.018 | 0.060 | -0.135 | 0.100 | | | | | | | | |
| **s ~ IMD_5** | -0.001 | 0.057 | -0.112 | 0.110 | 0.002 | 0.057 | -0.109 | 0.113 | | | | | | | | |
| **s ~ isced** | 0.132 | 0.062 | 0.011 | 0.253 | 0.117 | 0.062 | -0.004 | 0.238 | 0.132 | 0.062 | 0.011 | 0.253 | | | | |
| **s ~ own_mortgage** | -0.026 | 0.051 | -0.126 | 0.073 | -0.063 | 0.051 | -0.164 | 0.037 | | | | | | | | |
| **s ~ own_rent** | -0.142 | 0.067 | -0.273 | -0.010 | -0.163 | 0.065 | -0.292 | -0.035 | | | | | | | | |
| **s ~ save** | 0.157 | 0.043 | 0.072 | 0.241 | 0.154 | 0.044 | 0.068 | 0.239 | | | | | | | | |
| **s ~ sex_female** | 0.122 | 0.044 | 0.036 | 0.208 | 0.126 | 0.044 | 0.040 | 0.212 | 0.127 | 0.044 | 0.041 | 0.212 | | | | |
| **s ~ single** | -0.058 | 0.057 | -0.169 | 0.054 | -0.066 | 0.057 | -0.177 | 0.045 | -0.092 | 0.056 | -0.202 | 0.017 | | | | |
| **s ~ singleandchildren** | -0.014 | 0.130 | -0.268 | 0.240 | -0.005 | 0.128 | -0.256 | 0.245 | -0.051 | 0.127 | -0.300 | 0.197 | | | | |
| **s ~~ s** | 0.849 | 0.118 | 0.617 | 1.080 | 0.834 | 0.119 | 0.601 | 1.067 | 0.845 | 0.120 | 0.610 | 1.079 | 0.856 | 0.121 | 0.619 | 1.092 |

### Financial uncertainty (cross-sectional baseline weight)

| | Employment adjustment | | | | Socio-economic adjustment | | | | Demographic adjustment | | | | Unadjusted | | | |
|---|---|---|---|---|---|---|---|---|---|---|---|---|---|---|---|---|
| **term** | **Estimate** | **SE** | **CI lower** | **CI upper** | **Estimate** | **SE** | **CI lower** | **CI upper** | **Estimate** | **SE** | **CI lower** | **CI upper** | **Estimate** | **SE** | **CI lower** | **CI upper** |
| **ghq ~~ ghq** | 10.867 | 0.494 | 9.899 | 11.835 | 11.074 | 0.509 | 10.077 | 12.071 | 11.271 | 0.467 | 10.356 | 12.187 | 11.116 | 0.438 | 10.257 | 11.976 |
| **ghq_20 ~~ ghq_20** | 13.989 | 0.434 | 13.139 | 14.840 | 13.922 | 0.435 | 13.069 | 14.775 | 13.822 | 0.393 | 13.052 | 14.592 | 13.472 | 0.367 | 12.753 | 14.190 |
| **ghq_21 ~~ ghq_21** | 13.974 | 0.386 | 13.218 | 14.729 | 13.880 | 0.386 | 13.123 | 14.636 | 14.007 | 0.393 | 13.237 | 14.777 | 14.005 | 0.370 | 13.280 | 14.729 |
| **ghq_22 ~~ ghq_22** | 10.989 | 0.498 | 10.012 | 11.965 | 11.246 | 0.507 | 10.253 | 12.240 | 11.730 | 0.471 | 10.806 | 12.654 | 11.438 | 0.443 | 10.571 | 12.306 |
| **i ~ age** | -0.033 | 0.005 | -0.043 | -0.023 | -0.038 | 0.004 | -0.046 | -0.030 | -0.044 | 0.003 | -0.050 | -0.037 | | | | |
| **i ~ country_LondonandSouthEngland** | 0.239 | 0.107 | 0.029 | 0.448 | 0.215 | 0.110 | -0.001 | 0.431 | 0.229 | 0.105 | 0.023 | 0.435 | | | | |

| | | | | | | | | | | | | | | | | | | | |
|---|---|---|---|---|---|---|---|---|---|---|---|---|---|---|---|---|---|---|---|
| **i ~ coupleandchildren** | -0.033 | 0.135 | -0.299 | 0.232 | | -0.074 | 0.139 | -0.346 | 0.198 | | 0.048 | 0.124 | -0.194 | 0.290 | | | | | |
| **i ~ empl_maternity** | 0.384 | 0.329 | -0.261 | 1.030 | | 0.129 | 0.325 | -0.509 | 0.766 | | 0.570 | 0.302 | -0.023 | 1.163 | | | | | |
| **i ~ empl_retired** | -0.355 | 0.151 | -0.652 | -0.059 | | | | | | | | | | | | | | | |
| **i ~ empl_self** | -0.194 | 0.185 | -0.556 | 0.168 | | | | | | | | | | | | | | | |
| **i ~ empl_sick** | 5.750 | 0.510 | 4.749 | 6.750 | | | | | | | | | | | | | | | |
| **i ~ empl_student** | -0.160 | 0.284 | -0.717 | 0.397 | | | | | | | | | | | | | | | |
| **i ~ empl_unemployed** | 1.968 | 0.403 | 1.177 | 2.758 | | | | | | | | | | | | | | | |
| **i ~ FinancialUncertainty2019** | 2.174 | 0.172 | 1.836 | 2.512 | | 2.353 | 0.179 | 2.002 | 2.705 | | 2.459 | 0.168 | 2.131 | 2.788 | | 2.429 | 0.156 | 2.122 | 2.735 |
| **i ~ IMD_1** | -0.157 | 0.192 | -0.533 | 0.218 | | 0.020 | 0.202 | -0.375 | 0.416 | | | | | | | | | | |
| **i ~ IMD_2** | 0.178 | 0.168 | -0.151 | 0.507 | | 0.217 | 0.173 | -0.122 | 0.556 | | | | | | | | | | |
| **i ~ IMD_4** | -0.039 | 0.146 | -0.325 | 0.247 | | -0.052 | 0.150 | -0.346 | 0.242 | | | | | | | | | | |
| **i ~ IMD_5** | -0.095 | 0.141 | -0.370 | 0.181 | | -0.118 | 0.143 | -0.399 | 0.163 | | | | | | | | | | |
| **i ~ isced** | -0.032 | 0.142 | -0.311 | 0.247 | | -0.066 | 0.142 | -0.345 | 0.213 | | -0.362 | 0.131 | -0.619 | -0.105 | | | | | |
| **i ~ own_mortgage** | 0.129 | 0.133 | -0.131 | 0.389 | | 0.235 | 0.132 | -0.023 | 0.493 | | | | | | | | | | |
| **i ~ own_rent** | 1.008 | 0.172 | 0.672 | 1.344 | | 1.493 | 0.178 | 1.145 | 1.841 | | | | | | | | | | |
| **i ~ save** | -0.712 | 0.105 | -0.917 | -0.506 | | -0.878 | 0.107 | -1.087 | -0.669 | | | | | | | | | | |
| **i ~ sex_female** | 1.019 | 0.104 | 0.815 | 1.224 | | 1.011 | 0.107 | 0.801 | 1.221 | | 1.056 | 0.099 | 0.862 | 1.250 | | | | | |
| **i ~ single** | 0.321 | 0.145 | 0.037 | 0.605 | | 0.484 | 0.149 | 0.191 | 0.776 | | 0.966 | 0.138 | 0.695 | 1.237 | | | | | |
| **i ~ singleandchildren** | 0.221 | 0.319 | -0.404 | 0.846 | | 0.322 | 0.333 | -0.330 | 0.974 | | 0.973 | 0.297 | 0.391 | 1.554 | | | | | |
| **i ~~ i** | 14.122 | 0.528 | 13.088 | 15.157 | | 15.285 | 0.563 | 14.182 | 16.388 | | 15.872 | 0.538 | 14.818 | 16.927 | | 17.122 | 0.518 | 16.106 | 18.137 |
| **i ~~ s** | -0.858 | 0.184 | -1.219 | -0.498 | | -0.955 | 0.191 | -1.331 | -0.580 | | -0.957 | 0.178 | -1.306 | -0.608 | | -0.888 | 0.166 | -1.213 | -0.563 |
| **s ~ age** | -0.002 | 0.002 | -0.006 | 0.002 | | -0.002 | 0.001 | -0.005 | 0.000 | | -0.002 | 0.001 | -0.004 | 0.001 | | | | | |
| **s ~ country_LondonandSouthEngland** | -0.053 | 0.040 | -0.131 | 0.025 | | -0.051 | 0.040 | -0.130 | 0.028 | | -0.053 | 0.038 | -0.128 | 0.021 | | | | | |
| **s ~ coupleandchildren** | 0.086 | 0.053 | -0.018 | 0.190 | | 0.093 | 0.053 | -0.010 | 0.197 | | 0.059 | 0.047 | -0.033 | 0.152 | | | | | |
| **s ~ empl_20_maternity** | -0.110 | 0.148 | -0.399 | 0.180 | | | | | | | | | | | | | | | |
| **s ~ empl_20_retired** | 0.096 | 0.088 | -0.077 | 0.269 | | | | | | | | | | | | | | | |
| **s ~ empl_20_self** | -0.083 | 0.129 | -0.336 | 0.169 | | | | | | | | | | | | | | | |
| **s ~ empl_20_sick** | -1.144 | 0.229 | -1.592 | -0.696 | | | | | | | | | | | | | | | |
| **s ~ empl_20_student** | -0.015 | 0.147 | -0.303 | 0.273 | | | | | | | | | | | | | | | |
| **s ~ empl_20_unemployed** | -0.311 | 0.147 | -0.598 | -0.023 | | | | | | | | | | | | | | | |
| **s ~ empl_21_maternity** | 0.191 | 0.153 | -0.108 | 0.490 | | | | | | | | | | | | | | | |
| **s ~ empl_21_retired** | 0.004 | 0.110 | -0.211 | 0.220 | | | | | | | | | | | | | | | |
| **s ~ empl_21_self** | 0.283 | 0.140 | 0.010 | 0.557 | | | | | | | | | | | | | | | |
| **s ~ empl_21_sick** | -0.073 | 0.259 | -0.581 | 0.434 | | | | | | | | | | | | | | | |
| **s ~ empl_21_student** | 0.076 | 0.195 | -0.305 | 0.458 | | | | | | | | | | | | | | | |
| **s ~ empl_21_unemployed** | -0.057 | 0.133 | -0.318 | 0.204 | | | | | | | | | | | | | | | |
| **s ~ empl_22_maternity** | 0.154 | 0.192 | -0.222 | 0.529 | | | | | | | | | | | | | | | |
| **s ~ empl_22_retired** | -0.028 | 0.102 | -0.229 | 0.173 | | | | | | | | | | | | | | | |
| **s ~ empl_22_self** | -0.104 | 0.126 | -0.351 | 0.142 | | | | | | | | | | | | | | | |
| **s ~ empl_22_sick** | 1.133 | 0.289 | 0.566 | 1.700 | | | | | | | | | | | | | | | |
| **s ~ empl_22_student** | 0.335 | 0.195 | -0.047 | 0.717 | | | | | | | | | | | | | | | |
| **s ~ empl_22_unemployed** | 0.726 | 0.154 | 0.423 | 1.028 | | | | | | | | | | | | | | | |
| **s ~ empl_maternity** | -0.196 | 0.143 | -0.476 | 0.084 | | -0.099 | 0.115 | -0.324 | 0.125 | | -0.142 | 0.105 | -0.348 | 0.064 | | | | | |
| **s ~ FinancialUncertainty2019** | -0.346 | 0.065 | -0.473 | -0.219 | | -0.349 | 0.066 | -0.478 | -0.221 | | -0.375 | 0.061 | -0.495 | -0.255 | | -0.336 | 0.056 | -0.446 | -0.226 |
| **s ~ FinancialHardship2020to2022_Constant_uncertainty** | 0.969 | 0.100 | 0.773 | 1.164 | | 1.000 | 0.103 | 0.799 | 1.201 | | 0.989 | 0.096 | 0.801 | 1.176 | | 0.838 | 0.089 | 0.663 | 1.012 |
| **s ~ FinancialHardship2020to2022_b+3_uncertainty** | 0.443 | 0.054 | 0.338 | 0.549 | | 0.465 | 0.054 | 0.359 | 0.571 | | 0.434 | 0.051 | 0.334 | 0.534 | | 0.393 | 0.049 | 0.297 | 0.490 |
| **s ~ FinancialHardship2020to2022_b+1 and b+3_uncertainty** | 0.618 | 0.098 | 0.427 | 0.810 | | 0.619 | 0.100 | 0.423 | 0.815 | | 0.582 | 0.093 | 0.401 | 0.764 | | 0.521 | 0.091 | 0.342 | 0.699 |
| **s ~ FinancialHardship2020to2022_b+1 uncertainty** | 0.287 | 0.083 | 0.125 | 0.449 | | 0.300 | 0.082 | 0.140 | 0.461 | | 0.287 | 0.075 | 0.140 | 0.434 | | 0.261 | 0.071 | 0.121 | 0.401 |

| | | | | | | | | | | | | | | | | |
|---|---|---|---|---|---|---|---|---|---|---|---|---|---|---|---|---|
| **s ~ FinancialHardship2020to2022_b+1 and b+2 uncertainty** | 0.635 | 0.132 | 0.376 | 0.893 | 0.640 | 0.134 | 0.377 | 0.903 | 0.628 | 0.121 | 0.391 | 0.865 | 0.567 | 0.117 | 0.337 | 0.796 |
| **s ~ FinancialHardship2020to2022_b+2 and b+3 uncertainty** | 0.810 | 0.104 | 0.605 | 1.014 | 0.808 | 0.105 | 0.602 | 1.015 | 0.832 | 0.099 | 0.637 | 1.027 | 0.764 | 0.093 | 0.582 | 0.945 |
| **s ~ FinancialHardship2020to2022_b+2 uncertainty** | 0.512 | 0.088 | 0.339 | 0.684 | 0.523 | 0.089 | 0.349 | 0.698 | 0.530 | 0.079 | 0.374 | 0.685 | 0.466 | 0.072 | 0.324 | 0.608 |
| **s ~ IMD_1** | 0.106 | 0.070 | -0.031 | 0.243 | 0.103 | 0.071 | -0.036 | 0.242 | | | | | | | | |
| **s ~ IMD_2** | 0.091 | 0.064 | -0.035 | 0.218 | 0.099 | 0.065 | -0.028 | 0.226 | | | | | | | | |
| **s ~ IMD_4** | -0.003 | 0.055 | -0.112 | 0.106 | -0.008 | 0.056 | -0.117 | 0.102 | | | | | | | | |
| **s ~ IMD_5** | -0.022 | 0.053 | -0.126 | 0.083 | -0.018 | 0.053 | -0.123 | 0.086 | | | | | | | | |
| **s ~ isced** | 0.091 | 0.057 | -0.021 | 0.203 | 0.075 | 0.057 | -0.037 | 0.188 | 0.090 | 0.053 | -0.013 | 0.194 | | | | |
| **s ~ own_mortgage** | -0.039 | 0.048 | -0.134 | 0.056 | -0.072 | 0.048 | -0.166 | 0.022 | | | | | | | | |
| **s ~ own_rent** | -0.078 | 0.064 | -0.204 | 0.047 | -0.102 | 0.063 | -0.226 | 0.022 | | | | | | | | |
| **s ~ save** | 0.134 | 0.038 | 0.060 | 0.209 | 0.127 | 0.038 | 0.052 | 0.202 | | | | | | | | |
| **s ~ sex_female** | 0.102 | 0.039 | 0.026 | 0.178 | 0.105 | 0.039 | 0.029 | 0.181 | 0.088 | 0.035 | 0.018 | 0.157 | | | | |
| **s ~ single** | -0.054 | 0.052 | -0.155 | 0.048 | -0.060 | 0.052 | -0.161 | 0.041 | -0.089 | 0.046 | -0.179 | 0.002 | | | | |
| **s ~ singleandchildren** | -0.037 | 0.117 | -0.267 | 0.194 | -0.014 | 0.117 | -0.243 | 0.215 | 0.010 | 0.104 | -0.194 | 0.214 | | | | |
| **s ~~ s** | 0.862 | 0.097 | 0.673 | 1.052 | 0.859 | 0.098 | 0.667 | 1.052 | 0.832 | 0.090 | 0.655 | 1.009 | 0.826 | 0.084 | 0.662 | 0.990 |

### Financial uncertainty (cross-sectional baseline weight and ipw for attrition)

| | Employment adjustment | | | | Socio-economic adjustment | | | | Demographic adjustment | | | | Unadjusted | | | |
|---|---|---|---|---|---|---|---|---|---|---|---|---|---|---|---|---|
| **term** | **Estimate** | **SE** | **CI lower** | **CI upper** | **Estimate** | **SE** | **CI lower** | **CI upper** | **Estimate** | **SE** | **CI lower** | **CI upper** | **Estimate** | **SE** | **CI lower** | **CI upper** |
| **ghq ~~ ghq** | 11.903 | 0.640 | 10.648 | 13.158 | 12.172 | 0.657 | 10.883 | 13.461 | 12.247 | 0.670 | 10.933 | 13.560 | 12.297 | 0.689 | 10.946 | 13.647 |
| **ghq_20 ~~ ghq_20** | 14.886 | 0.517 | 13.872 | 15.900 | 14.796 | 0.517 | 13.783 | 15.809 | 14.789 | 0.524 | 13.763 | 15.816 | 14.759 | 0.527 | 13.726 | 15.791 |
| **ghq_21 ~~ ghq_21** | 14.645 | 0.430 | 13.802 | 15.487 | 14.526 | 0.432 | 13.680 | 15.372 | 14.489 | 0.432 | 13.642 | 15.335 | 14.510 | 0.435 | 13.657 | 15.363 |
| **ghq_22 ~~ ghq_22** | 11.828 | 0.581 | 10.689 | 12.966 | 12.129 | 0.591 | 10.971 | 13.286 | 12.147 | 0.596 | 10.978 | 13.316 | 12.117 | 0.609 | 10.922 | 13.311 |
| **i ~ age** | -0.033 | 0.006 | -0.044 | -0.022 | -0.038 | 0.004 | -0.047 | -0.029 | -0.045 | 0.004 | -0.053 | -0.037 | | | | |
| **i ~ country_LondonandSouthEngland** | 0.249 | 0.124 | 0.005 | 0.492 | 0.220 | 0.129 | -0.033 | 0.473 | 0.197 | 0.131 | -0.060 | 0.454 | | | | |
| **i ~ coupleandchildren** | -0.066 | 0.151 | -0.363 | 0.230 | -0.104 | 0.155 | -0.409 | 0.200 | 0.108 | 0.154 | -0.195 | 0.410 | | | | |
| **i ~ empl_maternity** | 0.641 | 0.407 | -0.157 | 1.440 | 0.343 | 0.403 | -0.446 | 1.132 | 0.810 | 0.409 | 0.009 | 1.611 | | | | |
| **i ~ empl_retired** | -0.454 | 0.167 | -0.782 | -0.127 | | | | | | | | | | | | |
| **i ~ empl_self** | -0.168 | 0.206 | -0.573 | 0.237 | | | | | | | | | | | | |
| **i ~ empl_sick** | 5.789 | 0.558 | 4.695 | 6.883 | | | | | | | | | | | | |
| **i ~ empl_student** | -0.278 | 0.323 | -0.911 | 0.355 | | | | | | | | | | | | |
| **i ~ empl_unemployed** | 1.877 | 0.450 | 0.995 | 2.759 | | | | | | | | | | | | |
| **i ~ FinancialUncertainty2019** | 2.123 | 0.189 | 1.753 | 2.493 | 2.337 | 0.199 | 1.946 | 2.727 | 2.331 | 0.201 | 1.937 | 2.726 | 2.209 | 0.203 | 1.812 | 2.606 |
| **i ~ IMD_1** | -0.162 | 0.217 | -0.587 | 0.264 | 0.020 | 0.229 | -0.428 | 0.468 | | | | | | | | |
| **i ~ IMD_2** | 0.220 | 0.192 | -0.157 | 0.596 | 0.256 | 0.198 | -0.133 | 0.645 | | | | | | | | |
| **i ~ IMD_4** | -0.084 | 0.162 | -0.401 | 0.233 | -0.086 | 0.167 | -0.414 | 0.242 | | | | | | | | |
| **i ~ IMD_5** | -0.102 | 0.154 | -0.403 | 0.200 | -0.123 | 0.157 | -0.431 | 0.185 | | | | | | | | |
| **i ~ isced** | -0.059 | 0.152 | -0.358 | 0.239 | -0.097 | 0.153 | -0.397 | 0.204 | -0.408 | 0.158 | -0.717 | -0.099 | | | | |
| **i ~ own_mortgage** | 0.080 | 0.143 | -0.199 | 0.360 | 0.236 | 0.143 | -0.045 | 0.517 | | | | | | | | |
| **i ~ own_rent** | 1.064 | 0.183 | 0.706 | 1.422 | 1.612 | 0.188 | 1.244 | 1.980 | | | | | | | | |
| **i ~ save** | -0.815 | 0.116 | -1.043 | -0.587 | -0.992 | 0.119 | -1.225 | -0.760 | | | | | | | | |
| **i ~ sex_female** | 1.065 | 0.122 | 0.826 | 1.304 | 1.048 | 0.126 | 0.801 | 1.295 | 1.034 | 0.129 | 0.781 | 1.286 | | | | |
| **i ~ single** | 0.365 | 0.164 | 0.045 | 0.686 | 0.559 | 0.169 | 0.228 | 0.890 | 1.139 | 0.174 | 0.797 | 1.481 | | | | |
| **i ~ singleandchildren** | 0.142 | 0.359 | -0.561 | 0.844 | 0.230 | 0.375 | -0.506 | 0.965 | 1.070 | 0.377 | 0.331 | 1.809 | | | | |
| **i ~~ i** | 15.114 | 0.646 | 13.849 | 16.379 | 16.498 | 0.678 | 15.170 | 17.827 | 17.256 | 0.725 | 15.834 | 18.677 | 18.584 | 0.762 | 17.091 | 20.077 |
| **i ~~ s** | -0.939 | 0.239 | -1.407 | -0.471 | -1.040 | 0.244 | -1.518 | -0.562 | -1.076 | 0.253 | -1.573 | -0.579 | -1.017 | 0.258 | -1.522 | -0.511 |

| | | | | | | | | | | | | | | | | | | | |
|---|---|---|---|---|---|---|---|---|---|---|---|---|---|---|---|---|---|---|---|
| **s ~ age** | -0.002 | 0.002 | -0.006 | 0.002 | | -0.002 | 0.002 | -0.005 | 0.001 | | -0.002 | 0.001 | -0.005 | 0.001 | | | | | |
| **s ~ country_LondonandSouthEngland** | -0.044 | 0.046 | -0.134 | 0.046 | | -0.040 | 0.047 | -0.132 | 0.051 | | -0.047 | 0.046 | -0.137 | 0.043 | | | | | |
| **s ~ coupleandchildren** | 0.095 | 0.059 | -0.020 | 0.210 | | 0.101 | 0.059 | -0.014 | 0.216 | | 0.064 | 0.057 | -0.048 | 0.177 | | | | | |
| **s ~ empl_20_maternity** | -0.190 | 0.165 | -0.513 | 0.133 | | | | | | | | | | | | | | | |
| **s ~ empl_20_retired** | 0.078 | 0.098 | -0.113 | 0.270 | | | | | | | | | | | | | | | |
| **s ~ empl_20_self** | -0.102 | 0.144 | -0.385 | 0.181 | | | | | | | | | | | | | | | |
| **s ~ empl_20_sick** | -1.151 | 0.253 | -1.647 | -0.656 | | | | | | | | | | | | | | | |
| **s ~ empl_20_student** | -0.072 | 0.150 | -0.366 | 0.223 | | | | | | | | | | | | | | | |
| **s ~ empl_20_unemployed** | -0.217 | 0.162 | -0.534 | 0.100 | | | | | | | | | | | | | | | |
| **s ~ empl_21_maternity** | 0.207 | 0.168 | -0.123 | 0.536 | | | | | | | | | | | | | | | |
| **s ~ empl_21_retired** | 0.043 | 0.128 | -0.208 | 0.294 | | | | | | | | | | | | | | | |
| **s ~ empl_21_self** | 0.327 | 0.153 | 0.027 | 0.626 | | | | | | | | | | | | | | | |
| **s ~ empl_21_sick** | -0.014 | 0.296 | -0.594 | 0.566 | | | | | | | | | | | | | | | |
| **s ~ empl_21_student** | 0.060 | 0.201 | -0.333 | 0.453 | | | | | | | | | | | | | | | |
| **s ~ empl_21_unemployed** | -0.032 | 0.143 | -0.313 | 0.249 | | | | | | | | | | | | | | | |
| **s ~ empl_22_maternity** | 0.176 | 0.217 | -0.249 | 0.600 | | | | | | | | | | | | | | | |
| **s ~ empl_22_retired** | -0.028 | 0.114 | -0.251 | 0.194 | | | | | | | | | | | | | | | |
| **s ~ empl_22_self** | -0.133 | 0.137 | -0.401 | 0.136 | | | | | | | | | | | | | | | |
| **s ~ empl_22_sick** | 1.091 | 0.339 | 0.427 | 1.755 | | | | | | | | | | | | | | | |
| **s ~ empl_22_student** | 0.279 | 0.201 | -0.115 | 0.674 | | | | | | | | | | | | | | | |
| **s ~ empl_22_unemployed** | 0.671 | 0.160 | 0.358 | 0.985 | | | | | | | | | | | | | | | |
| **s ~ empl_maternity** | -0.258 | 0.167 | -0.586 | 0.069 | | -0.175 | 0.135 | -0.440 | 0.090 | | -0.204 | 0.135 | -0.468 | 0.060 | | | | | |
| **s ~ FinancialUncertainty2019** | -0.329 | 0.072 | -0.470 | -0.188 | | -0.334 | 0.073 | -0.478 | -0.191 | | -0.334 | 0.073 | -0.477 | -0.190 | | -0.311 | 0.073 | -0.454 | -0.167 |
| **s ~ FinancialHardship2020to2022_Constant_uncertainty** | 1.037 | 0.111 | 0.821 | 1.254 | | 1.082 | 0.116 | 0.854 | 1.310 | | 1.067 | 0.118 | 0.836 | 1.298 | | 0.936 | 0.119 | 0.703 | 1.170 |
| **s ~ FinancialHardship2020to2022_b+3_uncertainty** | 0.473 | 0.059 | 0.357 | 0.588 | | 0.507 | 0.059 | 0.391 | 0.623 | | 0.506 | 0.060 | 0.388 | 0.624 | | 0.452 | 0.061 | 0.333 | 0.572 |
| **s ~ FinancialHardship2020to2022_b+1 and b+3_uncertainty** | 0.646 | 0.107 | 0.437 | 0.855 | | 0.654 | 0.112 | 0.434 | 0.873 | | 0.638 | 0.113 | 0.416 | 0.860 | | 0.563 | 0.115 | 0.337 | 0.790 |
| **s ~ FinancialHardship2020to2022_b+1 uncertainty** | 0.317 | 0.096 | 0.129 | 0.505 | | 0.328 | 0.095 | 0.143 | 0.514 | | 0.314 | 0.096 | 0.126 | 0.503 | | 0.298 | 0.099 | 0.103 | 0.493 |
| **s ~ FinancialHardship2020to2022_b+1 and b+2 uncertainty** | 0.750 | 0.148 | 0.460 | 1.040 | | 0.753 | 0.151 | 0.456 | 1.049 | | 0.742 | 0.153 | 0.441 | 1.043 | | 0.720 | 0.157 | 0.412 | 1.027 |
| **s ~ FinancialHardship2020to2022_b+2 and b+3 uncertainty** | 0.829 | 0.117 | 0.600 | 1.057 | | 0.830 | 0.117 | 0.602 | 1.059 | | 0.829 | 0.116 | 0.601 | 1.056 | | 0.754 | 0.115 | 0.528 | 0.979 |
| **s ~ FinancialHardship2020to2022_b+2 uncertainty** | 0.483 | 0.106 | 0.275 | 0.690 | | 0.493 | 0.109 | 0.280 | 0.707 | | 0.476 | 0.107 | 0.265 | 0.686 | | 0.442 | 0.109 | 0.228 | 0.656 |
| **s ~ IMD_1** | 0.124 | 0.077 | -0.026 | 0.274 | | 0.123 | 0.078 | -0.030 | 0.275 | | | | | | | | | | |
| **s ~ IMD_2** | 0.093 | 0.073 | -0.051 | 0.237 | | 0.103 | 0.074 | -0.041 | 0.248 | | | | | | | | | | |
| **s ~ IMD_4** | 0.002 | 0.061 | -0.118 | 0.121 | | -0.002 | 0.061 | -0.122 | 0.117 | | | | | | | | | | |
| **s ~ IMD_5** | -0.011 | 0.057 | -0.124 | 0.101 | | -0.007 | 0.057 | -0.120 | 0.105 | | | | | | | | | | |
| **s ~ isced** | 0.104 | 0.062 | -0.019 | 0.226 | | 0.089 | 0.063 | -0.034 | 0.211 | | 0.102 | 0.063 | -0.022 | 0.225 | | | | | |
| **s ~ own_mortgage** | -0.035 | 0.051 | -0.136 | 0.065 | | -0.072 | 0.052 | -0.173 | 0.029 | | | | | | | | | | |
| **s ~ own_rent** | -0.102 | 0.068 | -0.235 | 0.032 | | -0.121 | 0.066 | -0.250 | 0.009 | | | | | | | | | | |
| **s ~ save** | 0.167 | 0.042 | 0.085 | 0.248 | | 0.159 | 0.042 | 0.075 | 0.242 | | | | | | | | | | |
| **s ~ sex_female** | 0.120 | 0.045 | 0.033 | 0.207 | | 0.119 | 0.045 | 0.032 | 0.206 | | 0.119 | 0.045 | 0.031 | 0.206 | | | | | |
| **s ~ single** | -0.079 | 0.057 | -0.192 | 0.034 | | -0.086 | 0.057 | -0.199 | 0.026 | | -0.107 | 0.056 | -0.217 | 0.004 | | | | | |
| **s ~ singleandchildren** | -0.018 | 0.126 | -0.266 | 0.230 | | -0.001 | 0.124 | -0.245 | 0.243 | | -0.043 | 0.122 | -0.283 | 0.196 | | | | | |
| **s ~~ s** | 0.876 | 0.123 | 0.636 | 1.117 | | 0.866 | 0.123 | 0.625 | 1.107 | | 0.867 | 0.124 | 0.623 | 1.111 | | 0.868 | 0.126 | 0.621 | 1.114 |

#### Financial hardship and uncertainty (cross-sectional baseline)

| | Employment adjustment | | | |
|---|---|---|---|---|
| **term** | **Estimate** | **SE** | **CI lower** | **CI upper** |
| **ghq ~~ ghq** | 10.778 | 0.487 | 9.824 | 11.732 |
| **ghq_20 ~~ ghq_20** | 14.051 | 0.431 | 13.207 | 14.896 |
| **ghq_21 ~~ ghq_21** | 14.024 | 0.387 | 13.266 | 14.783 |
| **ghq_22 ~~ ghq_22** | 10.874 | 0.494 | 9.906 | 11.841 |
| **i ~ age** | -0.035 | 0.005 | -0.044 | -0.025 |
| **i ~ country_LondonandSouthEngland** | 0.213 | 0.104 | 0.009 | 0.417 |
| **i ~ coupleandchildren** | -0.281 | 0.133 | -0.542 | -0.020 |
| **i ~ empl_maternity** | 0.428 | 0.325 | -0.210 | 1.065 |
| **i ~ empl_retired** | -0.117 | 0.146 | -0.404 | 0.170 |
| **i ~ empl_self** | -0.211 | 0.180 | -0.563 | 0.142 |
| **i ~ empl_sick** | 5.211 | 0.490 | 4.252 | 6.171 |
| **i ~ empl_student** | -0.061 | 0.273 | -0.596 | 0.474 |
| **i ~ empl_unemployed** | 1.432 | 0.383 | 0.681 | 2.183 |
| **i ~ FinancialUncertainty2019** | 1.754 | 0.170 | 1.421 | 2.087 |
| **i ~ FinancialHardship2019** | 2.765 | 0.146 | 2.479 | 3.051 |
| **i ~ IMD_1** | -0.219 | 0.187 | -0.584 | 0.147 |
| **i ~ IMD_2** | 0.139 | 0.163 | -0.180 | 0.458 |
| **i ~ IMD_4** | 0.049 | 0.144 | -0.233 | 0.331 |
| **i ~ IMD_5** | 0.038 | 0.138 | -0.232 | 0.309 |
| **i ~ isced** | 0.099 | 0.137 | -0.168 | 0.367 |
| **i ~ own_mortgage** | -0.032 | 0.129 | -0.284 | 0.220 |
| **i ~ own_rent** | 0.555 | 0.169 | 0.224 | 0.886 |
| **i ~ save** | -0.072 | 0.102 | -0.272 | 0.129 |
| **i ~ sex_female** | 1.006 | 0.101 | 0.808 | 1.205 |
| **i ~ single** | 0.121 | 0.140 | -0.153 | 0.395 |
| **i ~ singleandchildren** | -0.186 | 0.316 | -0.804 | 0.433 |
| **i ~~ i** | 13.012 | 0.494 | 12.044 | 13.981 |
| **i ~~ s** | -0.823 | 0.177 | -1.169 | -0.476 |
| **s ~ age** | -0.001 | 0.002 | -0.005 | 0.002 |
| **s ~ country_LondonandSouthEngland** | -0.049 | 0.040 | -0.127 | 0.028 |
| **s ~ coupleandchildren** | 0.098 | 0.053 | -0.005 | 0.202 |
| **s ~ empl_20_maternity** | -0.110 | 0.146 | -0.396 | 0.176 |
| **s ~ empl_20_retired** | 0.071 | 0.087 | -0.099 | 0.241 |
| **s ~ empl_20_self** | -0.097 | 0.128 | -0.348 | 0.155 |
| **s ~ empl_20_sick** | -1.050 | 0.224 | -1.489 | -0.612 |
| **s ~ empl_20_student** | 0.003 | 0.143 | -0.277 | 0.283 |
| **s ~ empl_20_unemployed** | -0.299 | 0.144 | -0.582 | -0.017 |
| **s ~ empl_21_maternity** | 0.192 | 0.150 | -0.103 | 0.487 |
| **s ~ empl_21_retired** | 0.011 | 0.109 | -0.203 | 0.225 |
| **s ~ empl_21_self** | 0.277 | 0.140 | 0.003 | 0.551 |
| **s ~ empl_21_sick** | -0.104 | 0.256 | -0.605 | 0.397 |
| **s ~ empl_21_student** | 0.030 | 0.191 | -0.345 | 0.405 |
| **s ~ empl_21_unemployed** | -0.070 | 0.131 | -0.328 | 0.187 |
| **s ~ empl_22_maternity** | 0.162 | 0.194 | -0.217 | 0.542 |
| **s ~ empl_22_retired** | -0.007 | 0.103 | -0.208 | 0.194 |
| **s ~ empl_22_self** | -0.113 | 0.127 | -0.362 | 0.136 |
| **s ~ empl_22_sick** | 1.065 | 0.282 | 0.511 | 1.618 |
| **s ~ empl_22_student** | 0.312 | 0.194 | -0.067 | 0.692 |
| **s ~ empl_22_unemployed** | 0.636 | 0.153 | 0.336 | 0.935 |
| **s ~ empl_maternity** | -0.242 | 0.145 | -0.526 | 0.043 |

| | | | | |
|---|---|---|---|---|
| **s ~ FinancialUncertainty2019** | -0.306 | 0.065 | -0.434 | -0.179 |
| **s ~ FinancialHardship2020to2022_Constant_uncertainty** | 0.798 | 0.101 | 0.599 | 0.996 |
| **s ~ FinancialHardship2020to2022_b+3_uncertainty** | 0.361 | 0.054 | 0.256 | 0.467 |
| **s ~ FinancialHardship2020to2022_b+1 and b+3_uncertainty** | 0.506 | 0.096 | 0.317 | 0.694 |
| **s ~ FinancialHardship2020to2022_b+1 uncertainty** | 0.236 | 0.080 | 0.079 | 0.393 |
| **s ~ FinancialHardship2020to2022_b+1 and b+2 uncertainty** | 0.519 | 0.131 | 0.263 | 0.775 |
| **s ~ FinancialHardship2020to2022_b+2 and b+3 uncertainty** | 0.663 | 0.102 | 0.464 | 0.862 |
| **s ~ FinancialHardship2020to2022_b+2 uncertainty** | 0.447 | 0.088 | 0.274 | 0.620 |
| **s ~ FinancialHardship2019** | -0.616 | 0.066 | -0.745 | -0.487 |
| **s ~ FinancialHardship2020to2022_Constant_hardship** | 0.692 | 0.077 | 0.542 | 0.842 |
| **s ~ FinancialHardship2020to2022_b+3_hardship** | 0.587 | 0.089 | 0.412 | 0.761 |
| **s ~ FinancialHardship2020to2022_b+1 and b+3_hardship** | 0.442 | 0.117 | 0.213 | 0.670 |
| **s ~ FinancialHardship2020to2022_b+1 hardship** | 0.192 | 0.080 | 0.035 | 0.350 |
| **s ~ FinancialHardship2020to2022_b+1 and b+2 hardship** | 0.192 | 0.099 | -0.002 | 0.386 |
| **s ~ FinancialHardship2020to2022_b+2 and b+3 hardship** | 0.849 | 0.119 | 0.615 | 1.084 |
| **s ~ FinancialHardship2020to2022_b+2 hardship** | 0.204 | 0.106 | -0.004 | 0.412 |
| **s ~ IMD_1** | 0.090 | 0.070 | -0.047 | 0.226 |
| **s ~ IMD_2** | 0.089 | 0.065 | -0.037 | 0.216 |
| **s ~ IMD_4** | -0.017 | 0.055 | -0.125 | 0.092 |
| **s ~ IMD_5** | -0.019 | 0.053 | -0.123 | 0.086 |
| **s ~ isced** | 0.101 | 0.057 | -0.010 | 0.212 |
| **s ~ own_mortgage** | -0.032 | 0.048 | -0.127 | 0.062 |
| **s ~ own_rent** | -0.096 | 0.065 | -0.222 | 0.031 |
| **s ~ save** | 0.102 | 0.039 | 0.025 | 0.179 |
| **s ~ sex_female** | 0.108 | 0.038 | 0.033 | 0.183 |
| **s ~ single** | -0.033 | 0.052 | -0.134 | 0.068 |
| **s ~ singleandchildren** | -0.030 | 0.120 | -0.265 | 0.206 |
| **s ~~ s** | 0.844 | 0.095 | 0.659 | 1.030 |

#### Financial hardship and uncertainty (cross-sectional baseline and ipw for attrition)

| | Employment adjustment | | | |
|---|---|---|---|---|
| **term** | **Estimate** | **SE** | **CI lower** | **CI upper** |
| **ghq ~~ ghq** | 11.804 | 0.634 | 10.561 | 13.047 |
| **ghq_20 ~~ ghq_20** | 14.946 | 0.513 | 13.940 | 15.951 |
| **ghq_21 ~~ ghq_21** | 14.703 | 0.431 | 13.858 | 15.549 |
| **ghq_22 ~~ ghq_22** | 11.710 | 0.573 | 10.586 | 12.833 |
| **i ~ age** | -0.036 | 0.005 | -0.047 | -0.025 |
| **i ~ country_LondonandSouthEngland** | 0.216 | 0.120 | -0.019 | 0.452 |
| **i ~ coupleandchildren** | -0.320 | 0.148 | -0.610 | -0.029 |
| **i ~ empl_maternity** | 0.688 | 0.410 | -0.116 | 1.491 |
| **i ~ empl_retired** | -0.156 | 0.160 | -0.471 | 0.158 |
| **i ~ empl_self** | -0.172 | 0.201 | -0.567 | 0.222 |
| **i ~ empl_sick** | 5.270 | 0.530 | 4.230 | 6.309 |
| **i ~ empl_student** | -0.102 | 0.310 | -0.709 | 0.504 |
| **i ~ empl_unemployed** | 1.358 | 0.429 | 0.518 | 2.198 |
| **i ~ FinancialUncertainty2019** | 1.677 | 0.187 | 1.310 | 2.044 |
| **i ~ FinancialHardship2019** | 2.835 | 0.166 | 2.510 | 3.159 |
| **i ~ IMD_1** | -0.208 | 0.211 | -0.622 | 0.205 |
| **i ~ IMD_2** | 0.199 | 0.186 | -0.166 | 0.563 |
| **i ~ IMD_4** | 0.003 | 0.160 | -0.310 | 0.315 |
| **i ~ IMD_5** | 0.038 | 0.151 | -0.257 | 0.334 |
| **i ~ isced** | 0.075 | 0.146 | -0.211 | 0.361 |
| **i ~ own_mortgage** | -0.092 | 0.139 | -0.363 | 0.180 |
| **i ~ own_rent** | 0.591 | 0.178 | 0.243 | 0.940 |
| **i ~ save** | -0.146 | 0.114 | -0.369 | 0.078 |
| **i ~ sex_female** | 1.058 | 0.118 | 0.827 | 1.289 |
| **i ~ single** | 0.141 | 0.157 | -0.167 | 0.450 |
| **i ~ singleandchildren** | -0.244 | 0.354 | -0.937 | 0.449 |
| **i ~~ i** | 13.928 | 0.598 | 12.755 | 15.101 |
| **i ~~ s** | -0.908 | 0.228 | -1.355 | -0.461 |
| **s ~ age** | -0.002 | 0.002 | -0.006 | 0.002 |
| **s ~ country_LondonandSouthEngland** | -0.045 | 0.046 | -0.135 | 0.044 |
| **s ~ coupleandchildren** | 0.106 | 0.058 | -0.007 | 0.220 |
| **s ~ empl_20_maternity** | -0.191 | 0.164 | -0.512 | 0.131 |
| **s ~ empl_20_retired** | 0.045 | 0.095 | -0.142 | 0.232 |
| **s ~ empl_20_self** | -0.122 | 0.145 | -0.406 | 0.162 |
| **s ~ empl_20_sick** | -1.063 | 0.244 | -1.541 | -0.585 |
| **s ~ empl_20_student** | -0.045 | 0.145 | -0.330 | 0.240 |
| **s ~ empl_20_unemployed** | -0.211 | 0.158 | -0.521 | 0.099 |
| **s ~ empl_21_maternity** | 0.202 | 0.167 | -0.125 | 0.529 |
| **s ~ empl_21_retired** | 0.050 | 0.128 | -0.201 | 0.300 |
| **s ~ empl_21_self** | 0.322 | 0.153 | 0.022 | 0.623 |
| **s ~ empl_21_sick** | -0.035 | 0.290 | -0.604 | 0.533 |
| **s ~ empl_21_student** | 0.023 | 0.196 | -0.361 | 0.408 |
| **s ~ empl_21_unemployed** | -0.037 | 0.141 | -0.313 | 0.240 |
| **s ~ empl_22_maternity** | 0.195 | 0.219 | -0.234 | 0.624 |
| **s ~ empl_22_retired** | -0.003 | 0.114 | -0.226 | 0.220 |
| **s ~ empl_22_self** | -0.141 | 0.138 | -0.412 | 0.130 |
| **s ~ empl_22_sick** | 1.018 | 0.332 | 0.367 | 1.668 |
| **s ~ empl_22_student** | 0.253 | 0.199 | -0.137 | 0.642 |
| **s ~ empl_22_unemployed** | 0.575 | 0.160 | 0.262 | 0.889 |
| **s ~ empl_maternity** | -0.316 | 0.172 | -0.654 | 0.022 |
| **s ~ FinancialUncertainty2019** | -0.284 | 0.073 | -0.426 | -0.141 |

| | | | | |
|---|---|---|---|---|
| **s ~ FinancialHardship2020to2022_Constant_uncertainty** | 0.856 | 0.112 | 0.636 | 1.075 |
| **s ~ FinancialHardship2020to2022_b+3_uncertainty** | 0.380 | 0.059 | 0.264 | 0.495 |
| **s ~ FinancialHardship2020to2022_b+1 and b+3_uncertainty** | 0.538 | 0.104 | 0.333 | 0.742 |
| **s ~ FinancialHardship2020to2022_b+1 uncertainty** | 0.265 | 0.092 | 0.084 | 0.446 |
| **s ~ FinancialHardship2020to2022_b+1 and b+2 uncertainty** | 0.622 | 0.147 | 0.335 | 0.910 |
| **s ~ FinancialHardship2020to2022_b+2 and b+3 uncertainty** | 0.669 | 0.114 | 0.445 | 0.892 |
| **s ~ FinancialHardship2020to2022_b+2 uncertainty** | 0.415 | 0.107 | 0.206 | 0.623 |
| **s ~ FinancialHardship2019** | -0.632 | 0.073 | -0.775 | -0.489 |
| **s ~ FinancialHardship2020to2022_Constant_hardship** | 0.702 | 0.083 | 0.540 | 0.864 |
| **s ~ FinancialHardship2020to2022_b+3_hardship** | 0.631 | 0.097 | 0.441 | 0.821 |
| **s ~ FinancialHardship2020to2022_b+1 and b+3_hardship** | 0.435 | 0.126 | 0.187 | 0.683 |
| **s ~ FinancialHardship2020to2022_b+1 hardship** | 0.219 | 0.088 | 0.047 | 0.391 |
| **s ~ FinancialHardship2020to2022_b+1 and b+2 hardship** | 0.212 | 0.111 | -0.006 | 0.431 |
| **s ~ FinancialHardship2020to2022_b+2 and b+3 hardship** | 0.905 | 0.127 | 0.657 | 1.153 |
| **s ~ FinancialHardship2020to2022_b+2 hardship** | 0.183 | 0.113 | -0.039 | 0.405 |
| **s ~ IMD_1** | 0.108 | 0.076 | -0.041 | 0.257 |
| **s ~ IMD_2** | 0.091 | 0.073 | -0.053 | 0.234 |
| **s ~ IMD_4** | -0.010 | 0.061 | -0.129 | 0.109 |
| **s ~ IMD_5** | -0.008 | 0.057 | -0.121 | 0.104 |
| **s ~ isced** | 0.116 | 0.062 | -0.005 | 0.238 |
| **s ~ own_mortgage** | -0.024 | 0.051 | -0.124 | 0.076 |
| **s ~ own_rent** | -0.119 | 0.068 | -0.252 | 0.014 |
| **s ~ save** | 0.134 | 0.043 | 0.050 | 0.218 |
| **s ~ sex_female** | 0.123 | 0.044 | 0.036 | 0.209 |
| **s ~ single** | -0.053 | 0.057 | -0.165 | 0.060 |
| **s ~ singleandchildren** | -0.016 | 0.129 | -0.269 | 0.238 |
| **s ~~ s** | 0.858 | 0.120 | 0.623 | 1.093 |
